# Prevalence and factors associated with multidrug resistant *Mycobacterium tuberculosis* infection in Cameroon: a systematic review and meta-analysis

**DOI:** 10.64898/2026.07.13.26357969

**Authors:** Fabrice Zobel Lekeumo Cheuyem, Chabeja Achangwa, Paul Etoga Mbarga, Rick Tchamani, Solange Dabou, Henri Donald Mutarambirwa, Mazou Ngou Temgoua

## Abstract

**Background:** Multidrug-resistant tuberculosis (MDR-TB) remains a significant public health threat in low-and middle-income countries, including Cameroon. This systematic review and meta-analysis aimed to determine the pooled prevalence of MDR-TB and other specific anti-tuberculosis drug resistance patterns, as well as to identify factors associated with drug-resistant tuberculosis in Cameroon.

**Methods:** A comprehensive literature search was conducted in PubMed, Scopus, Web of Science, Embase, Cochrane Library, and African Journals Online. Additional studies were identified through Google Scholar and reference list screening. Observational studies (cross-sectional, cohort, and case-control) reporting drug resistance among bacteriologically confirmed tuberculosis patients in Cameroon were eligible. Joanna Briggs Institute critical appraisal tools were used to critically assessed the study quality. Pooled prevalence estimates were calculated using random-effects meta-analysis. Subgroup analyses and meta-regression explored sources of heterogeneity. A *p*-value 11 0.05 was considered statistically significant.

**Results:** Twenty-eight studies conducted between 1995 and 2022 were included. The pooled prevalence of MDR-TB was 5.2% (95% CI: 2.7-9.6; 21 studies; n = 7,515), with significantly higher acquired resistance (11.6%; 95% CI: 6.3-20.3) than initial resistance (2.0%; 95% CI: 1.1-3.5). The highest pooled MDR-TB prevalence was observed in the most recent studies (38.8%; 95% CI: 33.7-44.2), and the lowest in 2015-2019 (2.7%; 95% CI: 0.4-15.2). Any resistance to anti-tuberculosis drugs was 16.0% (95% CI: 10.3-23.9; 28 studies; n = 9,931), and rifampicin resistance was 4.6% (95% CI: 2.4-8.6; 25 studies; n = 8,728). Monoresistance was highest for streptomycin (6.4%; 95% CI: 3.7-10.8) and isoniazid (4.7%; 95% CI: 3.0-7.4). Previous tuberculosis infection was the strongest predictor of drug resistance (OR = 3.9; 95% CI: 1.8-8.4), followed by alcohol consumption (OR = 1.8; 95% CI: 1.2-2.7) and history of incarceration (OR = 1.7; 95% CI: 1.1-2.6). High heterogeneity was observed across most of the pooled estimates.

**Conclusions:** Drug-resistant tuberculosis, particularly MDR-TB, poses a substantial burden in Cameroon, with acquired resistance significantly exceeding initial resistance. Previous tuberculosis infection, alcohol use, and incarceration are key modifiable risk factors. These findings underscore the urgent need to strengthen routine drug susceptibility testing, scale up rapid molecular diagnostics, enhance treatment adherence strategies, and implement targeted interventions for high-risk populations.

## 1. Introduction

Tuberculosis (TB) is still a major cause of morbidity and mortality around the world, especially in low-and middle-income countries where the burden has been consistently high over the years. In 2022, the World Health Organization (WHO) reported that TB was responsible for about 10.6 million new cases and 1.3 million deaths worldwide, showing its ongoing and persistent impact on public health [1, 2].

Multidrug-Resistant Tuberculosis (MDR-TB) is a form of TB that shows resistance to at least isoniazid and rifampin. It is a global threat and a challenge for TB control worldwide, particularly in lower-and middle-income countries, where the burden remains high. MDR-TB usually requires longer treatment, costs more, has lower success rates, and leads to more deaths than drug-susceptible TB [3, 4]. According to World Health Organization (WHO) estimates, approximately 390,000 incident cases of MDR/RR-TB occurred worldwide in 2024, with an estimated 150,000 associated deaths. More than half of the global burden was concentrated in India (32%), China (7.1%), the Philippines (7.1%), and the Russian Federation (6.7%) [5].

Sub-Saharan Africa bears a substantial burden of tuberculosis (TB) and drug-resistant tuberculosis (DR-TB), driven by delayed diagnosis, limited access to drug susceptibility testing, weak health systems, and high rates of HIV co-infection [6, 7]. HIV infection substantially increases the risk of progression from latent to active TB and has been associated with poorer treatment outcomes, including drug resistance [7]. According to recent WHO estimates, the number of individuals diagnosed with drug-resistant tuberculosis in 2024 varied considerably across the region, with approximately 3,662 cases in Nigeria, 2,099 cases in Democratic Republic of the Congo, 835 cases in Ethiopia, and 122 cases in Burkina Faso [8]. Previous anti-tuberculosis treatment remains the most consistently identified predictor of multidrug-resistant tuberculosis (MDR-TB) in the region, while other commonly reported associated factors include previous TB episodes, poor treatment adherence, treatment interruption, contact with known MDR-TB cases, delayed diagnosis, socioeconomic disadvantage, and healthcare-related exposure history of previous tuberculosis [9].

In Cameroon, an estimated 6,810 tuberculosis (TB) deaths were reported in 2024, corresponding to a TB mortality rate of 23 per 100,000 population, while 188 people were diagnosed with drug-resistant TB in the same year [10]. Studies conducted in the country have identified several potential risk factors for multidrug-resistant tuberculosis (MDR-TB), including previous TB treatment, treatment failure, poor adherence to therapy, HIV co-infection, and close contact with individuals diagnosed with MDR-TB [11].

However, these findings are not always consistent across studies due to differences in study design, population characteristics, diagnostic methods, and study settings [10, 11]. This lack of pooled evidence limits informed decision-making for TB control programs, resource allocation, and policy development. This systematic review and meta-analysis will help fill this gap by giving combined estimates of MDR-TB rates and identifying related risk factors in Cameroon. This systematic review and meta-analysis therefore aim to address this gap by providing pooled estimates of MDR-TB prevalence and synthesizing associated risk factors in Cameroon.

## 2. Methods

### 2.1. Study design

This review was reported in accordance with the Preferred Reporting Items for Systematic Reviews and Meta-Analyses (PRISMA) guidelines. It was registered with the International Prospective Register of Systematic Reviews under the number CRD420261391710 [12].

### 2.2. Eligibility criteria

#### Inclusion criteria

Studies conducted in Cameroon reporting data on tuberculosis patients with confirmed multidrug-resistant tuberculosis (MDR-TB) prevalence were eligible. Participants included individuals of any age or sex diagnosed with tuberculosis, including new and previously treated cases. MDR-TB must have been confirmed using standard diagnostic methods, such as culture and drug susceptibility testing (DST), performed phenotypically or by molecular diagnosis. Eligible study designs were observational studies (cross-sectional, cohort, and case-control) and randomized controlled trials. Studies published in English or French were considered. There was no restriction on study timeframe.

### Exclusion criteria

Studies without primary data (reviews, editorials, commentaries), duplicate publications, and studies lacking sufficient data (abstract only) to calculate prevalence or assess methodological quality were excluded. Multi-country studies without extractable Cameroon-specific data were disregarded.

### 2.3. Search strategy

#### Databases

A comprehensive literature search was conducted in PubMed, Embase, Scopus, Web of Science, and African Journals Online (AJOL). Additional studies will be identified through Google Scholar searches and screening the reference lists of included studies.

#### Search terms

The search strategy was combined Medical Subject Headings (MeSH) and free-text terms using Boolean operators: (“multidrug-resistant tuberculosis” OR “MDR-TB” OR “drug-resistant tuberculosis” OR “rifampicin-resistant tuberculosis”) AND (“prevalence” OR “epidemiology” OR “risk factors” OR “associated factors” OR “determinants”) AND (“Cameroon” OR “Cameroun”). Study selection was performed independently by two reviewers (FZLC and RT) (**Table S1**). In cases of disagreement, consensus was reached through discussion or by consulting a third reviewer (MNT). The last search was conducted on April 15, 2026.

### 2.4. Data extraction

Data were extracted by two independent reviewers (FZLC and CA) using a standardized extraction form developed in Microsoft Excel. Cases of disagreement were resolved through discussion or by contacting a third reviewer (MNT). For each included study, the following data were extracted: first author surname, publication year, year of study completion, design, region of study, sample size, number of MDR-TB cases, number of monoresistance cases, diagnostic methods, participant characteristics, and reported risk factors with corresponding effect estimates (number of cases among confirmed and control groups for each risk factor).

### 2.5. Outcomes measurement

The prevalence was calculated by multiplying the number of positive events (antimicrobial resistance cases) by 100 and dividing by the total number of TB patients screened for drug susceptibility testing using either phenotypic or molecular diagnosis. Participants tested. MDR-TB was defined as a form of TB that shows resistance to at least isoniazid and rifampin [13]. Mono-resistance was resistance to one specific anti-TB drug only. Any antituberculosis drug resistance was defined as any resistance to at least one antituberculosis drug. Any resistance to rifampicin referred to as rifampicin-monoresistance, and/or MDR-TB patients [14, 15].

### 2.6. Risk of bias assessment

Two independent reviewers (FZLC and CA) assessed study quality and risk of bias using the Joanna Briggs Institute (JBI) critical appraisal tools tailored to each study design. Disagreements were resolved through discussion or by consulting a third reviewer (MNT). For the cross-sectional study, the criteria included: a clear definition of inclusion criteria, comprehensive descriptions of study subjects and settings, validity and reliability of exposure measurements, use of objective, standardized criteria for outcome assessment, identification of potential confounding factors, implementation of appropriate strategies to address them, and the appropriateness of statistical methods. Each criterion was scored as 1 (yes) or 0 (no or unclear). In this study, we defined and applied the following categorization of the overall risk of bias: low = 7-9, moderate = 6-5, or high = 3-0.

### 2.7. Statistical analysis

The pooled prevalence estimates were calculated for each study with 95% confidence intervals. A random-effects meta-analysis using the DerSimonian–Laird method was performed to account for potential heterogeneity, as assessed by the I² statistic. Subgroup analyses and meta-regression were conducted based on study timeframe, type of study participants, method, geographic region, type of participants, localization of TB infection, and drug susceptibility. The generalized linear mixed models (GLMM), coupled with the probit-logit transformation (PLOGIT), were utilized for their effectiveness in handling meta-analyses of binary data [16]. Statistical significance was set at a *p*-value of <0.05. All analyses were conducted using the’meta’ package in R Statistics version 4.5.3 [17]. Correlates to anti-tuberculosis drug resistance were also computed to generate pooled ORs and their corresponding 95% CIs.

### 2.8. Publication bias and sensitivity analysis

Publication bias was assessed graphically using funnel plots and statistically using Egger’s and Begg’s tests [18, 19]. A p-value below 0.05 was considered evidence of significant publication bias. In such cases, the trim-and-fill technique was employed to correct the pooled estimate. Furthermore, a leave-one-out sensitivity analysis was performed to determine the influence of individual studies on the overall findings.

## 3. Results

### 3.1. Studies selection

A systematic search of electronic databases identified 349 records from PubMed (n = 16), Scopus (n = 43), Web of Science (n = 120), Embase (n = 43), Cochrane Library (n = 11), and African Journals Online (n = 116). After removing 37 duplicate records, 312 unique records were screened based on titles and abstracts. Of these, 274 records were excluded as they did not meet the inclusion criteria.

The full texts of the remaining 38 reports were retrieved and assessed for eligibility. Following full-text review, 10 reports were excluded with reasons. Additionally, 160 records were identified through Google Scholar, and 1,777 records were identified through reference list searching of included studies and five additional studies were identified and included from these sources. Ultimately, 28 studies met the eligibility criteria and were included in the systematic review and meta-analysis (**Fig. 1**).

**Fig. 1.**
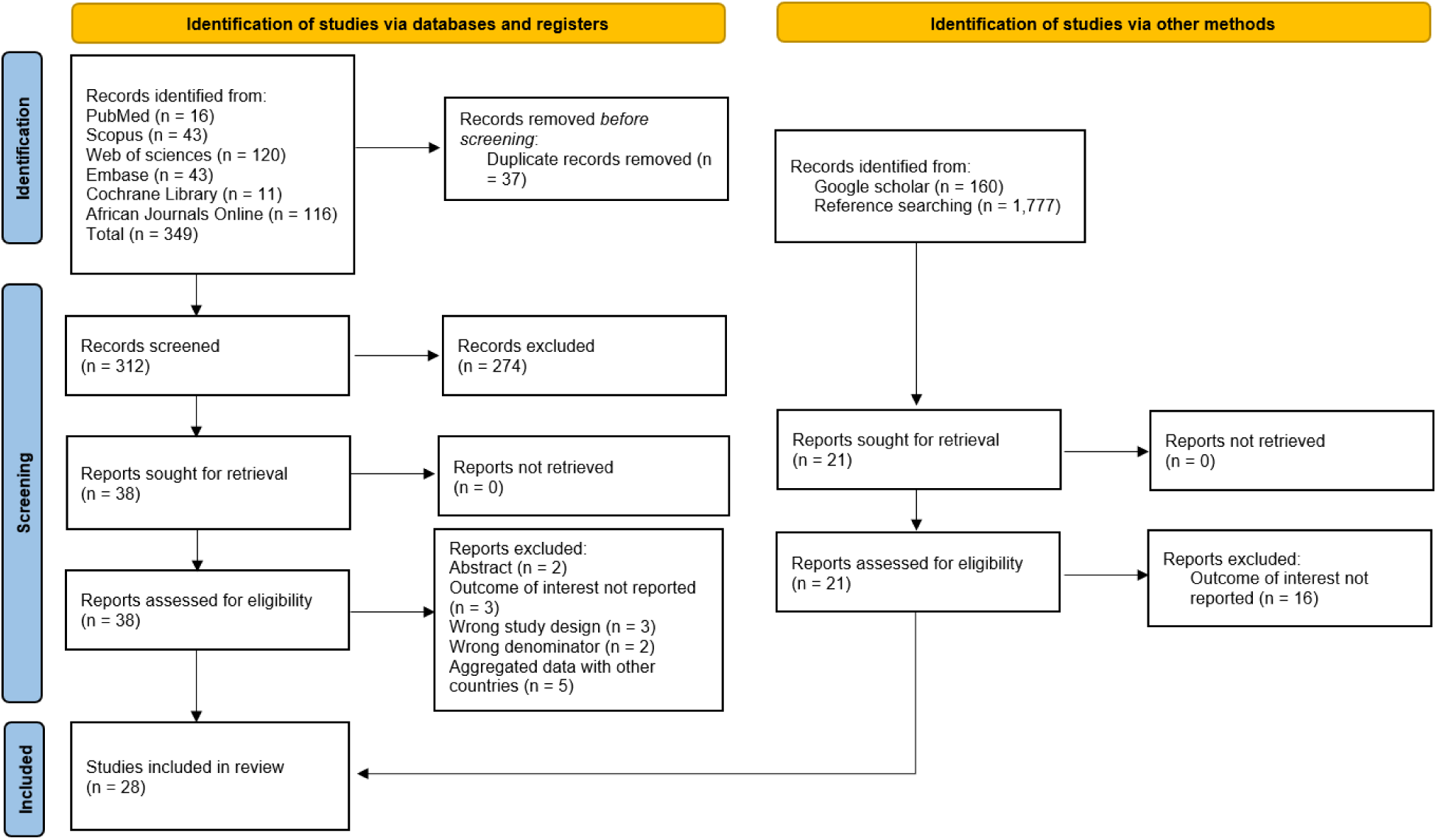
PRISMA diagram flow from study identification to inclusion in the systematic review and meta-analysis [12]

### 3.2. Studies characteristics

A total of 28 studies, conducted between 1995 and 2022 and published between 1997 and 2025, were included. Most studies were cross-sectional (n = 22; 78.6%), followed by cohort (n = 5; 17.9%) and case-control (n = 1; 3.6%). The majority of studies were conducted in the Centre (n = 9; 32.1%) and Littoral (n = 10; 35.7%) regions, while 5 studies (17.9%) were conducted in the West, South-West, and North-West regions. Twenty studies (71.4%) were conducted in a single region, while 8 studies (28.6%) covered multiple regions. Twenty-three studies (82.1%) included participants from the general population, while 4 studies (14.3%) included retreated TB patients. Twenty-four studies (85.7%) were conducted among patients with pulmonary tuberculosis; 3 studies (10.7%) included a mix of pulmonary and extrapulmonary tuberculosis patients, and one study (3.6%) focused exclusively on extrapulmonary tuberculosis. Nearly half of the studies (44.0%) used the phenotypic proportion method on Löwenstein-Jensen medium, while molecular diagnosis was used for drug susceptibility testing in 9 reports (32.1%); 1 study (3.6%) did not specify the method used. Almost all included studies (n = 27; 96.4%) had low risk of bias at methodology quality assessment (**Table 1**).

**Table 1.** Characteristics of included studies.

| Author | Study period<br>(year)<sup>1</sup> | Publication<br>(year) | Design | Region | Participant | Type of TB cases | Diagnostic method | Risk of bias | Sample size (n) | Number of resistant cases (n) |  |  |  |  |  |  |  |  |  |
| --- | --- | --- | --- | --- | --- | --- | --- | --- | --- | --- | --- | --- | --- | --- | --- | --- | --- | --- | --- |
|  |  |  |  |  |  |  |  |  |  | Any resistance | Any RIF | Mono-resistance |  |  |  |  | Multidrug resistance |  |  |
|  |  |  |  |  |  |  |  |  |  |  |  | RIF | INH | EMB | STM | FLQ | Total | Initial | Acquired |
| Bercion, 1995 [20] | 1995 | 1997 | CS | CE | PGP | PTB | PLJ | Low | 516 | 164 | 4 | 2 | 11 | 1 | 78 | NR | 3 | 3/516 | NR |
| Kuaban, 1997 [21] | 1997 | 2000 | CS | CE | Retreated TB patient | PTB | PLJ | Low | 98 | 57 | 27 | 0 | 19 | 1 | 3 | NR | 27 | NR | 27/566 |
| Kuaban, 1998 [22] | 1998 | 2000 | CS | WE | PGP | PTB | PLJ | Low | 566 | 152 | 28 | 4 | 47 | 5 | 36 | NR | 23 | 6/437 | 17/129 |
| Noeske, 1998 [23] | 1998 | 2002 | Cohort | WE | PGP | PTB | PLJ | Low | 560 | 150 | 28 | NR | NR | NR | NR | NR | 23 | 6/436 | 17/124 |
| Mbacham, 2006 [24] | 2006 | 2011 | CS | WE and CE | PGP | PTB | Dot-blot hybridization | Low | 112 | 91 | 60 | 3 | 9 | 2 | 12 | NR | 56 | NR | NR |
| Ane-Anyangwe, 2007 [25] | 2007 | 2016 | CS | SW | PGP | PTB | Dot-blot hybridization | Low | 59 | 35 | 17 | NR | NR | NR | 32 | NR | NR | NR | NR |
| Assam-Assam, 2009 [11] | 2009 | 2011 | CS | CE and WE | PGP | PTB | PLJ | Low | 149 | 16 | 3 | 1 | 4 | 0 | 4 | NR | 5 | 2/130 | 3/19 |
| Koro, 2009 [26] | 2009 | 2016 | CS | AD | PGP | PTB | PLJ | Low | 437 | 38 | 5 | 2 | 13 | 0 | 12 | NR | 6 | 6/437 | NR |
| Noeske, 2009 [27] | 2009 | 2012 | Cohort | LT | Retreated TB patient | PTB | PLJ | Low | 216 | 51 | 26 | 0 | NR | NR | NR | NR | 26 | NR | 26/216 |
| Jubulis, 2010 [28] | 2010 | 2015 | CS | NW | PGP | PTB | MYCOTB phenotypic assay | Low | 65 | 26 | 5 | 0 | 2 | 0 | 5 | 0 | 5 | NR | NR |
| Kuaban, 2010 [29] | 2010 | 2014 | CS | AD | PGP | PTB | PLJ | Low | 445 | 35 | 11 | 3 | 12 | 1 | 10 | NR | 8 | 6/426 | 2/19 |
| Sidze, 2011 [30] | 2011 | 2014 | CS | CE | PGP | PTB | PLJ | Low | 576 | 63 | 6 | 1 | 27 | 0 | 19 | 1 | 6 | 4/517 | 2/59 |
| Meriki, 2012 [31] | 2012 | 2013 | CS | NW and SW | PGP | PTB | PLJ | Low | 256 | 71 | 20 | NR | 10 | 3 | 25 | NR | 15 | NR | NR |
| Pefure-Yone, 2012 [32] | 2012 | 2014 | Cohort | CE | PGP | PTB | PLJ | Low | 46 | 7 | 2 | 0 | 5 | NR | NR | NR | 2 | 2/46 | NR |
| Sander, 2013 [33] | 2013 | 2015 | Case-control | LT, NW, SW, and WE | Retreated TB patient | PTB | MTBDRplus assay and PLJ | Low | 601 | 92 | NR | NR | NR | NR | NR | NR | 80 | NR | 80/601 |
| Sidze, 2012 [34] | 2013 | 2012 | CS | CE | PGP | PTB | Spoligotyping | Low | 298 | 28 | NR | NR | NR | NR | NR | NR | 3 | NR | NR |
| Kuaban, 2015 [35] | 2015 | 2020 | Cohort | NW, SW, WE, and LT | Retreated TB patient | PTB | Xpert MTB/RIF | Low | 753 | 47 | 0 | 0 | 47 | NR | NR | NR | NR | NR | NR |
| Abanda, 2016 [36] | 2016 | 2017 | CS | CE, LT, and NW | PGP | PTB | MTBDRplus assay | Low | 226 | 74 | 49 | 0 | 6 | 0 | 14 | NR | 49 | 7/79 | 42/169 |
| Mbuh, 2017_1 [37] | 2017 | 2021 | CS | LT | PGP | PTB and EPTB | Xpert MTB/RIF | Low | 1665 | 86 | 86 | 11 | NR | NR | NR | NR | 75 | 19/435 | 67/1230 |
| Mbuh, 2017_2 [38] | 2017 | 2019 | CS | LT | PGP | EPTB | Xpert MTB/RIF | Moderate | 45 | 0 | 0 | 0 | NR | NR | NR | NR | 0 | 0/30 | 0/45 |
| Noeske, 2017 [39] | 2017 | 2018 | CS | Multi-region | PGP | PTB | Xpert MTB/RIF | Low | 1029 | 16 | 16 | NR | NR | NR | NR | NR | NR | NR | NR |
| Pokam, 2017 [40] | 2017 | 2020 | CS | LT | PGP | PTB | Xpert MTB/RIF | Low | 147 | 13 | 13 | NR | NR | NR | NR | NR | NR | NR | NR |
| Motse, 2019 [41] | 2019 | 2020 | CS | LT | PGP | PTB and EPTB | Xpert MTB/RIF | Low | 119 | 8 | 8 | NR | NR | NR | NR | NR | NR | NR | NR |
| Tiani, 2019 [42] | 2019 | 2022 | CS | CE | PGP | PTB | Spoligotyping and Line Probe Assay | Low | 308 | 25 | 2 | 0 | 2 | 7 | 12 | NR | 2 | NR | NR |
| Djuikoue, 2020 [10] | 2020 | 2022 | CS | LT | PGP | PTB | NR | Low | 304 | 122 | NR | NR | NR | NR | NR | NR | 122 | NR | NR |
| Foncha, 2020 [43] | 2020 | 2021 | Cohort | LT | PGP | PTB and EPTB | GeneXpert | Low | 272 | 6 | 6 | NR | NR | NR | NR | NR | NR | NR | NR |
| Djoufack, 2022 [44] | 2022 | 2025 | CS | CE | PLHIV | PTB | TB-LAMP | Low | 26 | 13 | 13 | 7 | 7 | NR | NR | 0 | 6 | NR | NR |
| Pokam, 2022 [45] | 2022 | 2025 | CS | SW | PGP | PTB | GeneXpert MTB/RIF Ultra | Low | 37 | 3 | 3 | NR | NR | NR | NR | NR | NR | NR | NR |

1: Year of study completion; TB: Mycobacterium tuberculosis; RIF: Rifampicin; INH: Isoniazid; EMB: Ethambutol; STM: Streptomycin; FLQ: Fluoroquinolones; CS: Cross-sectional; CE: Centre; LT: Littoral; NW: North-West; AD: Adamawa; SW: South-West; WE: West; PGP: Patient from general population; PTB: Pulmonary tuberculosis; EPTB: Extra-pulmonary tuberculosis; PLJ: Phenotypic proportion method on Löwenstein-Jensen medium; NR: Non reported

### 3.3. Prevalence of multidrug-resistant tuberculosis

The pooled prevalence of MDR tuberculosis was 5.2% (95% CI: 2.7-9.6; 21 studies; n = 7,515), with high heterogeneity among included studies (I^2^ = 97.0%; *p* < 0.001) (**Fig. 1**).

Subgroup analysis revealed that study timeframe (*p* < 0.001), region (p < 0.001), study design (*p* = 0.005), participant type (*p* < 0.001), and drug susceptibility testing methods (*p* < 0.001) were significant sources of heterogeneity. The evolution of pooled MDR prevalence in Cameroon was inconsistent. The highest pooled prevalence was observed in the most recent (2020+) studies (38.8%; 95% CI: 33.7-44.2; 2 studies; n = 330), while the lowest was observed during 2015-2019 (2.7%; 95% CI: 0.4-15.2; 7 studies; n = 2,244). Studies conducted in multiple regions revealed a higher pooled prevalence (13.7%; 95% CI: 5.3-30.7; 5 studies; n = 1,334) compared to the Adamawa region (1.6%; 95% CI: 0.9-2.7; 2 studies; n = 882), where the lowest pooled prevalence was observed. Patients enrolled for tuberculosis retreatment showed a higher pooled prevalence (16.2%; 95% CI: 10.6-24.9; 3 studies; n = 915) compared to patients from the general population (3.7%; 95% CI: 1.8-7.6) (**Table 2 and Figs. S1-6**).

**Table 2.** Subgroup meta-analysis of multidrug-resistant *Mycobacterium tuberculosis* among tuberculosis patients in Cameroon, 1995–2022.

| Subgroup | Sample size | Prevalence<sup>1</sup> (%) | 95% CI limits<sup>1</sup> |  | Number of studies | Heterogeneity statistic<sup>1</sup> |  | Subgroup difference |
| --- | --- | --- | --- | --- | --- | --- | --- | --- |
|  |  |  | Lower | Upper |  | I<sup>2</sup>(%) | p-value | p-value |
| **Study period (year)<sup>2</sup>** |  |  |  |  |  |  |  | <0.001 |
| □2005 | 1,740 | 4.3 | 1.1 | 16.1 | 4 | 96.5 | <0.001 |  |
| 2005-2009 | 914 | 8.2 | 1.7 | 31.2 | 4 | 97.7 | <0.001 |  |
| 2010-2014 | 2,287 | 3.3 | 1.6 | 7.0 | 7 | 92.9 | <0.001 |  |
| 2015-2019 | 2,244 | 2.7 | 0.4 | 15.2 | 4 | 96.7 | <0.001 |  |
| 2020+ | 330 | 38.8 | 33.7 | 44.2 | 2 | 64.4 | 0.094 |  |
| **Study design** |  |  |  |  |  |  |  | 0.005 |
| Cross-sectional | 6,092 | 4.7 | 2.1 | 10.0 | 17 | 97.5 | <0.001 |  |
| Cohort | 822 | 6.4 | 3.3 | 12.0 | 3 | 87.2 | <0.001 |  |
| Case-control | 601 | 13.3 | 10.7 | 16.3 | 1 | - | - |  |
| **Region** |  |  |  |  |  |  |  | <0.001 |
| Adamawa | 882 | 1.6 | 0.9 | 2.7 | 2 | 0.0 | 0.615 |  |
| Centre | 1,868 | 2.7 | 0.7 | 9.3 | 7 | 95.2 | <0.001 |  |
| Littoral | 2,230 | 7.8 | 1.6 | 30.8 | 4 | 98.8 | <0.001 |  |
| North-West | 65 | 7.7 | 2.5 | 17.0 | 1 | - | - |  |
| West | 1,126 | 4.1 | 3.1 | 5.4 | 2 | 0.0 | 0.971 |  |
| Multi-region | 1,344 | 13.7 | 5.3 | 30.7 | 5 | 96.5 | <0.001 |  |
| **Participant** |  |  |  |  |  |  |  | <0.001 |
| Patient from general population | 6,574 | 3.7 | 1.8 | 7.6 | 17 | 97.5 | <0.001 |  |
| Retreated tuberculosis patient | 915 | 16.2 | 10.6 | 24.9 | 3 | 86.1 | <0.001 |  |
| People living with HIV | 26 | 23.1 | 9.0 | 43.6 | 1 | - | - |  |
| **Tuberculosis infection localization** |  |  |  |  |  |  |  | 0.479 |
| Extrapulmonary or mixed | 1,710 | 4.4 | 3.5 | 5.5 | 2 | 0.0 | 0.999 |  |
| Pulmonary | 5,805 | 5.7 | 2.9 | 10.8 | 19 | 96.8 | <0.001 |  |
| **Drug susceptibility method used** |  |  |  |  |  |  |  | <0.001 |
| Phenotypic proportion method on LJ | 3,865 | 3.5 | 1.8 | 6.7 | 11 | 93.2 | <0.001 |  |
| Molecular diagnosis | 3,346 | 6.3 | 2.0 | 17.9 | 9 | 96.9 | <0.001 |  |
| Not reported | 304 | 40.1 | 34.6 | 45.9 | 1 | - | - |  |

1: Random effects model; 2: Time frame aligns with the development of successive National Strategic Plans against tuberculosis in Cameroon; CI: Confidence interval; HIV: Human immunodeficiency virus; LJ: Lowenstein-Jensen medium

The sensitivity analysis showed that our main findings were robust; however, the funnel plot revealed asymmetry, indicating potential publication bias, as reflected by a significant Egger’s test (*p* = 0.038). The resulting adjusted pooled prevalence from the trim-and-fill analysis raised the pooled estimate to 12.8% (95% CI: 6.3-24.1; 7 imputed studies) (**Figs. S7-9**).

The pooled prevalence of acquired MDR tuberculosis was significantly higher (11.6%; 95% CI: 6.3-20.3; 11 studies; n = 2,070) than the initial prevalence (2.0%; 95% CI: 1.1-3.5; 11 studies; n = 3,345) (**Figs. 2-3**).

**Fig. 2.**
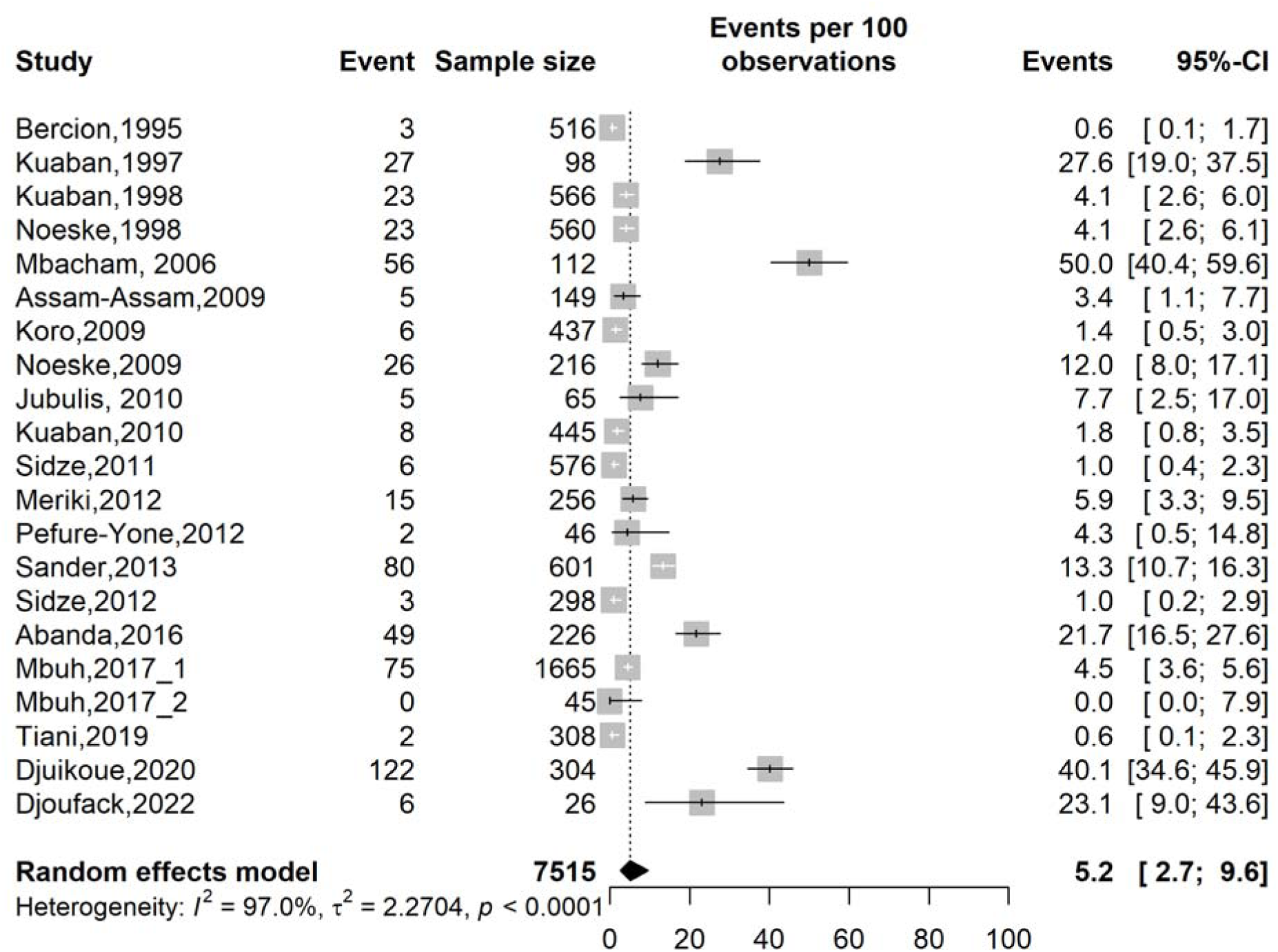
Pooled prevalence of multidrug-resistant *Mycobacterium tuberculosis* among tuberculosis patients in Cameroon, 1995–2022

**Fig. 3.**
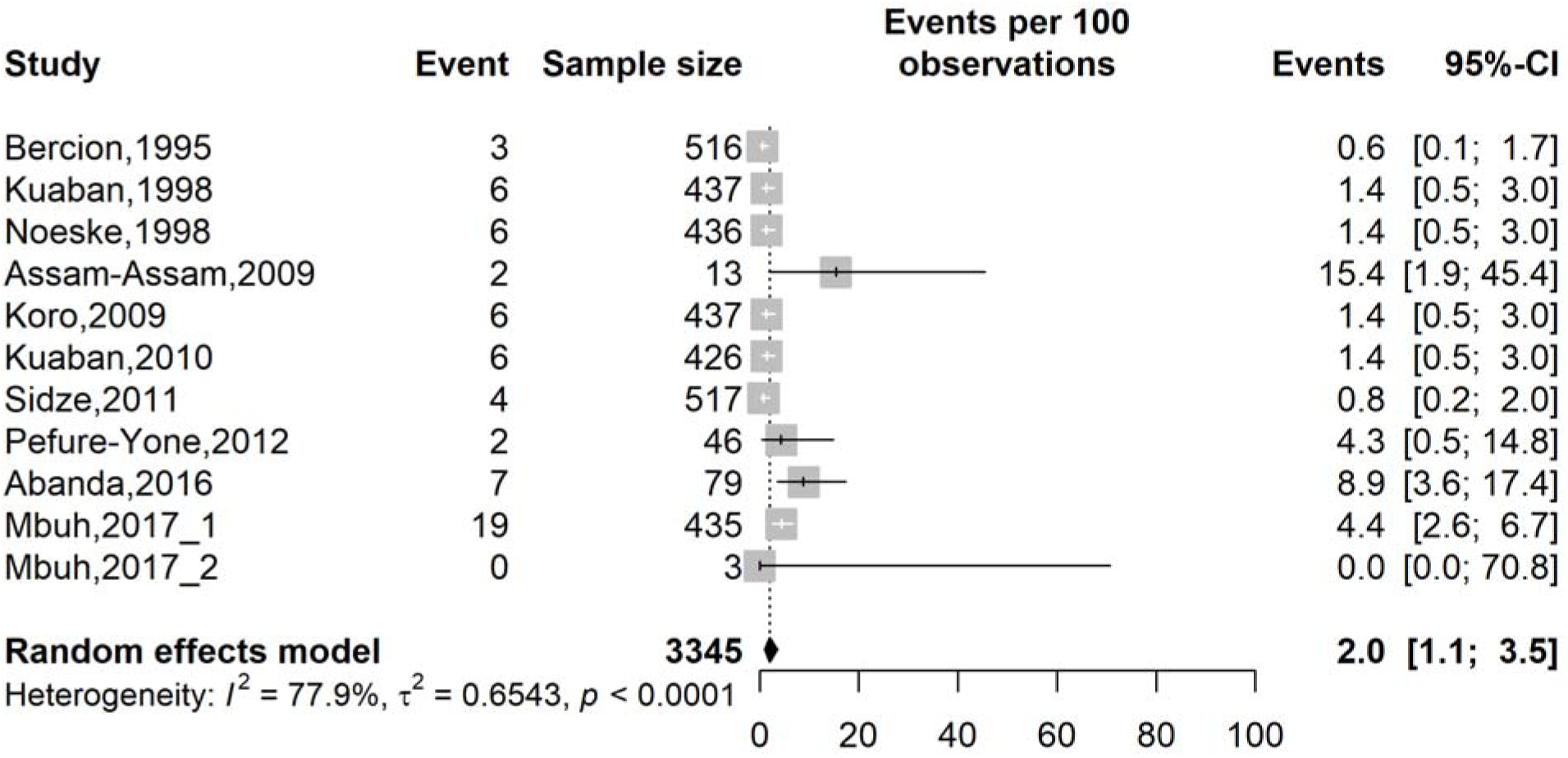
Pooled prevalence of initial multidrug-resistant *Mycobacterium tuberculosis* among new tuberculosis patients in Cameroon, 1995–2022

### 3.4. Prevalence of any resistance to anti-tuberculosis drugs

The pooled prevalence of any resistance to anti-tuberculosis drugs was 16.0% (95% CI: 10.3-23.9; 28 studies; n = 9,931), with high heterogeneity across included reports (I2 = 97.5%; *p* < 0.001) (**Fig. 4**).

**Fig. 4.**
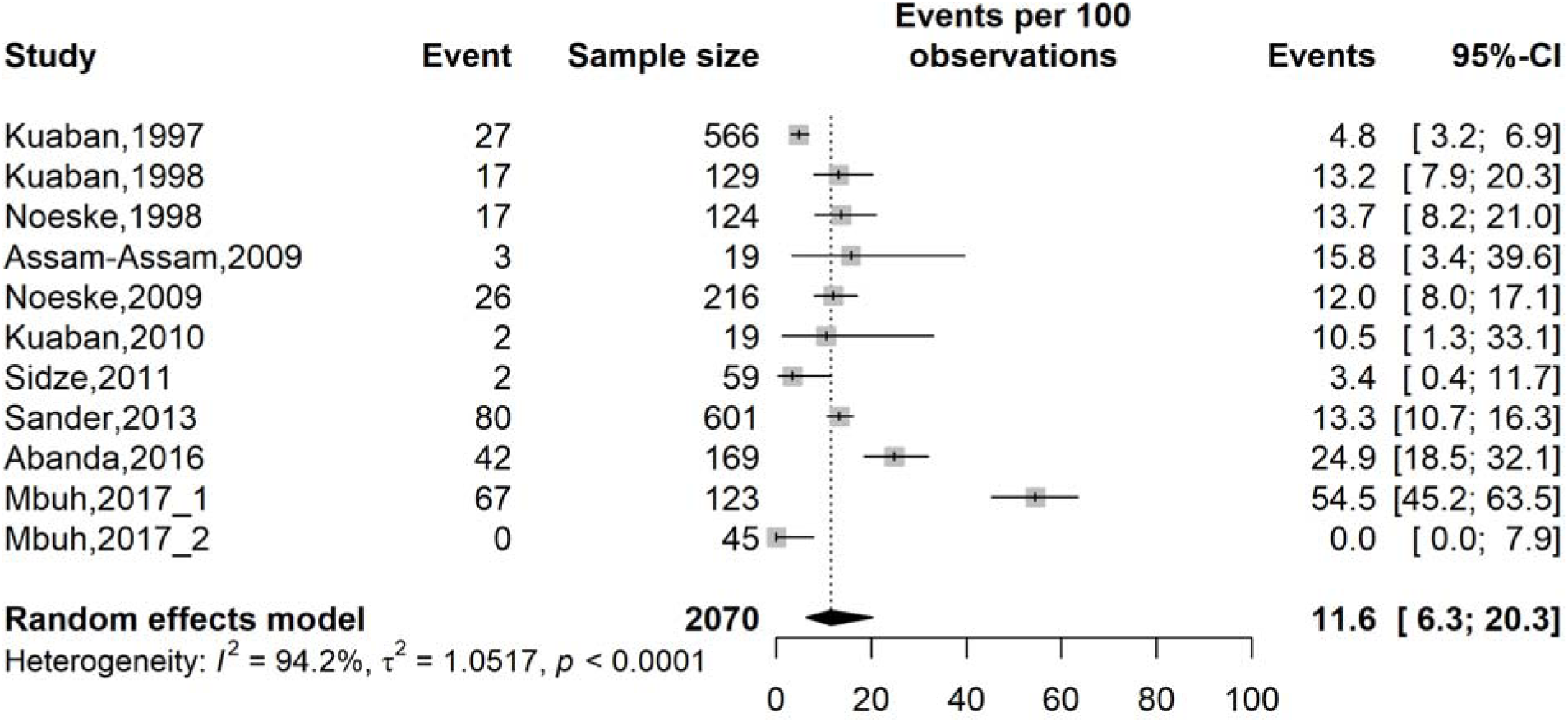
Pooled prevalence of acquired multidrug-resistant *Mycobacterium tuberculosis* among retreated tuberculosis patients in Cameroon, 1995–2022

The subgroup analysis indicated that study timeframe, region, participant type, tuberculosis infection localization, and drug susceptibility testing methods were significant sources of heterogeneity (p < 0.001). There was an overall decline in prevalence across study periods, reaching its lowest level during 2015–2019 before increasing in studies conducted from 2020 onwards. The highest pooled prevalence was recorded before 2005 (34.6%; 95% CI: 24.0-46.9; 4 studies; n = 1,740), and the lowest in 2015-2019 (6.1%; 95% CI: 3.0-12.0; 8 studies; n = 4,292). Significant disparities were observed in any resistance prevalence across Cameroonian regions, the lowest being observed in the Littoral region (7.7%; 95% CI: 2.9-18.6; 7 studies; n = 2,768) and the highest in the South-West region (27.2%; 95% CI: 4.7-73.8; 2 studies; n = 96). The prevalence of any resistance to anti-TB drugs was significantly higher among patients with pulmonary tuberculosis (20.0%; 95% CI: 13.3-29.1; 24 studies; n = 7,830) compared to the group of studies among mixed patients with either extrapulmonary or pulmonary tuberculosis (4.0%; 95% CI: 2.3-6.9; 4 studies; n = 2,101) (**Table 3 and Figs. S14-19**).

**Table 3.** Subgroup meta-analysis of the prevalence of any resistance to anti-tuberculosis drugs among tuberculosis patients in Cameroon, 1995–2022.

| Subgroup | Sample size | Prevalence<sup>1</sup><br>(%) | 95% CI limits<sup>1</sup> |  | Number of studies | Heterogeneity statistic<sup>1</sup> |  | Subgroup difference |
| --- | --- | --- | --- | --- | --- | --- | --- | --- |
|  |  |  | Lower | Upper |  | I<sup>2</sup>(%) | p-value | p-value |
| **Study period (year)<sup>2</sup>** |  |  |  |  |  |  |  | <0.001 |
| □2005 | 1,740 | 34.6 | 24.0 | 46.9 | 4 | 92.3 | <0.001 |  |
| 2005-2009 | 973 | 31.7 | 11.2 | 62.9 | 5 | 98.1 | <0.001 |  |
| 2010-2014 | 2,287 | 15.7 | 10.1 | 23.7 | 7 | 93.3 | <0.001 |  |
| 2015-2019 | 4,292 | 6.1 | 3.0 | 12.0 | 8 | 96.7 | <0.001 |  |
| 2020+ | 639 | 15.9 | 3.8 | 47.3 | 4 | 95.9 | <0.001 |  |
| **Study design** |  |  |  |  |  |  |  | 0.663 |
| Cross-sectional | 7,483 | 17.3 | 10.3 | 27.7 | 22 | 97.7 | <0.001 |  |
| Cohort | 1,847 | 11.2 | 4.8 | 23.9 | 5 | 96.8 | <0.001 |  |
| Case-control | 601 | 15.2 | 10.3 | 23.9 | 1 | - | - |  |
| **Region** |  |  |  |  |  |  |  | <0.001 |
| Adamawa | 882 | 8.3 | 6.6 | 10.3 | 2 | 0.0 | 0.655 |  |
| Centre | 1,868 | 21.4 | 11.1 | 37.3 | 7 | 96.4 | <0.001 |  |
| Littoral | 2,768 | 7.7 | 2.9 | 18.6 | 7 | 97.9 | <0.001 |  |
| North-West | 65 | 40.0 | 28.0 | 52.9 | 1 | - | - |  |
| South-West | 96 | 27.2 | 4.7 | 73.8 | 2 | 94.5 | <0.001 |  |
| West | 1,126 | 26.8 | 24.3 | 29.5 | 2 | 0.0 | 0.979 |  |
| Multi-region | 3,126 | 17.3 | 5.9 | 41.2 | 7 | 98.4 | <0.001 |  |
| **Participant** |  |  |  |  |  |  |  | 0.001 |
| Patient from general population | 8,237 | 14.3 | 8.6 | 22.7 | 23 | 97.5 | <0.001 |  |
| Retreated tuberculosis patient | 1,668 | 20.9 | 8.3 | 43.7 | 4 | 98.0 | <0.001 |  |
| People living with HIV | 26 | 50.0 | 29.9 | 70.1 | 1 | - | - |  |
| **Tuberculosis infection localization** |  |  |  |  |  |  |  | <0.001 |
| Extrapulmonary or mixed | 2,101 | 4.0 | 2.3 | 6.9 | 4 | 40.5 | 0.169 |  |
| Pulmonary | 7,830 | 20.0 | 13.3 | 29.1 | 24 | 97.2 | <0.001 |  |
| **Drug susceptibility method used** |  |  |  |  |  |  |  | <0.001 |
| Phenotypic proportion method on LJ | 3,865 | 19.9 | 13.3 | 28.6 | 11 | 95.9 | <0.001 |  |
| Molecular diagnosis | 5,762 | 12.4 | 5.9 | 24.3 | 17 | 97.6 | <0.001 |  |
| Not reported | 304 | 40.1 | 34.6 | 45.9 | 1 | - | - |  |

1: Random effects model; 2: Time frame aligns with the development of successive National Strategic Plans for the fight against tuberculosis in Cameroon; CI: Confidence interval; HIV: Human immunodeficiency virus; LJ: Lowenstein-Jensen medium

The sensitivity analysis demonstrated that no single study significantly influenced the pooled estimate, indicating its robustness. Furthermore, visual inspection showed an almost symmetrical funnel plot, with non-significant *p*-values for the Egger’s (0.414) and the Begg’s (0.527) tests (**Figs. S20-21**)

### 3.5. Prevalence of any resistance to rifampicin

The pooled prevalence of any resistance to rifampicin was 4.6% (95% CI: 2.4-8.6; 25 studies; n = 8,728) with high heterogeneity observed across studies (I^2^ = 95.6%; *p* Ill 0.001) (**Fig. 5**).

**Fig. 5.**
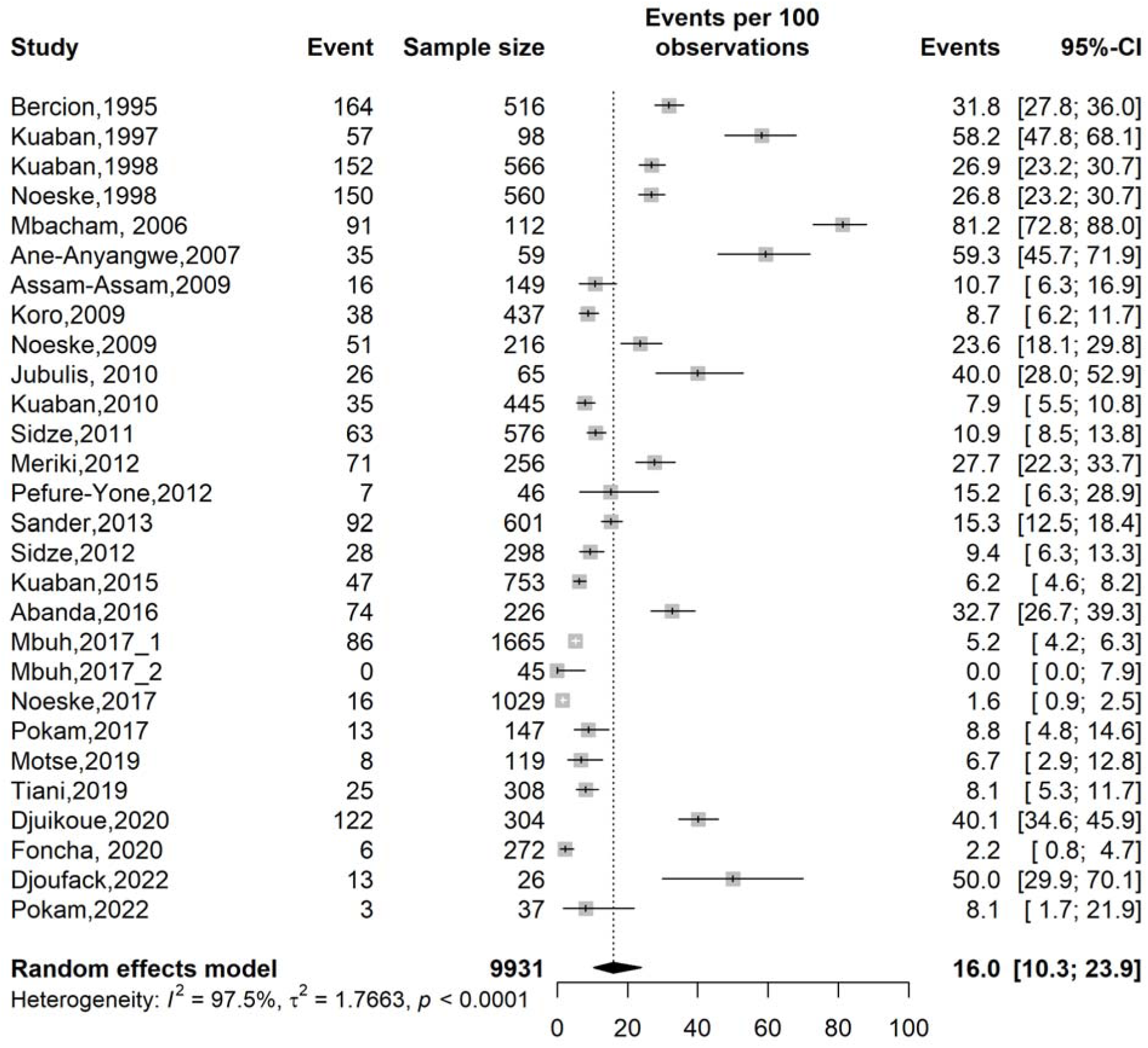
Pooled prevalence of any resistance to anti-tuberculosis drugs among tuberculosis patients in Cameroon, 1995–2022

**Fig. 6.**
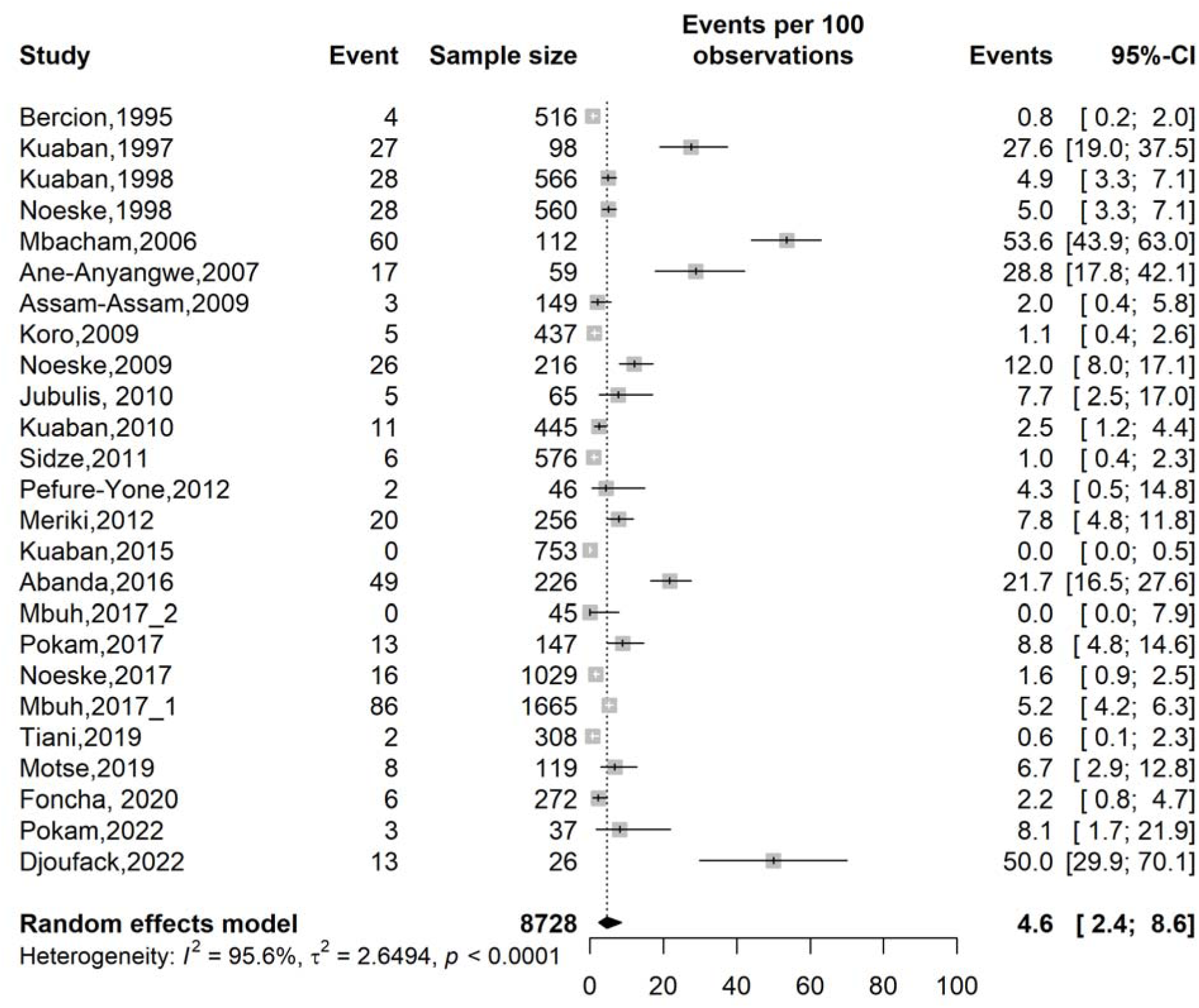
Pooled prevalence of any-resistance to rifampicin among tuberculosis patients in Cameroon, 1995–2022

The analysis of potential sources of heterogeneity revealed that the region (*p* = 0.001) and the type of participants (*p* Ill 0.001) were significant sources of observed heterogeneity. The highest prevalence was significantly higher in the South-West region of Cameroon (17.4%; 95% CI: 6.7-38.1; 2 studies; n = 96), and the lowest in the Adamawa region (1.8%; 95% CI: 1.1-3.0; 2 studies; n = 882). A single study revealed a high prevalence of any rifampicin resistance of 50.0% among HIV-TB coinfected patients. Much lower prevalences were detected among TB patients from the general population (4.2%; 95% CI: 2.4-7.4; 21 studies; n = 7,635) (**Table 4 and Figs. S22-27**).

**Table 4.**
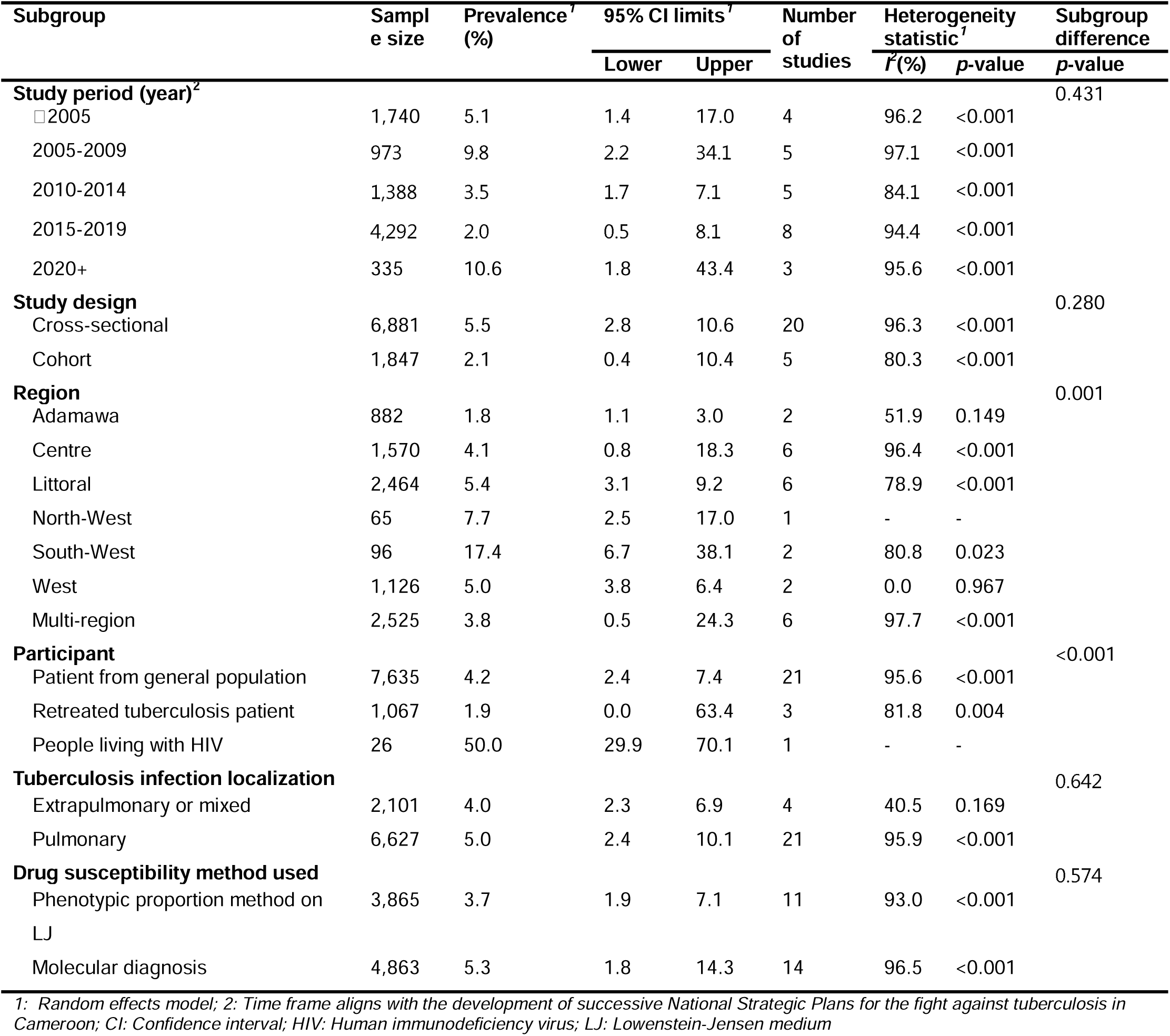
Subgroup meta-analysis of the prevalence of any-resistance to rifampicin among tuberculosis patients in Cameroon, 1995–2022.

No single study significantly influenced the pooled estimate, suggesting robustness of findings at the sensitivity analysis. In addition, no sign of publication bias was observed on the funnel plot and after performing statistical tests (*p*-value for Egger’s [0.189] and Begg’s [0.225] tests) (**Figs. S28-29**)

### 3.6. Prevalence of monoresistance to anti-tuberculosis drugs

Higher monoresistance to anti-TB drugs was observed for streptomycin (6.4%; 95% CI: 3.7-10.8; 13 studies, n = 3,813) and isoniazid (4.7%; 95% CI: 3.0-7.4; 15 studies; n = 4,579). Conversely, much lower monoresistance prevalences were recorded for rifampicin, ethambutol, and fluoroquinolones.

Most pooled estimates were robust at sensitivity analysis, with no significant risk of publication bias (*p* = 0.187-0.520). However, the pooled prevalence of ethambutol monoresistance revealed a significant risk of publication bias (*p* = 0.002 by Egger’s test), with an adjusted pooled estimate from the trim-and-fill analysis of 1.4% (95% CI: 0.7-2.6; 6 imputed studies), indicating that the initial pooled estimate was underestimated (**Table 5 and Figs. S30-45**).

**Table 5.** Psrevalence of anti-tuberculosis drug monoresistance among tuberculosis patients in Cameroon, 1995–2022.

| Drug | Sample size | Prevalence<sup>1</sup> (%) | 95% CI limits<sup>1</sup> |  | Number of studies | Heterogeneity statistic<sup>1</sup> |  | Egger's and Begg's tests | Sensitivity |
| --- | --- | --- | --- | --- | --- | --- | --- | --- | --- |
| | | | Lower | Upper | | $I^2$(%) | $p$-value | $p$-value | |
| Rifampicin | 6,249 | 0.3 | 0.1 | 0.9 | 17 | 78.9 | □ 0.001 | 0.209<br>0.187 | Robust |
| Isoniazid | 4,579 | 4.7 | 3.0 | 7.4 | 15 | 86.0 | □ 0.001 | 0.318<br>0.520 | Robust |
| Ethambutol | 3,754 | 0.3 | 0.1 | 0.9 | 12 | 0.0 | 0.540 | 0.002<br>0.273 | Robust |
| Streptomycin | 3,813 | 6.4 | 3.7 | 10.8 | 13 | 94.0 | □ 0.001 | 0.174<br>0.464 | Robust |
| Fluoroquinolones | 667 | 0.1 | 0.0 | 1.1 | 3 | 0.0 | 1.000 | NA | Robust |

1: Random effects model; CI: Confidence interval; NA: Not applicable

### 3.7. Determinants of antimicrobial resistance

A synthetic analysis of determinants of the risk of anti-tuberculosis drug resistance identified previous TB infection, alcohol consumption, and a history of incarceration as significant predictors of the occurrence of antimicrobial resistance. Previous infection was associated with a fourfold higher risk of developing resistance to anti-tuberculosis drugs (OR = 3.9; 95% CI: 1.8-8.4; 7 studies). Patients who reported alcohol consumption were 80% more at risk of developing resistance (OR = 1.8; 95% CI: 1.2-2.7; 2 studies), and those with a history of incarceration were 70% more likely to present resistance to anti-TB drugs (OR = 1.7; 95% CI: 1.1-2.6; 2 studies) (**Table 6 and Figs. S46-52**).

**Table 6.**
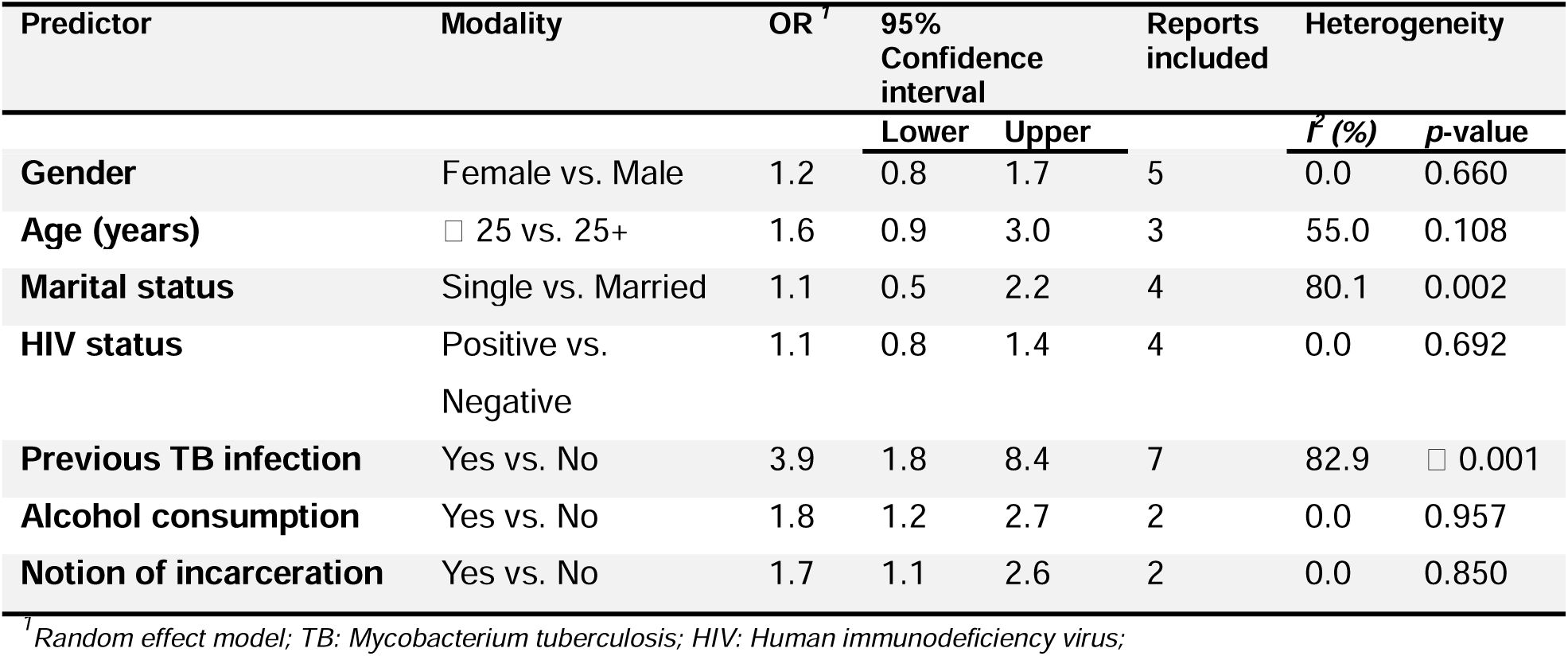
Predictors of anti-tuberculosis drug resistance among tuberculosis patients in Cameroon, 1995–2022.

| Predictor | Modality | OR<sup>†</sup> | 95% Confidence interval |  | Reports included | Heterogeneity |  |
| --- | --- | --- | --- | --- | --- | --- | --- |
|  |  |  | Lower | Upper |  | I<sup>2</sup> (%) | p-value |
| **Gender** | Female vs. Male | 1.2 | 0.8 | 1.7 | 5 | 0.0 | 0.660 |
| **Age (years)** | □ 25 vs. 25+ | 1.6 | 0.9 | 3.0 | 3 | 55.0 | 0.108 |
| **Marital status** | Single vs. Married | 1.1 | 0.5 | 2.2 | 4 | 80.1 | 0.002 |
| **HIV status** | Positive vs. Negative | 1.1 | 0.8 | 1.4 | 4 | 0.0 | 0.692 |
| **Previous TB infection** | Yes vs. No | 3.9 | 1.8 | 8.4 | 7 | 82.9 | □ 0.001 |
| **Alcohol consumption** | Yes vs. No | 1.8 | 1.2 | 2.7 | 2 | 0.0 | 0.957 |
| **Notion of incarceration** | Yes vs. No | 1.7 | 1.1 | 2.6 | 2 | 0.0 | 0.850 |

<sup>†</sup> Random effect model; TB: *Mycobacterium tuberculosis*; HIV: Human immunodeficiency virus;

## 4. Discussion

This systematic review and meta-analysis provide a comprehensive synthesis of the burden and determinants of multidrug-resistant tuberculosis (MDR-TB) and anti-tuberculosis drug resistance in Cameroon. In our analysis, the pooled prevalence of MDR-TB was 5.2%, while the prevalence of any resistance to anti-tuberculosis drugs was 16.0%. Rifampicin resistance was observed in 4.6% of patients with tuberculosis. Furthermore, previous tuberculosis infection, alcohol consumption, and a history of incarceration were identified as significant factors associated with anti-tuberculosis drug resistance. These findings indicate that drug-resistant tuberculosis remains an important public health challenge in Cameroon despite ongoing efforts to strengthen tuberculosis prevention, diagnosis, and treatment services.

The pooled MDR-TB rate found in our review is higher than the global average for new tuberculosis cases, showing that drug-resistant TB remains a major global threat in general and Cameroon in particular. The World Health Organization (WHO) estimates that 10.7 million people developed tuberculosis and 1.23 million died from it in 2024, with about 410,000 cases of multidrug-or rifampicin-resistant TB worldwide [46]. Similar MDR-TB rates have been reported in other sub-Saharan African countries. For instance, Musa *et al*. in 2017 reported a pooled prevalence of 2.1% [47]. Cameroon, like many other sub-Saharan African countries, faces ongoing challenges in TB care such as delayed diagnosis, gaps in treatment monitoring, treatment interruptions, high burden of HIV, and limited access to drug resistance testing that may still be driving the spread of resistant tuberculosis strains in Cameroon [48].

One of the major key findings of our review was that acquired MDR-TB was much more prevalent than initial MDR-TB. Our results support earlier evidence that previous anti-tuberculosis treatment is a strong risk factor for drug resistance [49]. Poor treatment adherence, interruptions, incorrect prescriptions, and delays in spotting treatment failure can all lead to more resistant strains [50]. The analysis showed that people with a history of tuberculosis infection were almost four times more likely to develop drug resistance. This result is consistent with previous studies carried out in sub-Saharan Africa, where previous tuberculosis was highlighted as one of the major risk factors [43].

Nevertheless, resistance to anti-tuberculosis drugs in our study generally dropped over time, from 34.6% before 2005 to 6.1% between 2015 and 2019. This may likely reflect better national TB programs, more directly observed treatment, improved drug availability, and better access to diagnostics [52]. Other African countries observed similar drops after improving surveillance, labs, and treatment adherence [47, 53]. However, studies after 2020 show an increase, suggesting drug resistance is still a concern. The wider use of rapid molecular tests such as GeneXpert MTB/RIF and line-probe assays in Cameroon may partly explain this, as they help detect more resistant cases that might have been missed previously with phenotypic methods [54].

Drug resistance rates vary widely across regions in Cameroon. These differences may reflect disparities in healthcare access, laboratory resources, socioeconomic factors, treatment adherence, population movement, and the level of TB control program implementation and management [55]. Urban areas in Cameroon have many specialized TB treatment centers with well-equipped laboratories that offer molecular testing, whereas rural areas rely heavily on sputum AFB microscopy for TB diagnosis, which contributes to poor characterization of resistance genes. These results underscore the need to strengthen local surveillance to identify high-burden areas and guide targeted actions. Better surveillance is especially important in resource-constrained settings, where access to diagnostics and specialized TB services can vary widely.

The pooled rate of rifampicin resistance was 4.6%, indicating that resistant strains remain in the population. This is important because rifampicin resistance is often used as a marker for MDR-TB and is typically detected by rapid molecular tests such as GeneXpert MTB/RIF [56]. Rifampicin-resistant TB leads to worse treatment outcomes, longer treatment duration, higher costs, and more deaths than drug-sensitive TB [57]. Therefore, improving access to rapid diagnostics and initiating appropriate treatment promptly are key components of TB control in Cameroon.

Streptomycin and isoniazid had the highest rates of monoresistance among first-line TB drugs. The high rate of isoniazid resistance is concerning because isoniazid is a key component of TB treatment and prevention. Studies show that failing to detect isoniazid resistance can lead to treatment failure, relapse, and more cases of multidrug-resistant TB [58, 59]. The higher rate of streptomycin resistance may reflect its past widespread use and the continued presence of resistant strains [60].

This review also found that prior TB infection, alcohol use, and incarceration are important factors linked to drug resistance. Prior TB infection is a strong risk factor for MDR-TB worldwide [49, 61]. Alcohol use can lead to an impaired immune system, which subsequently weakens the overall effect of drugs on the infectious agent and favors the emergence of resistant strains [62, 63]. Other reviews have also found a strong link between alcohol use and poor TB treatment outcomes [64]. Incarceration is another key risk factor, as prisons often have overcrowding, poor ventilation, late diagnosis, and weak infection control [65]. These findings underscore the need to strengthen TB treatment follow-up, integrate alcohol counseling and adherence support for those with regular alcohol consumption, and decongest prisons with routine screening and rapid molecular testing at entry.

## 5. Limitations

This review has some limitations. There was substantial heterogeneity across several analyses due to differences in study design, study populations, geographic coverage, and diagnostic methods. Additionally, the studies were unevenly distributed across Cameroon, limiting the representativeness of the results, and publication bias was detected in the MDR-TB prevalence analysis. Therefore, the pooled estimates should be interpreted as national approximations rather than precise measures of disease burden. However, sensitivity analyses showed that the findings were robust and not unduly influenced by any single study.

## 6. Conclusions

In summary, this review shows that drug-resistant tuberculosis remains a major challenge to tuberculosis control in Cameroon. The results highlight a high rate of MDR-TB and drug resistance, especially in previously treated patients. To reduce the impact of MDR-TB and stop the spread of resistant strains, it is important to strengthen routine drug susceptibility testing, make rapid molecular diagnostics more widely available, improve treatment adherence, strengthen surveillance systems, and focus on high-risk groups.

## Supplementary Information

Supplementary Material

## Abbreviations

AD: Adamawa region (Cameroon)
AIDS: Acquired Immunodeficiency Syndrome
AJOL: African Journals Online
CE: Centre region (Cameroon)
CI: Confidence interval
CS: Cross-sectional
DST: Drug susceptibility testing
EMB: Ethambutol
EPTB: Extrapulmonary tuberculosis
FLQ: Fluoroquinolones
GLMM: Generalized linear mixed models
HIV: Human Immunodeficiency Virus
INH: Isoniazid
JBI: Joanna Briggs Institute
LJ: Löwenstein-Jensen medium
LT: Littoral region (Cameroon)
MDR-TB: Multidrug-resistant tuberculosis
NR: Not reported
NW: North-West region (Cameroon)
OR: Odds Ratio
PGP: Patient from general population
PLHIV: People living with HIV
PLJ: Phenotypic proportion method on Löwenstein-Jensen medium
PLOGIT: Probit-Logit transformation
PRISMA: Preferred Reporting Items for Systematic Reviews and Meta-Analyses
PTB: Pulmonary tuberculosis
RIF: Rifampicin
STM: Streptomycin
SW: South-West region (Cameroon)
TB: Tuberculosis
WE: West region (Cameroon)
WHO: World Health Organization

## Data Availability

All data produced in the present work are contained in the manuscript and its supplementary materials.

## Acknowledgments

Not applicable.

## Authors’ contributions

FZLC conceived the original idea of the study; FZLC and RT conducted the literature search; FZLC, RT, CA, and MNT selected the studies, extracted the relevant information, critically assessed included reports and synthesized the data; FZLC performed the analyses; FZLC and CA wrote the first draft of the manuscript. FZLC, CA, PEM, RT, SD, HDM, and MNT critically reviewed and revised successive drafts of the manuscript. FZLC, CA, PEM, RT, SD, HDM, and MNT read and approved the final version of the manuscript.

## Data availability

All data generated in this study are available in the main manuscript and its supplementary material.

## Declarations

### Ethical approval and consent to participate

Not applicable.

### Consent for publication

Not applicable.

