## Supplementary Material for "Prevalence and factors associated with multidrug resistant *Mycobacterium tuberculosis* infection in Cameroon: a systematic review and meta-analysis"

**Table S1** Searching strategy by database

| **Database** | **Search string** | **Number of entries** |
| --- | --- | --- |
| **Pubmed** | "multidrug-resistant tuberculosis"[Title/Abstract] OR "MDR-TB"[Title/Abstract] OR "drug-resistant tuberculosis"[Title/Abstract] OR "rifampicin-resistant tuberculosis"[Title/Abstract]) AND ("prevalence"[Title/Abstract] OR "epidemiology"[Title/Abstract] OR "risk factors"[Title/Abstract] OR "associated factors"[Title/Abstract] OR "determinants"[Title/Abstract]) AND ("Cameroon"[Title/Abstract] OR "Cameroun"[Title/Abstract]) | 16 |
| **Scopus** | TITLE-ABS-KEY("multidrug-resistant tuberculosis" OR "MDR-TB" OR "drug-resistant tuberculosis" OR "rifampicin-resistant tuberculosis") AND TITLE-ABS-KEY("prevalence" OR "epidemiology" OR "risk factors" OR "associated factors" OR "determinants") AND TITLE-ABS-KEY("Cameroon" OR "Cameroun") | 43 |
| **Web of sciences** | ((((((TS = "multidrug-resistant tuberculosis") OR (TS = "MDR-TB")) OR (TS = "drug-resistant tuberculosis")) OR (TS = "rifampicin-resistant tuberculosis")) AND (((((TS = "prevalence") OR (TS = "epidemiology")) OR (TS = "risk factors")) OR (TS = "associated factors")) OR (TS = "determinants"))) AND ((TS = "Cameroon") OR (TS = "Cameroun"))) | 120 |
| **Embase** | **(**'multidrug resistant tuberculosis'/exp OR'drug resistant tuberculosis'/exp OR'rifampicin resistant tuberculosis'/exp OR  'multidrug-resistant tuberculosis':ti,ab,kw OR 'MDR-TB':ti,ab,kw OR'drug-resistant tuberculosis':ti,ab,kw OR 'rifampicin-resistant tuberculosis':ti,ab,kw) AND ('prevalence'/exp OR 'epidemiology'/exp OR 'risk factor'/exp OR 'determinant'/exp OR prevalence:ti,ab,kw OR epidemiology:ti,ab,kw OR'risk factor*':ti,ab,kw OR 'associated factor*':ti,ab,kw OR determinant*:ti,ab,kw) AND ('cameroon'/exp OR Cameroon:ti,ab,kw OR Cameroun:ti,ab,kw) | 43 |
| **Cochrane Library** | (multidrug-resistant tuberculosis OR MDR-TB OR drug-resistant tuberculosis OR rifampicin-resistant tuberculosis) AND (prevalence OR epidemiology OR risk factors OR associated factors OR determinant) AND (Cameroon OR Cameroon) | 11 |
| **AJOL** | (multidrug-resistant tuberculosis OR MDR-TB OR drug-resistant tuberculosis OR rifampicin-resistant tuberculosis) AND (prevalence OR epidemiology OR risk factors OR associated factors OR determinant) AND (Cameroon OR Cameroon) | 116 |

**Multidrug-resistant tuberculosis**

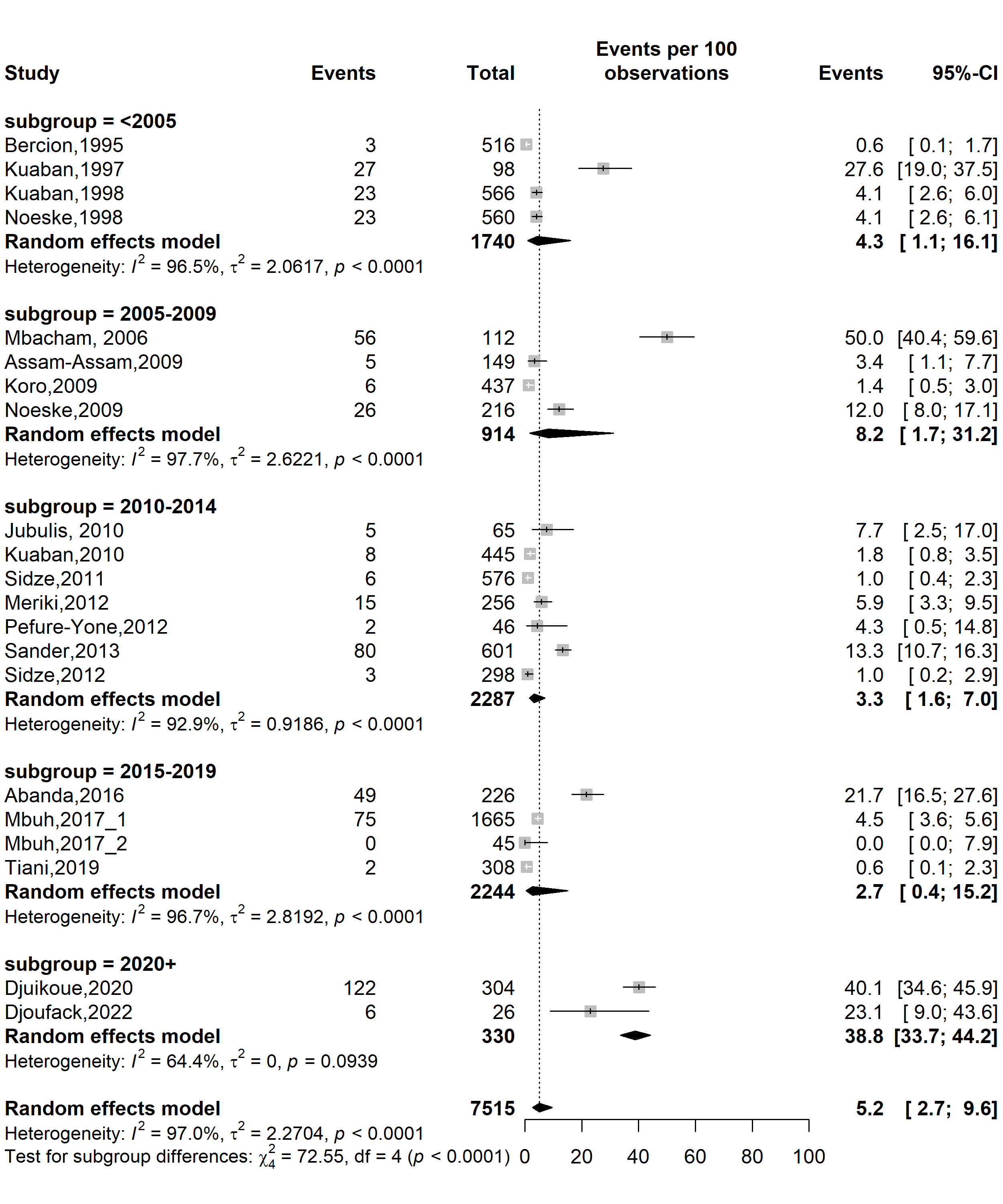

**Fig. S1** Pooled prevalence of multidrug-resistant *Mycobacterium tuberculosis* among tuberculosis patients in Cameroon by study periods, 1995–2022

*(Time frame aligns with the development of successive National Strategic Plans against tuberculosis in Cameroon)*

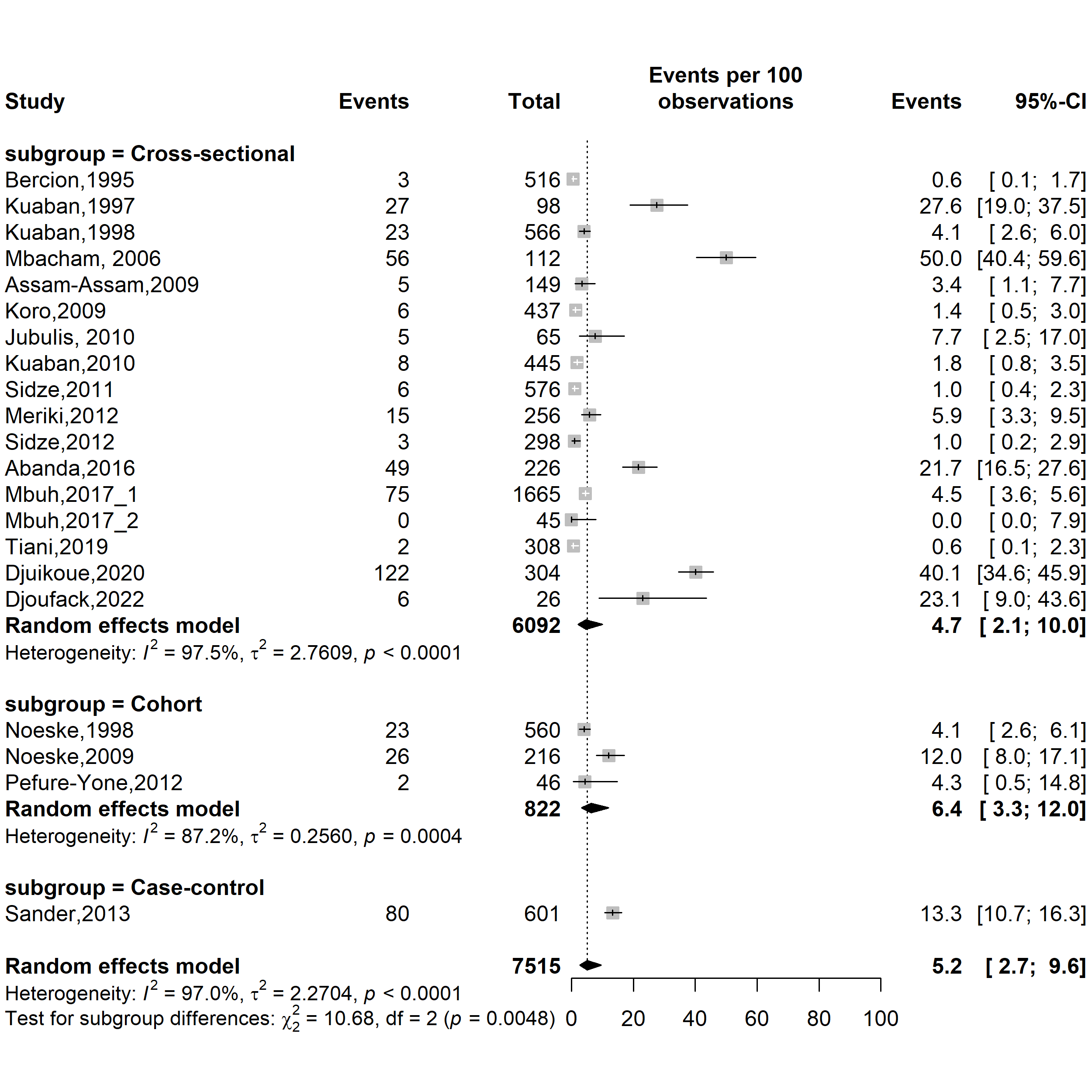

**Fig. S2** Pooled prevalence of multidrug-resistant *Mycobacterium tuberculosis* among tuberculosis patients in Cameroon by study designs, 1995–2022

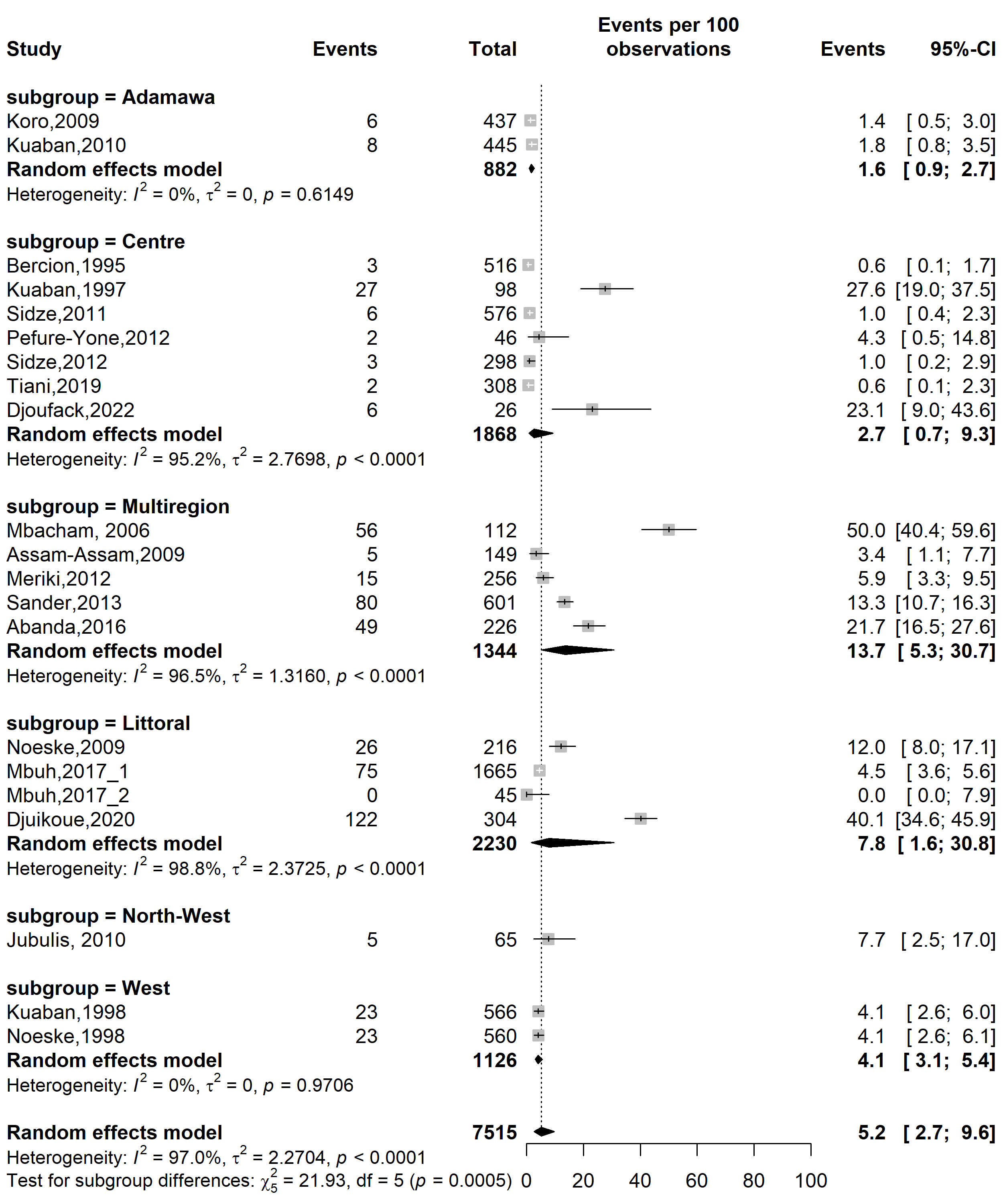

**Fig. S3** Pooled prevalence of multidrug-resistant *Mycobacterium tuberculosis* among tuberculosis patients by Cameroonian regions, 1995–2022

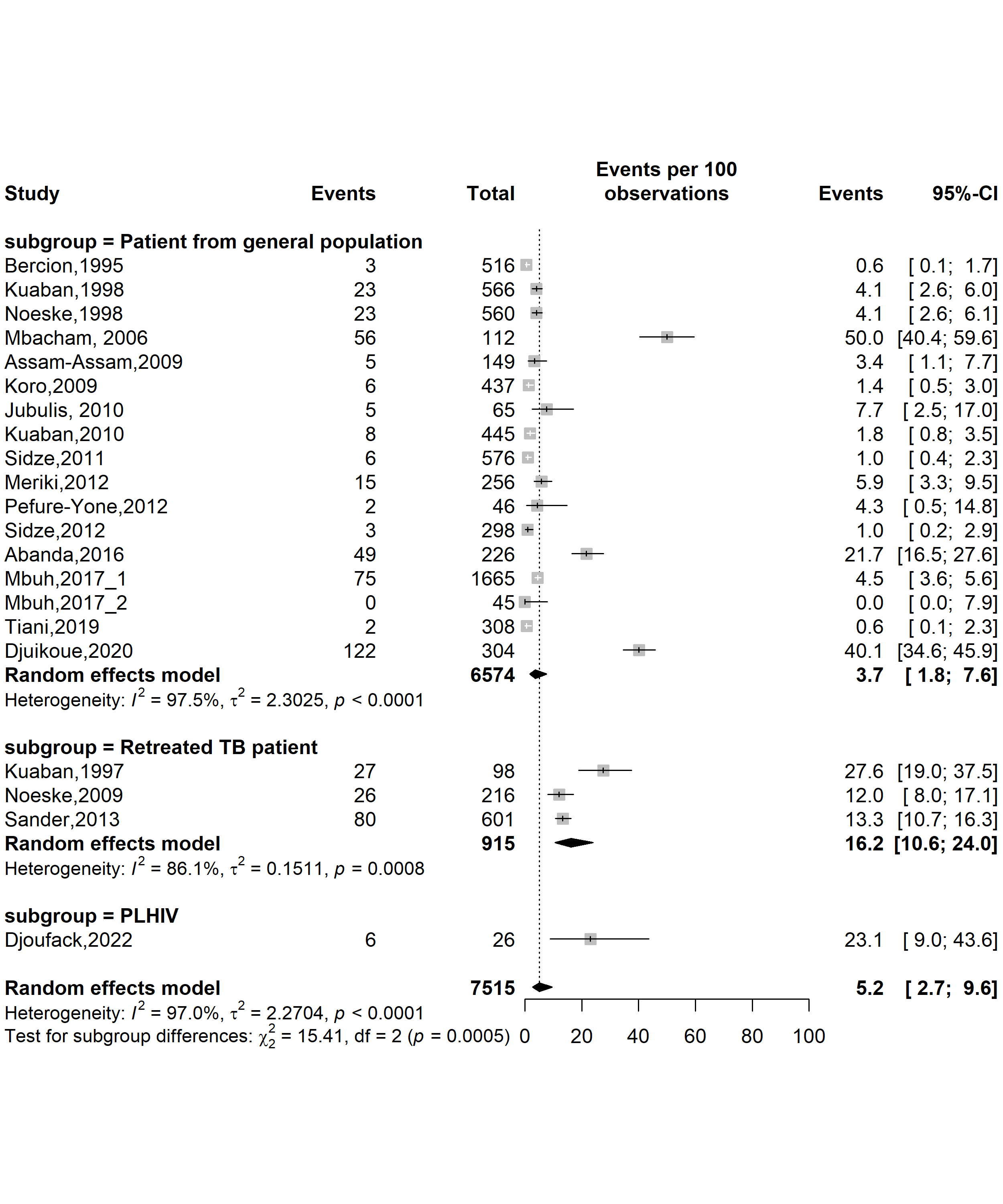

**Fig. S4** Pooled prevalence of multidrug-resistant *Mycobacterium tuberculosis* among tuberculosis patients in Cameroon by type of participants, 1995–2022

(*TB: Tuberculosis; PLHIV: People living with human immunodeficiency virus*)

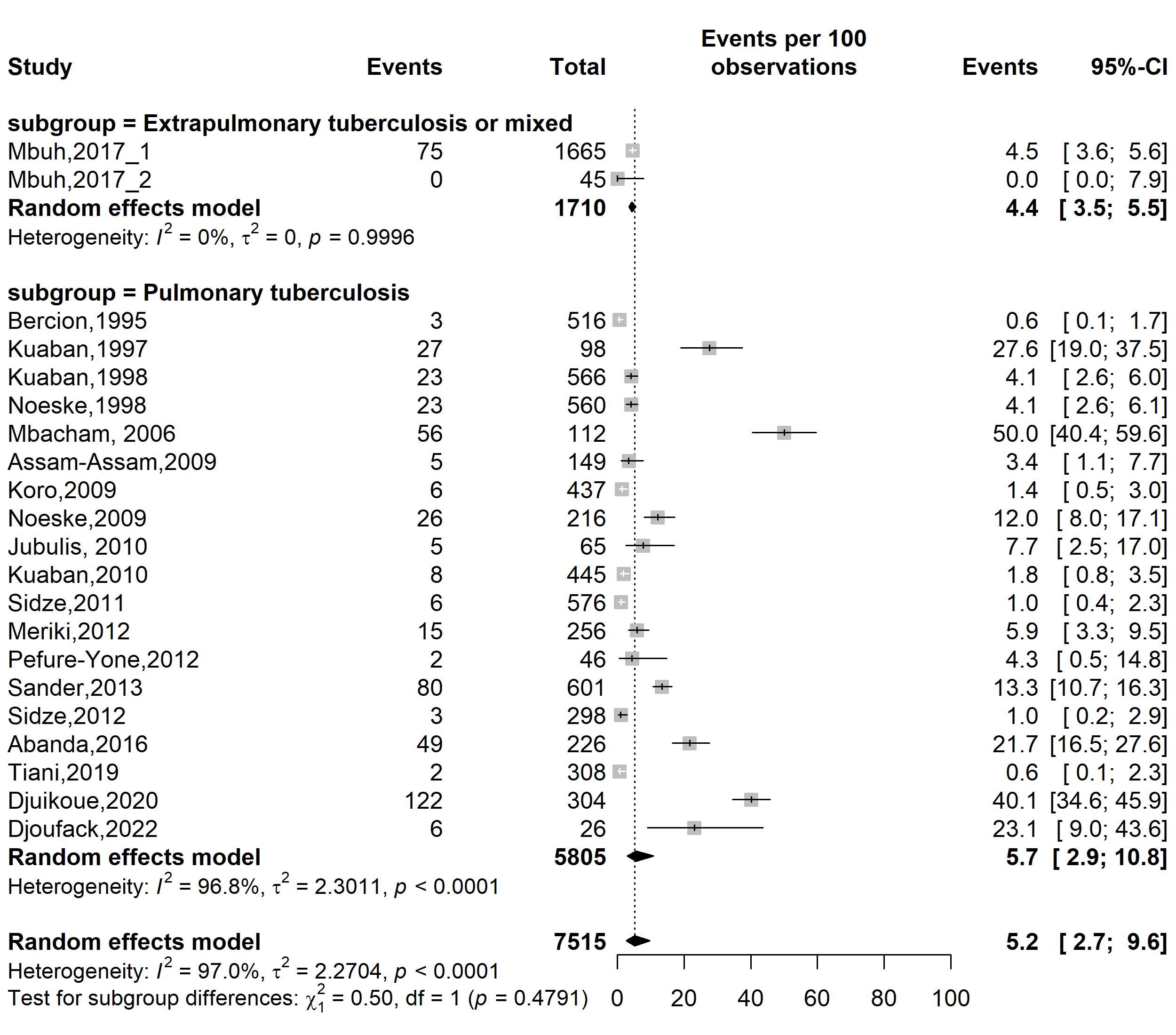

**Fig. S5** Pooled prevalence of multidrug-resistant *Mycobacterium tuberculosis* among tuberculosis patients in Cameroon by type of tuberculosis infection localizations, 1995–2022

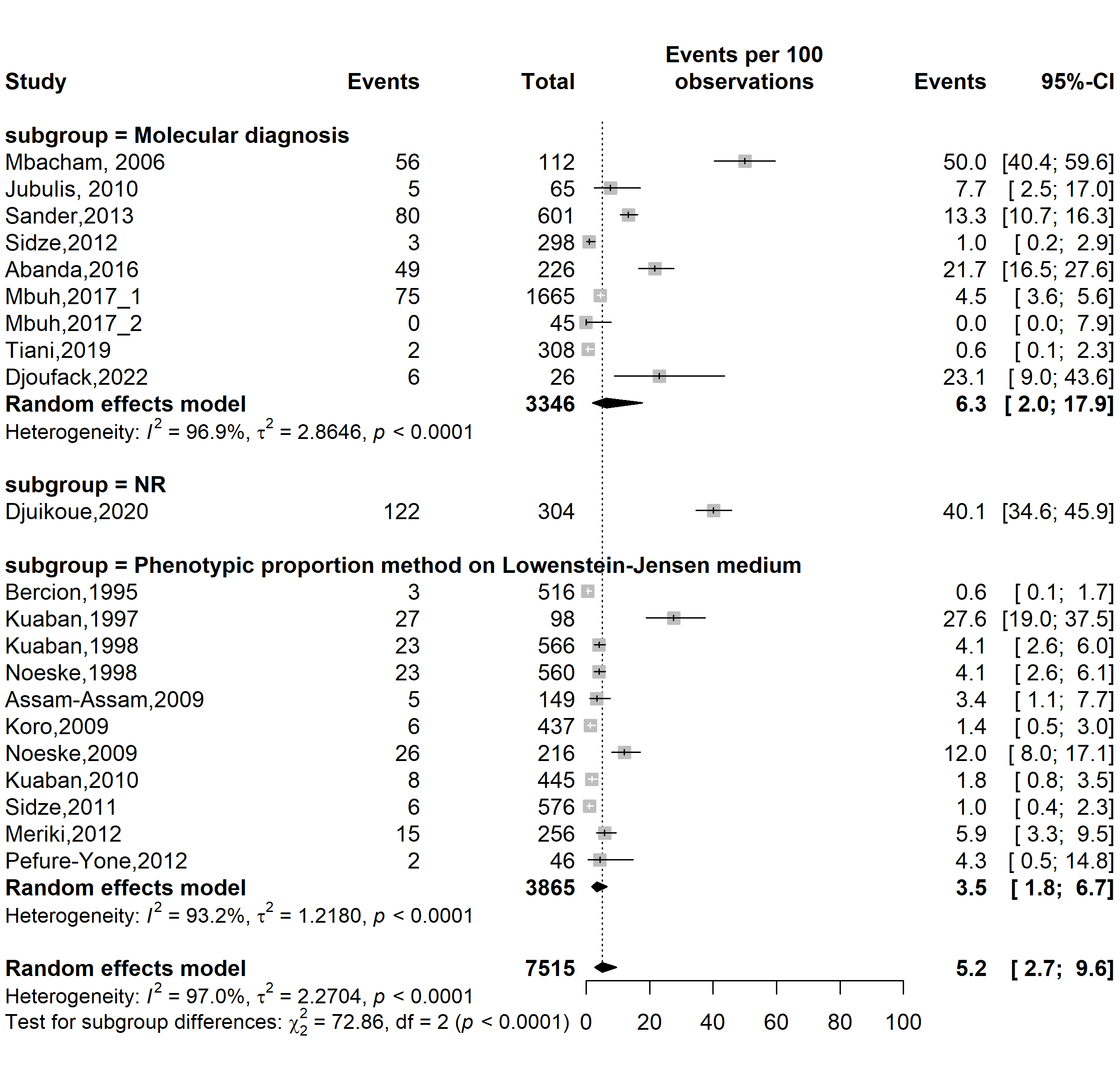

**Fig. S6** Pooled prevalence of multidrug-resistant *Mycobacterium tuberculosis* among tuberculosis patients in Cameroon by type of drug susceptibility testing methods used, 1995–2022

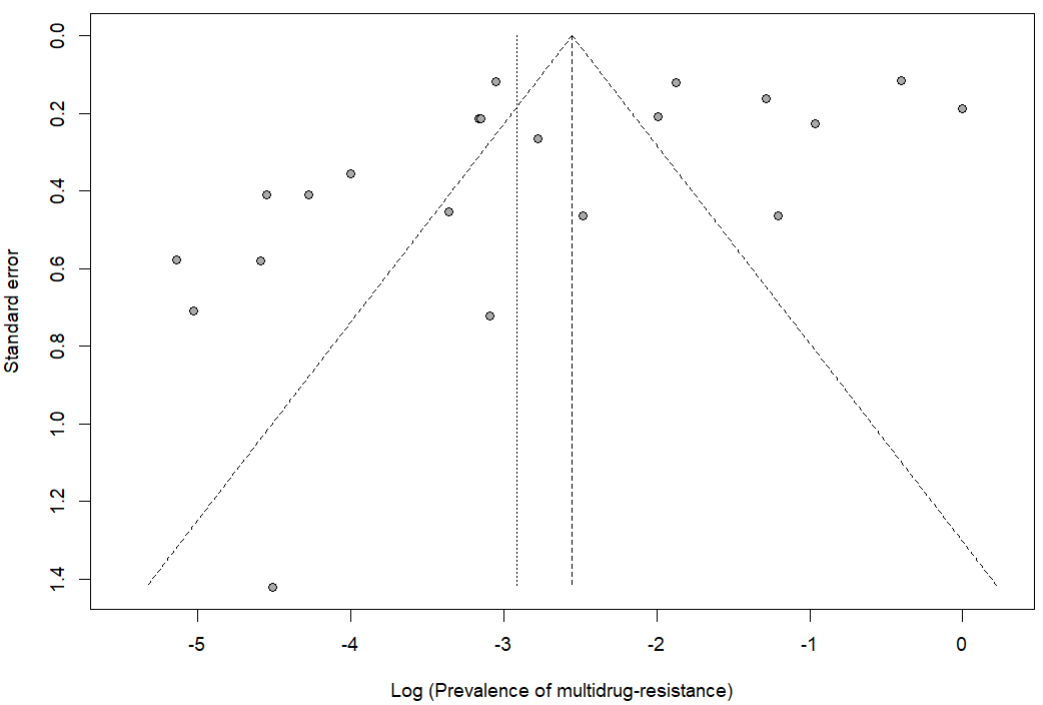

Egger’s test *p*-value = 0.038

Begg’s test *p*-value = 0.566

**Fig. S7** Publication bias of study reports assessing the prevalence of multidrug-resistant *Mycobacterium tuberculosis* among tuberculosis patients in Cameroon, 1995–2022

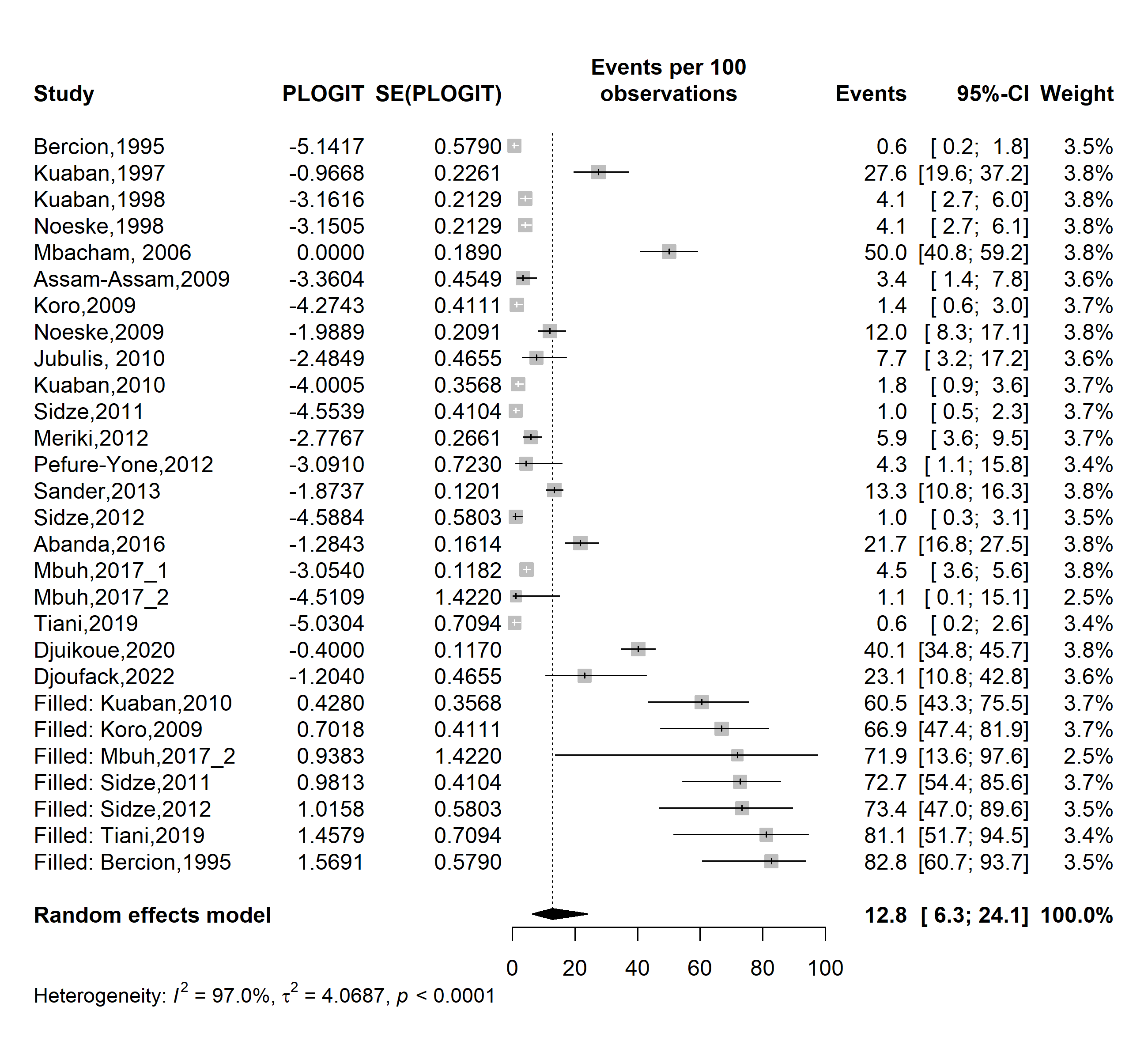

**Fig. S8** Trim and fill analysis adjusting for publication bias among study assessing the pooled prevalence of multidrug-resistant *Mycobacterium tuberculosis* among tuberculosis patients in Cameroon, 1995–2022

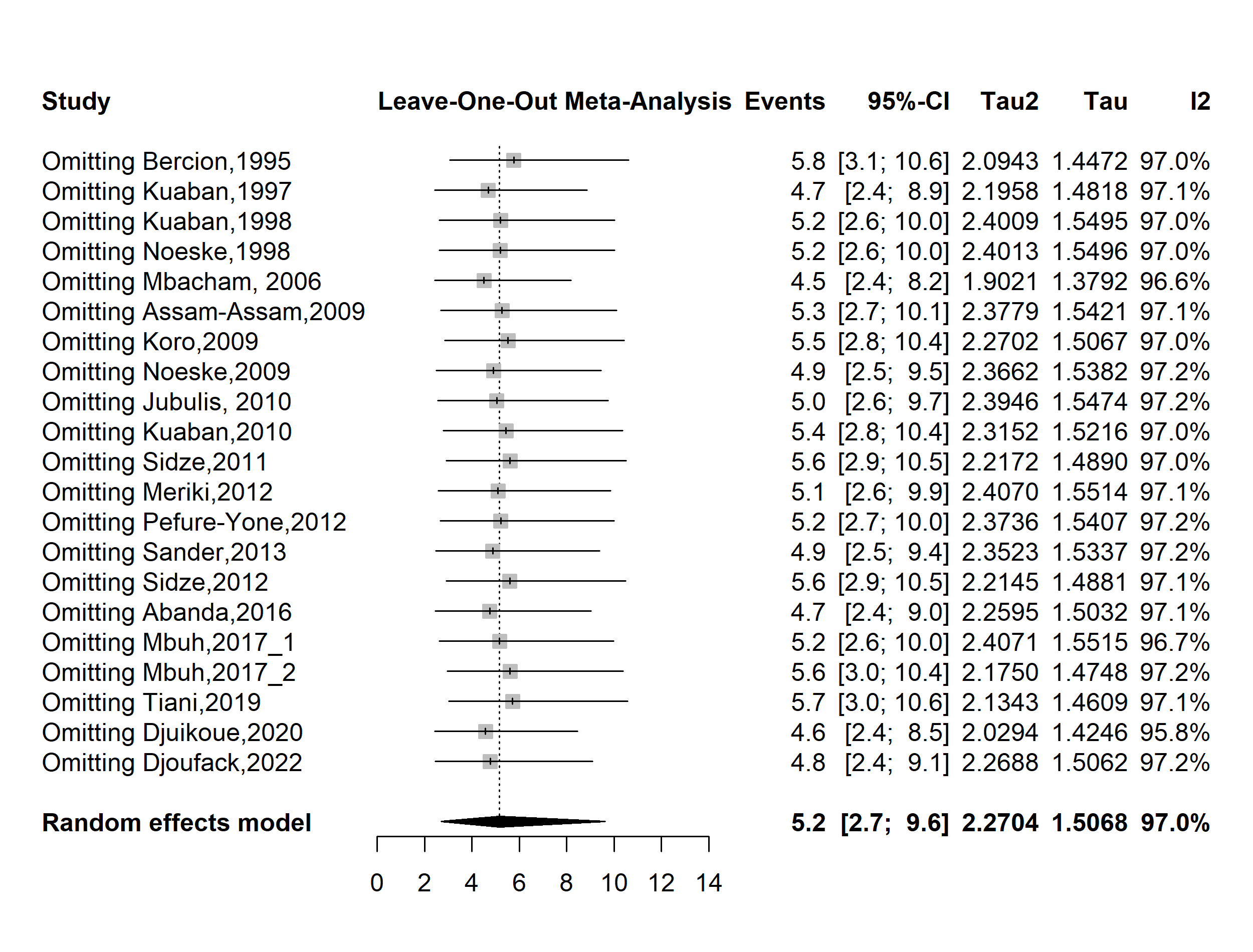

**Fig. S9** Sensitivity analysis of the pooled prevalence of multidrug-resistant *Mycobacterium tuberculosis* among tuberculosis patients in Cameroon, 1995–2022

**Initial multidrug-resistance**

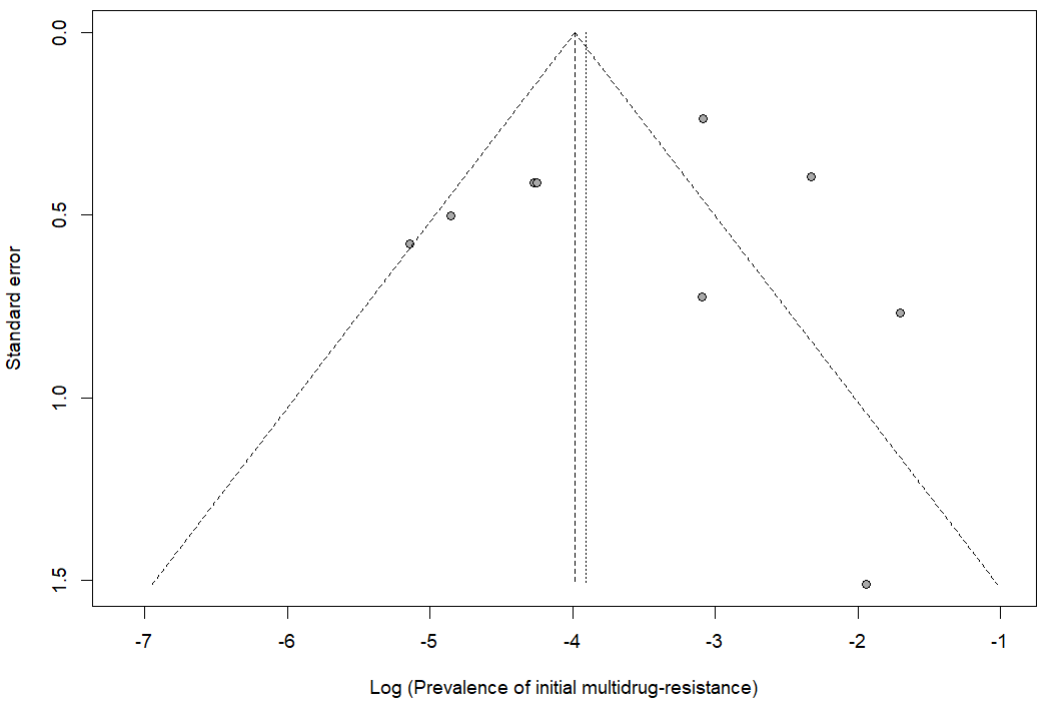

Egger’s test *p*-value = 0.790

Begg’s test *p*-value = 0.876

**Fig. S10** Publication bias of study reports assessing the pooled prevalence of initial multidrug-resistant *Mycobacterium tuberculosis* among new tuberculosis patients in Cameroon, 1995–2022

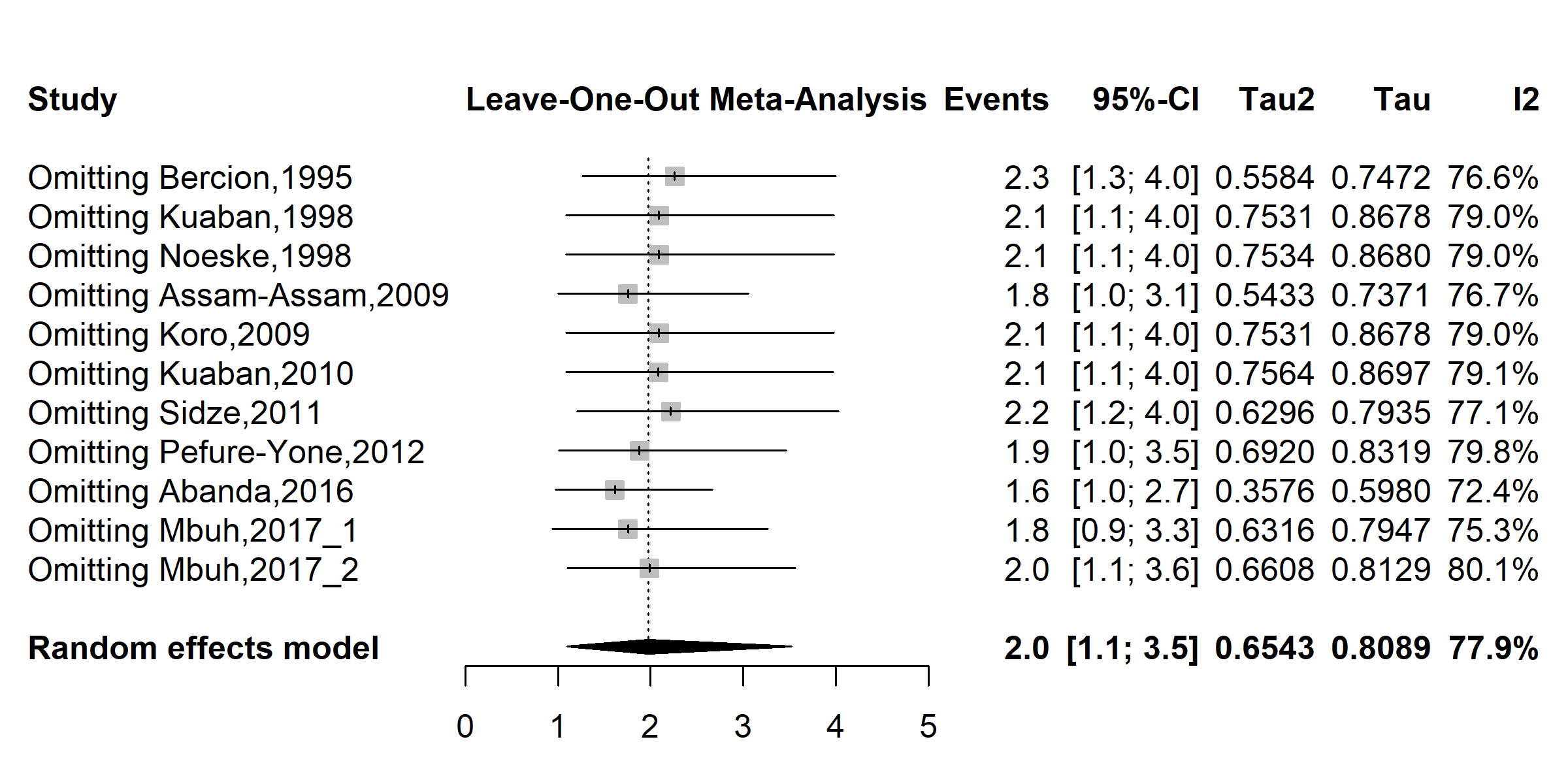

**Fig. S11** Sensitivity analysis of the pooled prevalence of initial multidrug-resistant *Mycobacterium tuberculosis* among new tuberculosis patients in Cameroon, 1995–2022

**Acquired multidrug-resistance**

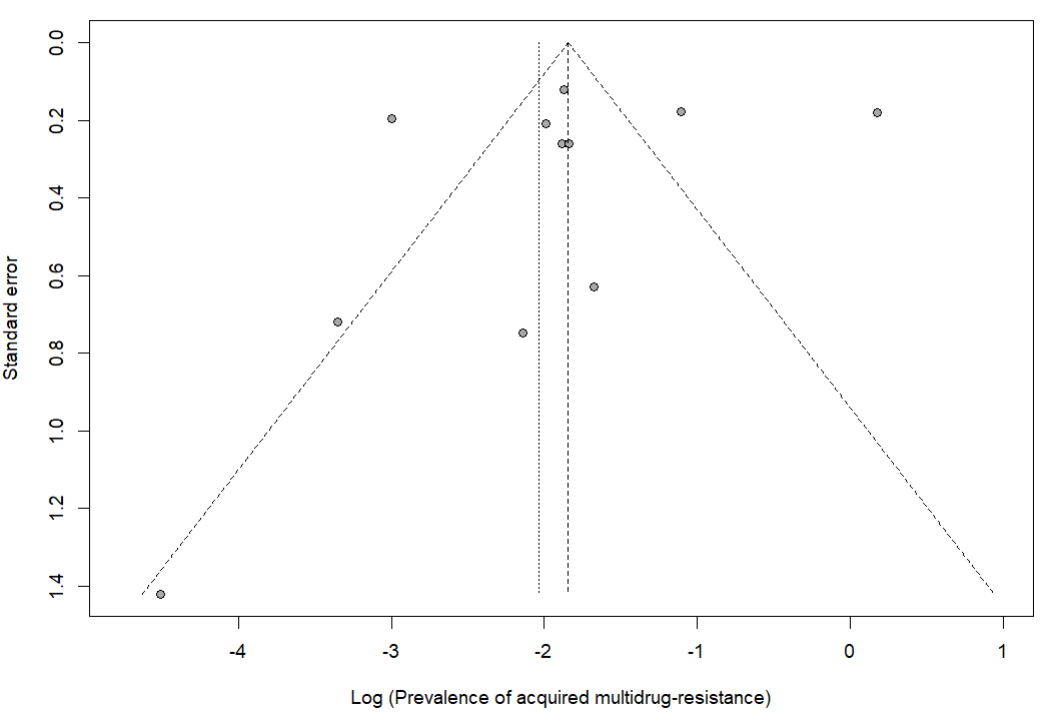

Egger’s test *p*-value = 0.554

Begg’s test *p*-value = 0.938

**Fig. S12** Publication bias of study reports assessing the pooled prevalence of acquired multidrug-resistant *Mycobacterium tuberculosis* among retreated tuberculosis patients in Cameroon, 1995–2022

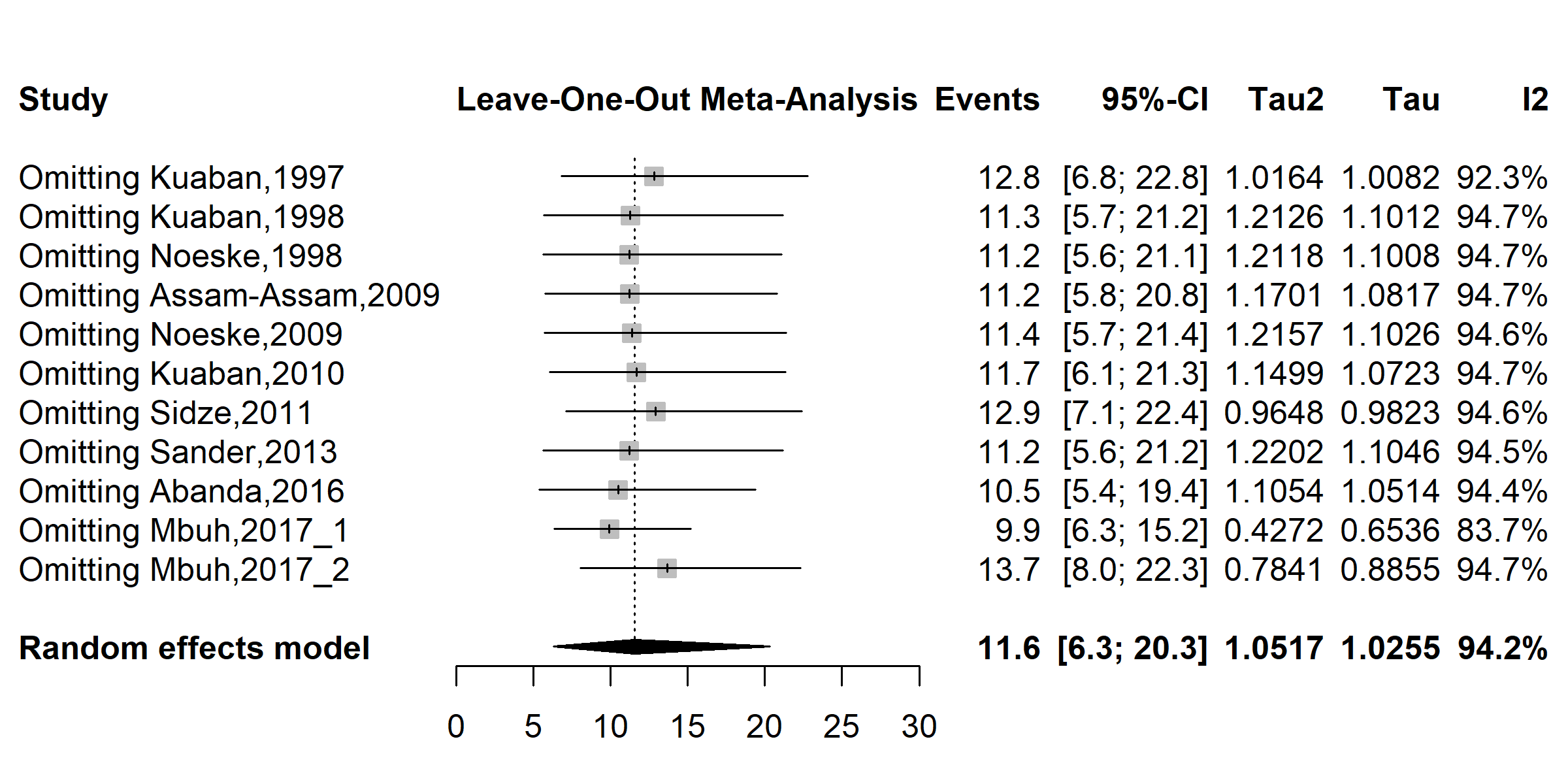

**Fig. S13** Sensitivity analysis of the pooled prevalence of acquired multidrug-resistant *Mycobacterium tuberculosis* among retreated tuberculosis patients in Cameroon, 1995–2022

**Any resistance to anti-TB drugs**

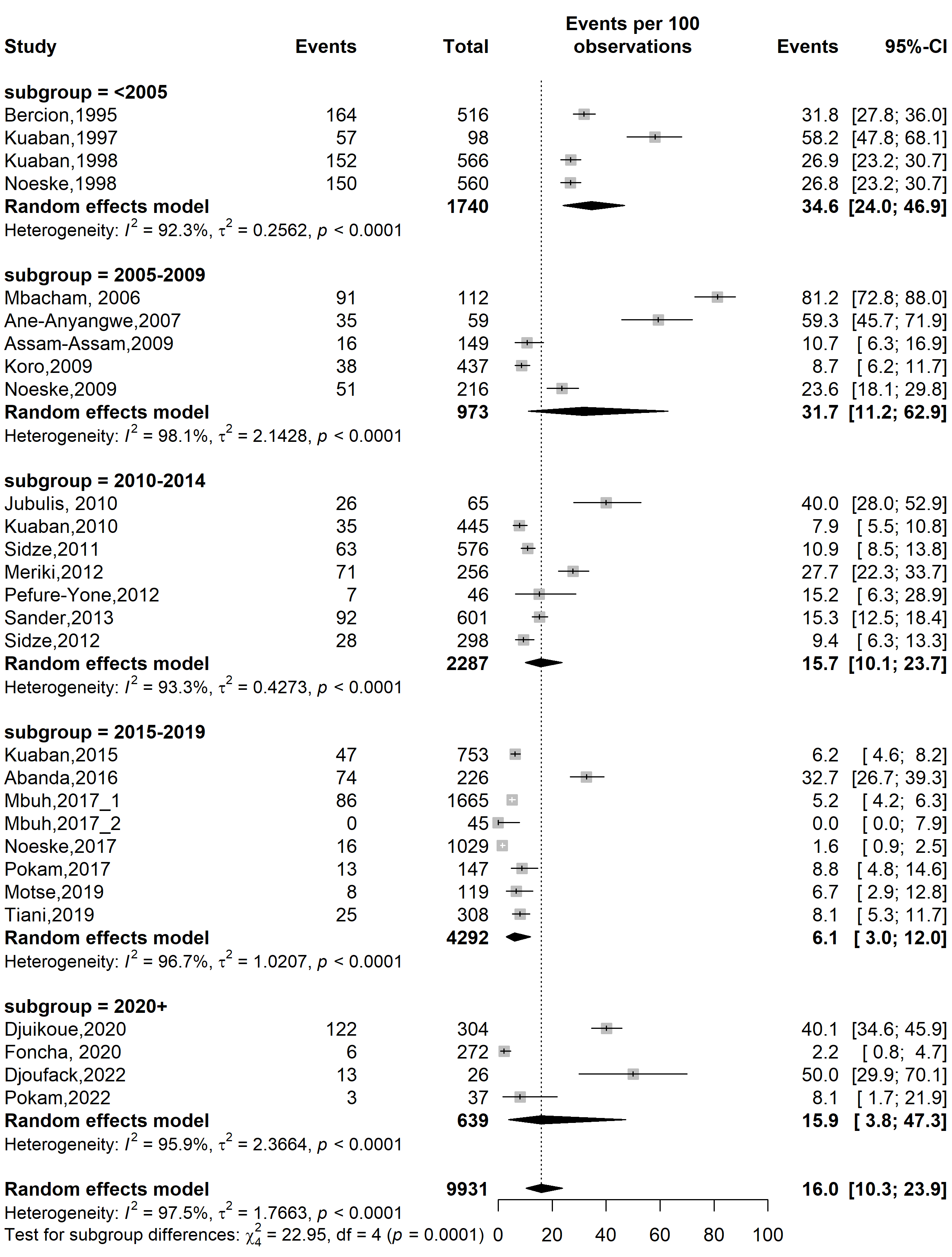

**Fig. S14** Pooled prevalence of any resistance to anti-tuberculosis drugs among tuberculosis patients in Cameroon by study period, 1995–2022

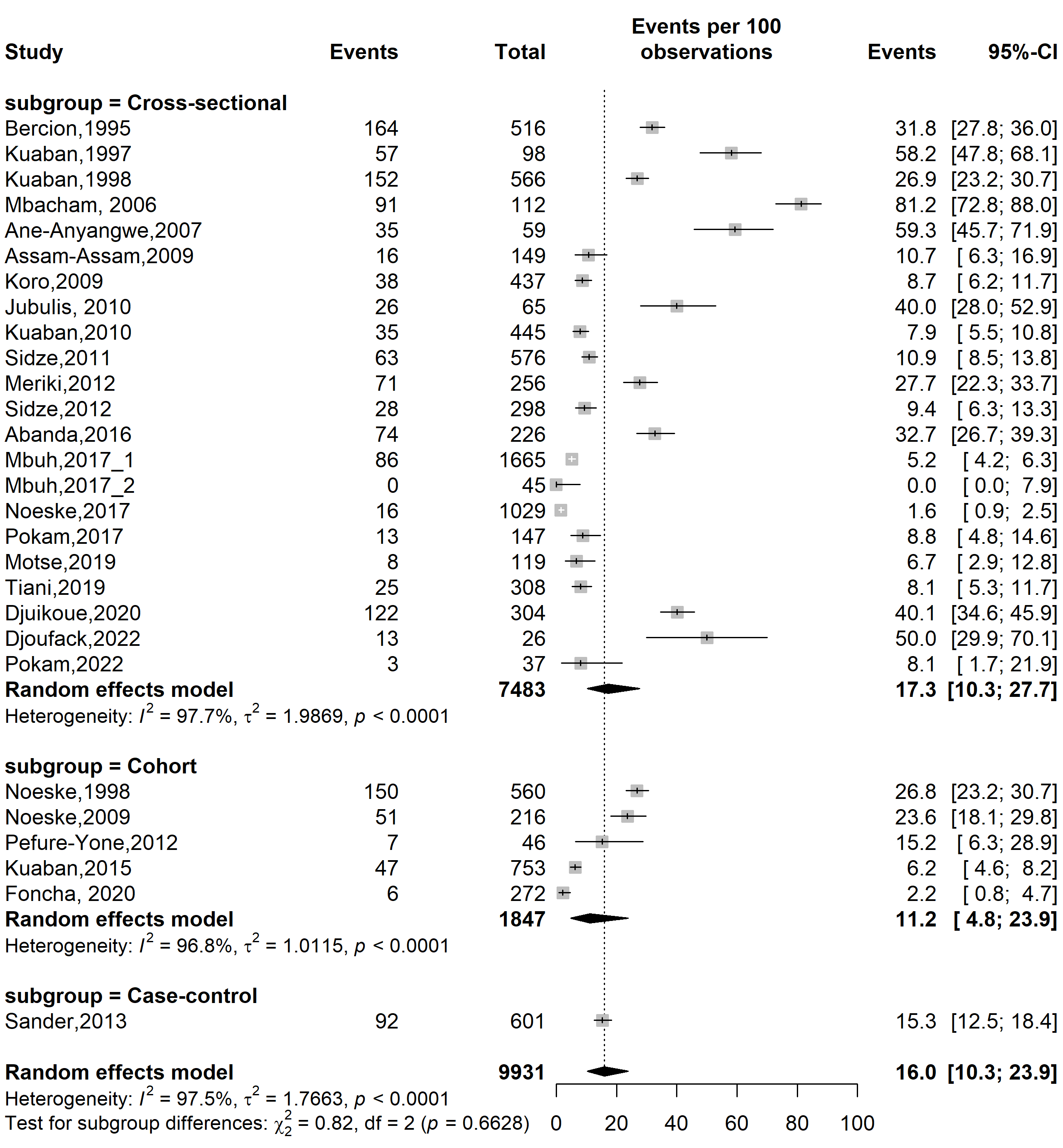

**Fig. S15** Pooled prevalence of any resistance to anti-tuberculosis drugs among tuberculosis patients in Cameroon by study design, 1995–2022

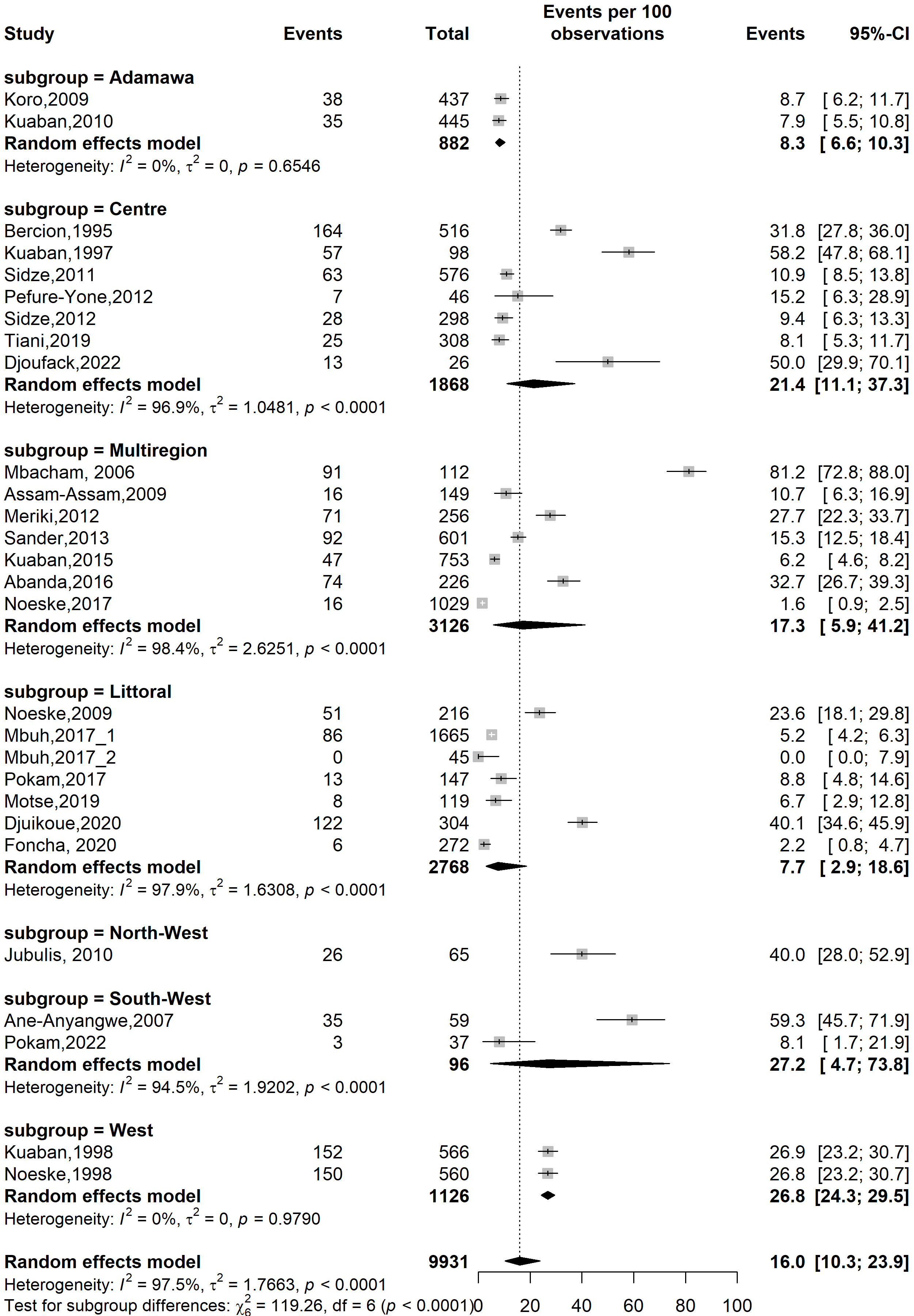

**Fig. S16** Pooled prevalence of any resistance to anti-tuberculosis drugs among tuberculosis patients in Cameroon by study design, 1995–2022

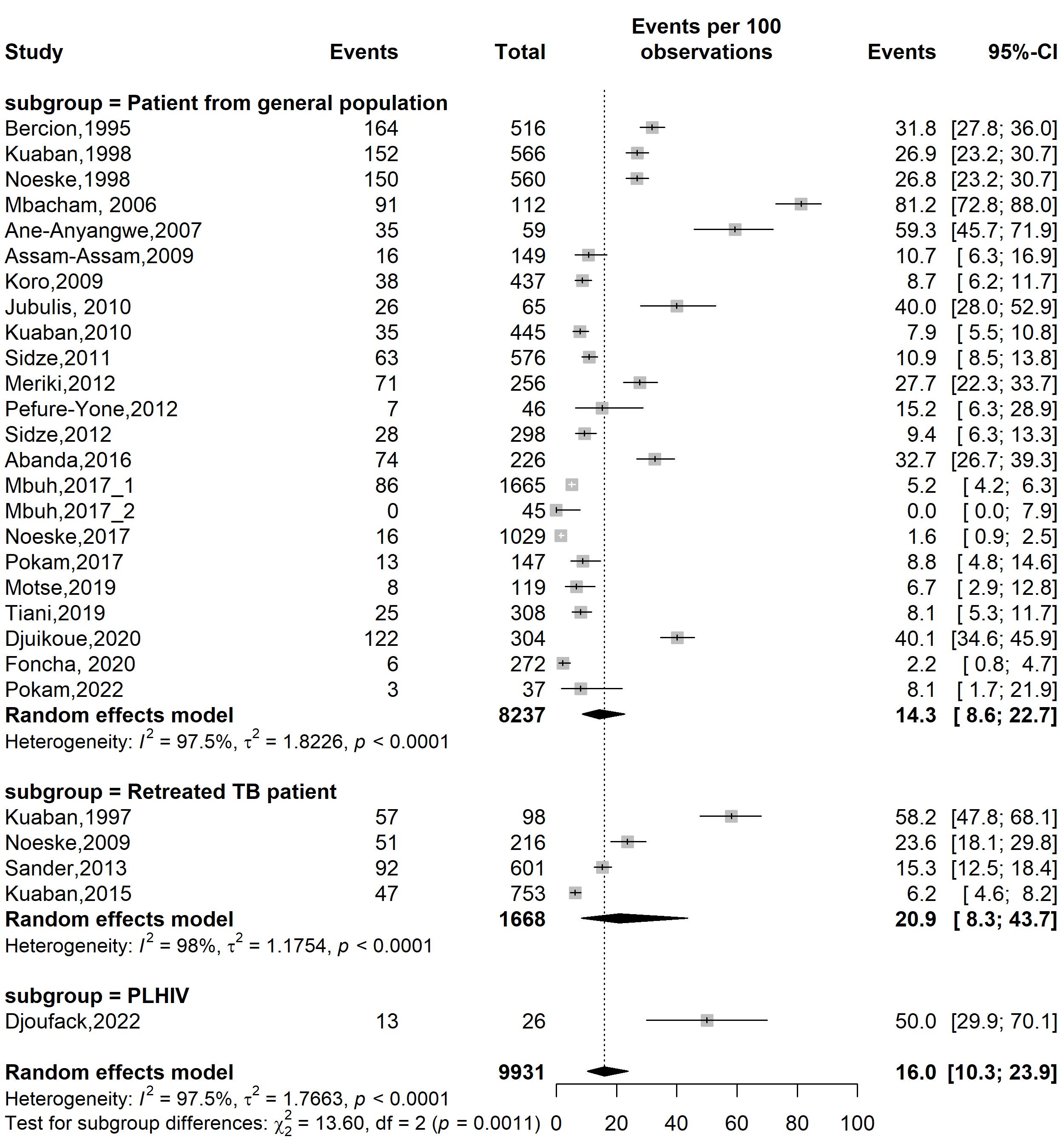

**Fig. S17** Pooled prevalence of any resistance to anti-tuberculosis drugs among tuberculosis patients in Cameroon by type of participants, 1995–2022

(*TB: Tuberculosis; PLHIV: People living with human immunodeficiency virus*)

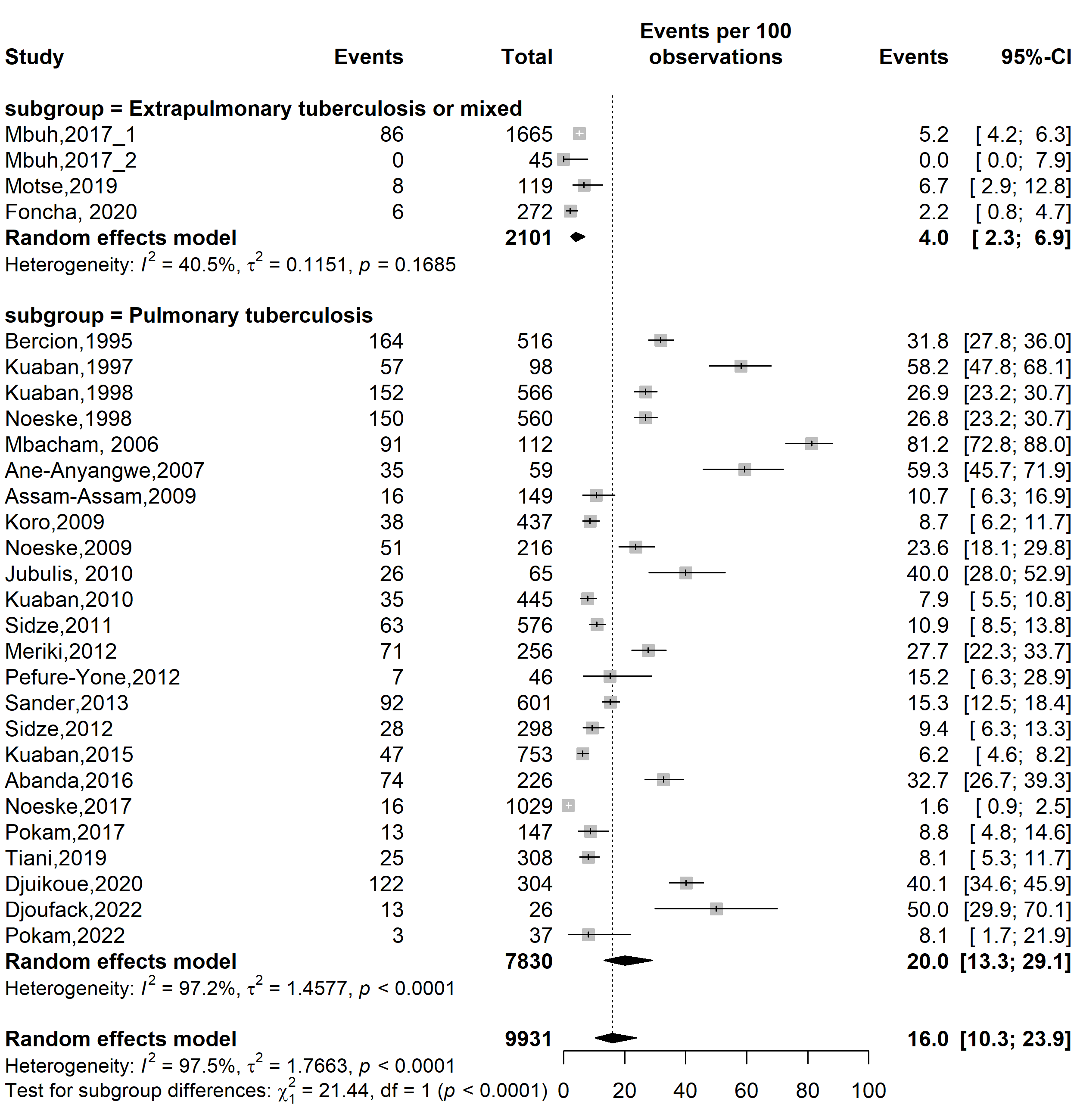

**Fig. S18** Pooled prevalence of any resistance to anti-tuberculosis drugs among tuberculosis patients in Cameroon by type of tuberculosis infection localizations, 1995–2022

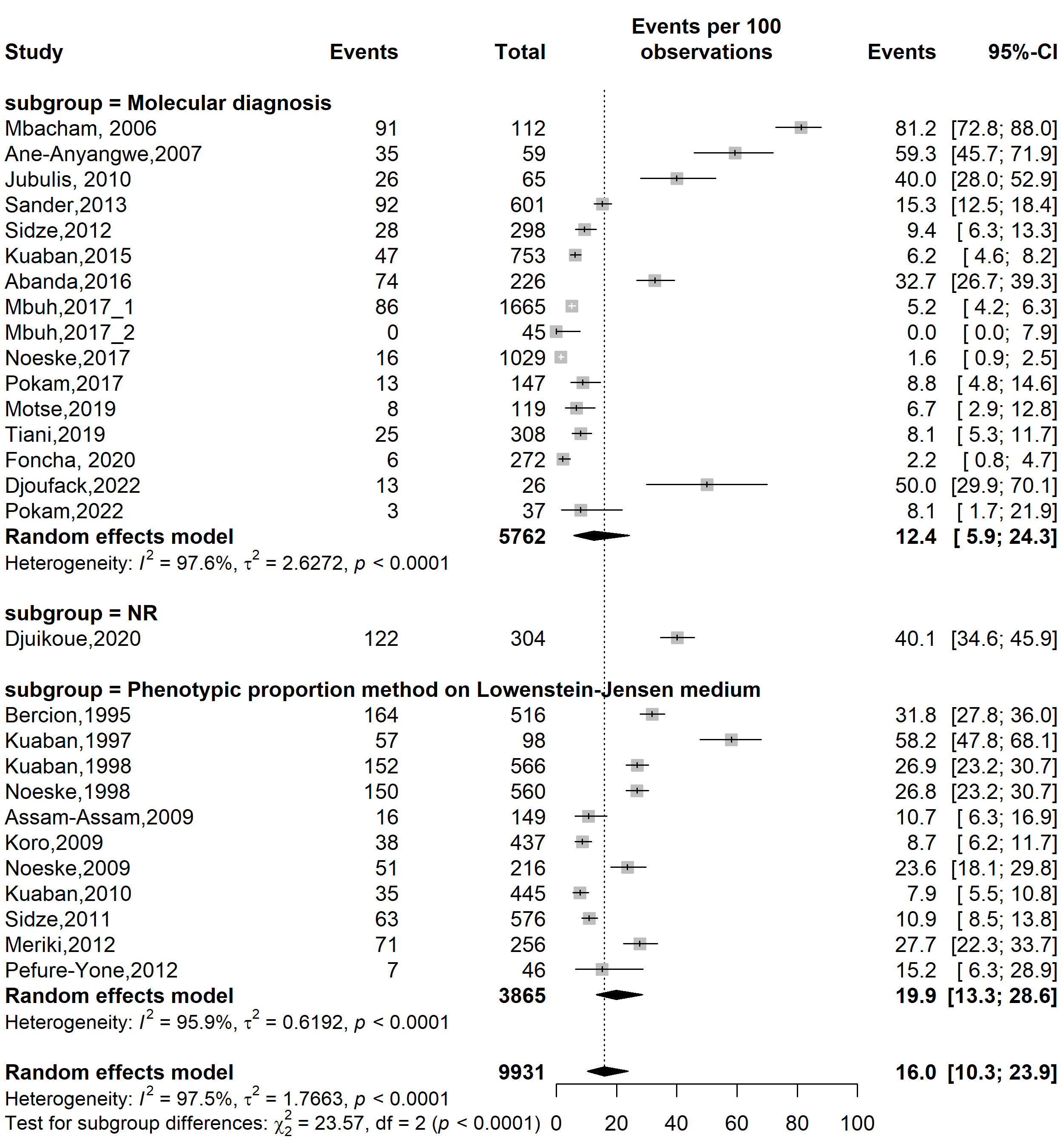

**Fig. S19** Pooled prevalence of any resistance to anti-tuberculosis drugs among tuberculosis patients in Cameroon by type of drug susceptibility testing methods used, 1995–2022

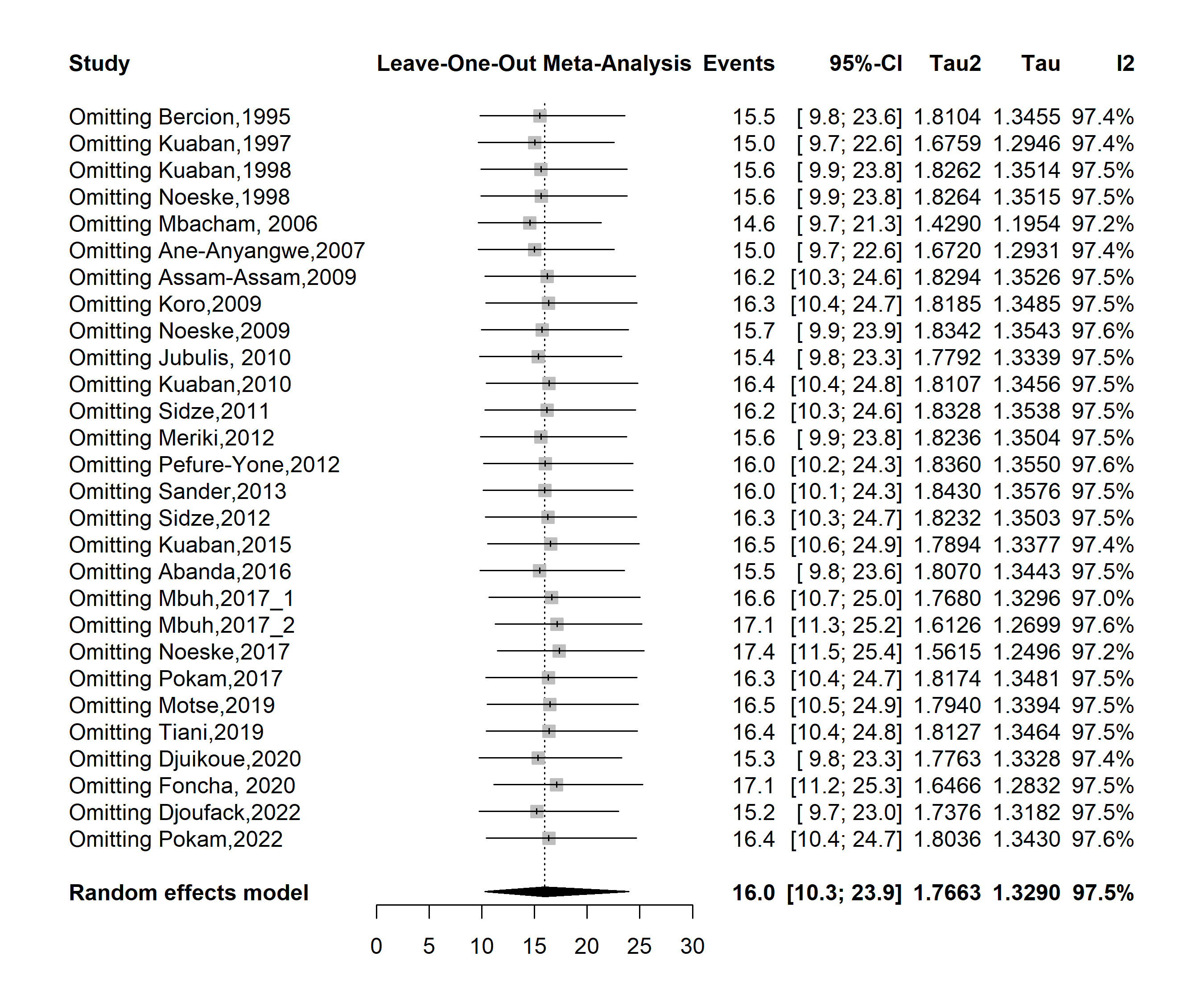

**Fig. S20** Sensitivity analysis of the pooled prevalence of any resistance to anti-tuberculosis drugs among tuberculosis patients in Cameroon, 1995–2022

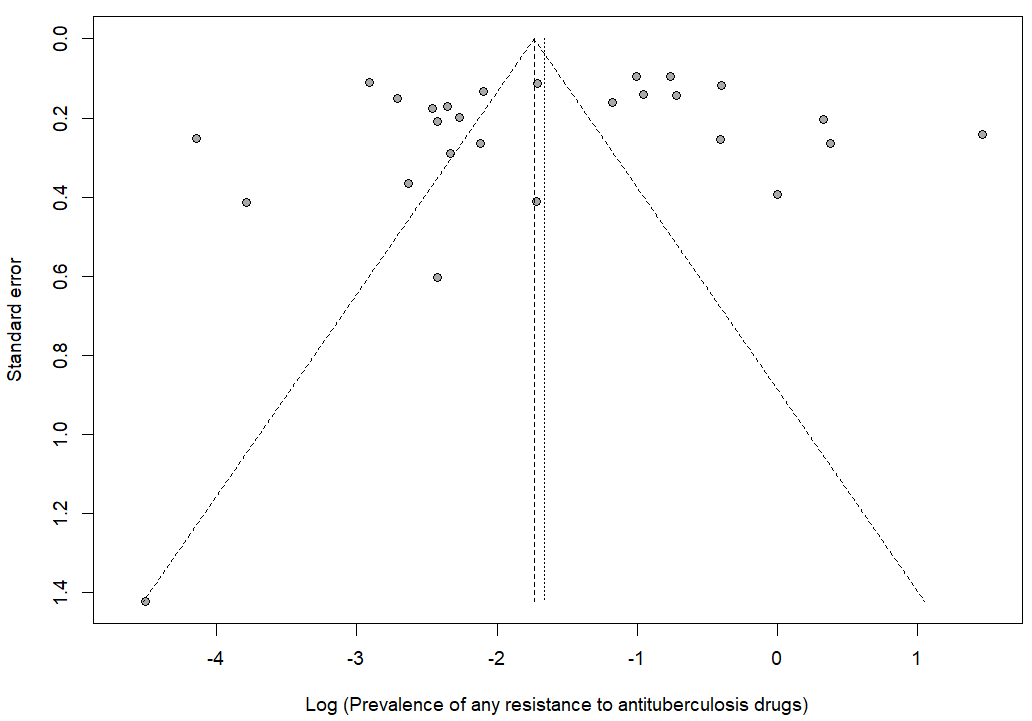

Egger’s test *p*-value = 0.414

Begg’s test *p*-value = 0.527

**Fig. S21** Sensitivity analysis of the pooled prevalence of any resistance to anti-tuberculosis drugs among tuberculosis patients in Cameroon, 1995–2022

**Any resistance to rifampicin**

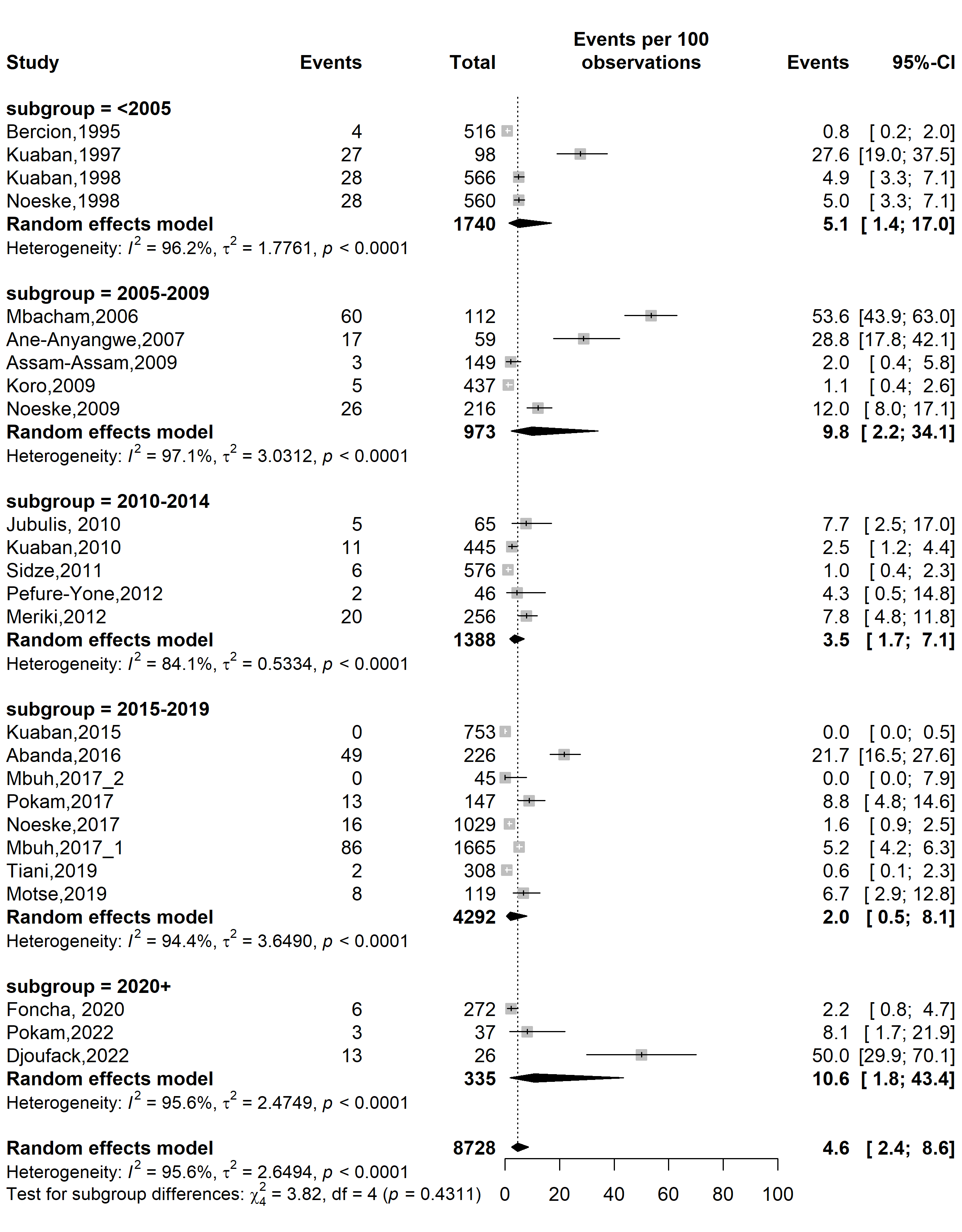

**Fig. S22** Pooled prevalence of any resistance to rifampicin among tuberculosis patients in Cameroon by study period, 1995–2022

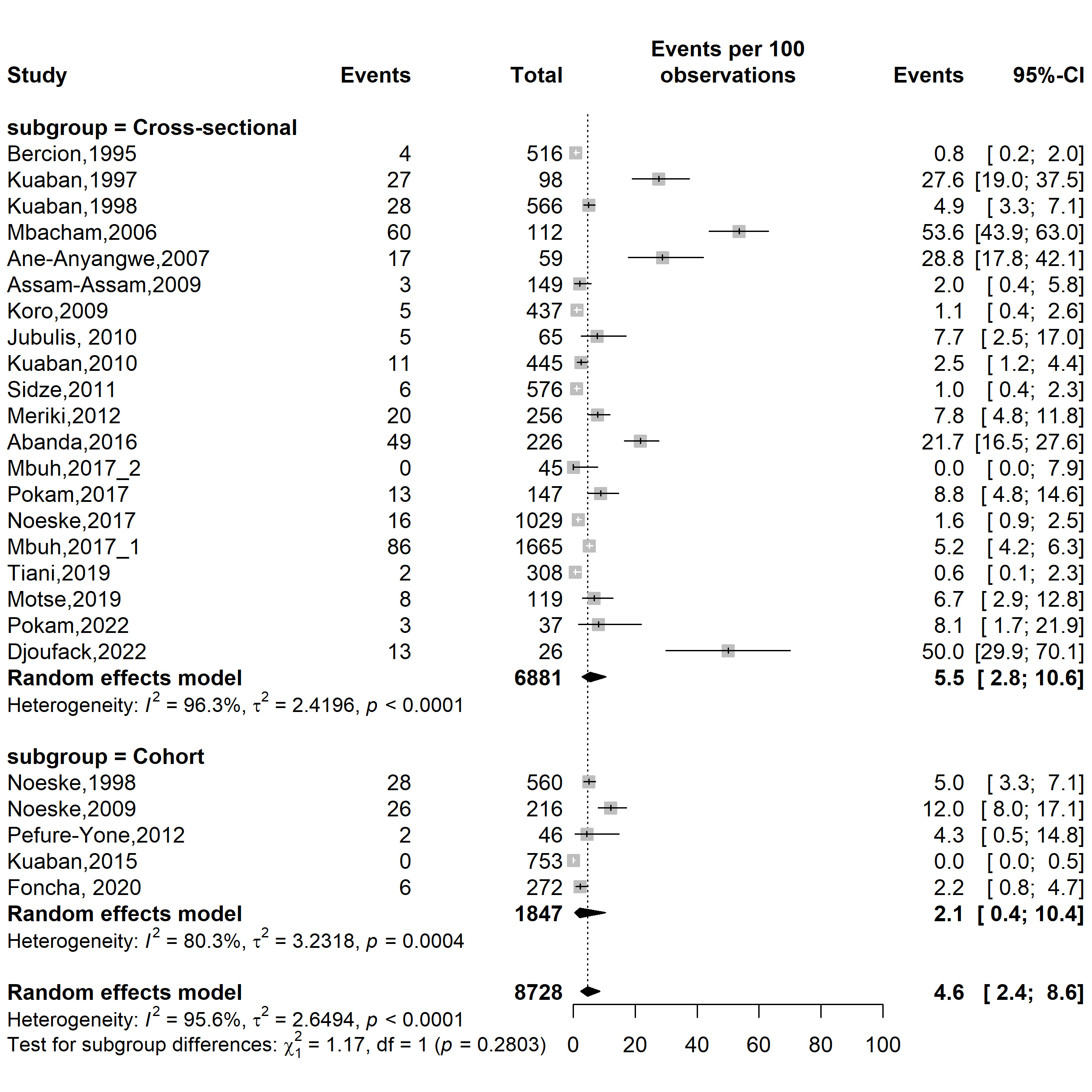

**Fig. S23** Pooled prevalence of any resistance to rifampicin among tuberculosis patients in Cameroon by study design, 1995–2022

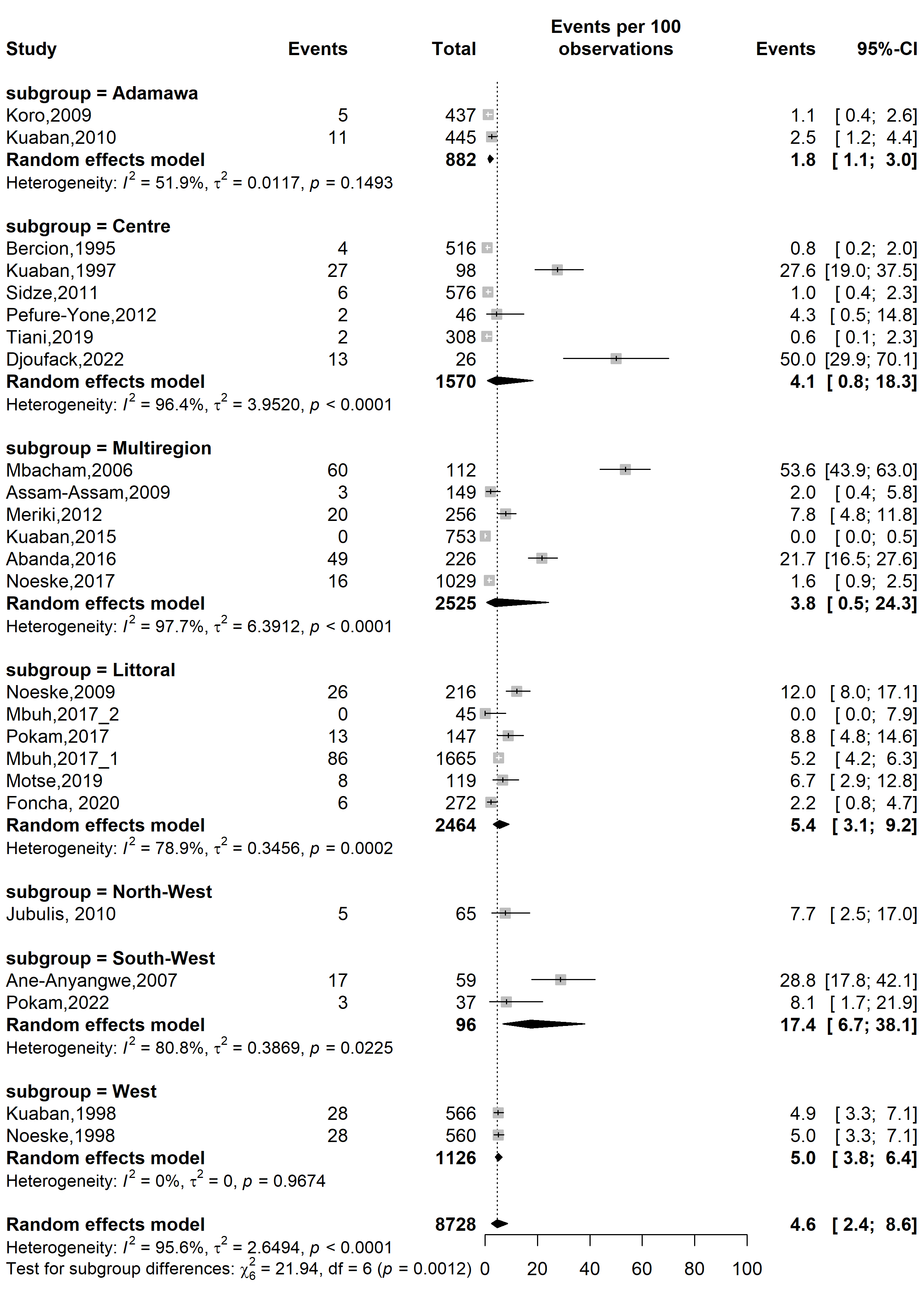

**Fig. S24** Pooled prevalence of any resistance to rifampicin among tuberculosis patients in Cameroon by region, 1995–2022

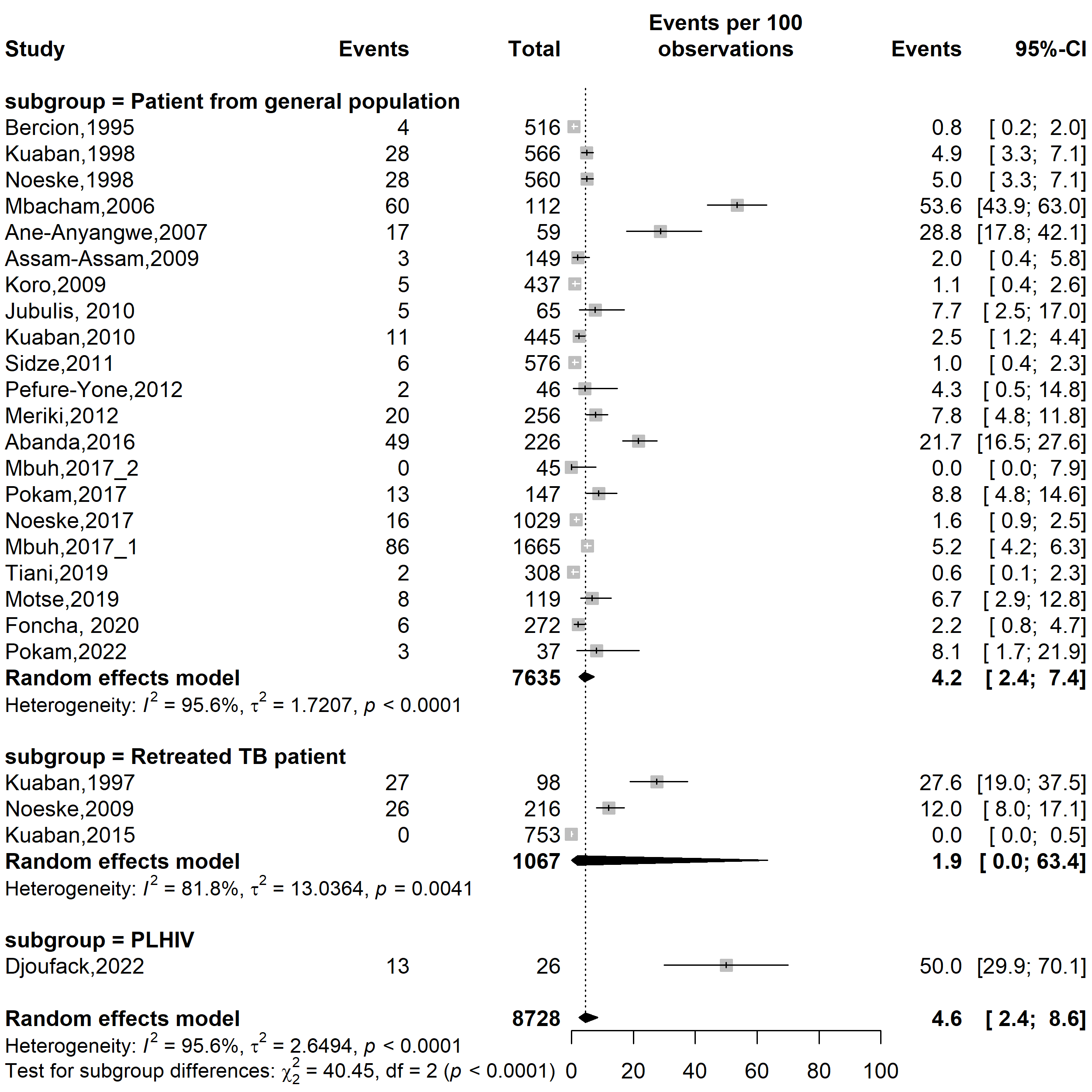

**Fig. S25** Pooled prevalence of any resistance to rifampicin among tuberculosis patients in Cameroon by type of participants, 1995–2022

(*TB: Tuberculosis; PLHIV: People living with human immunodeficiency virus*)

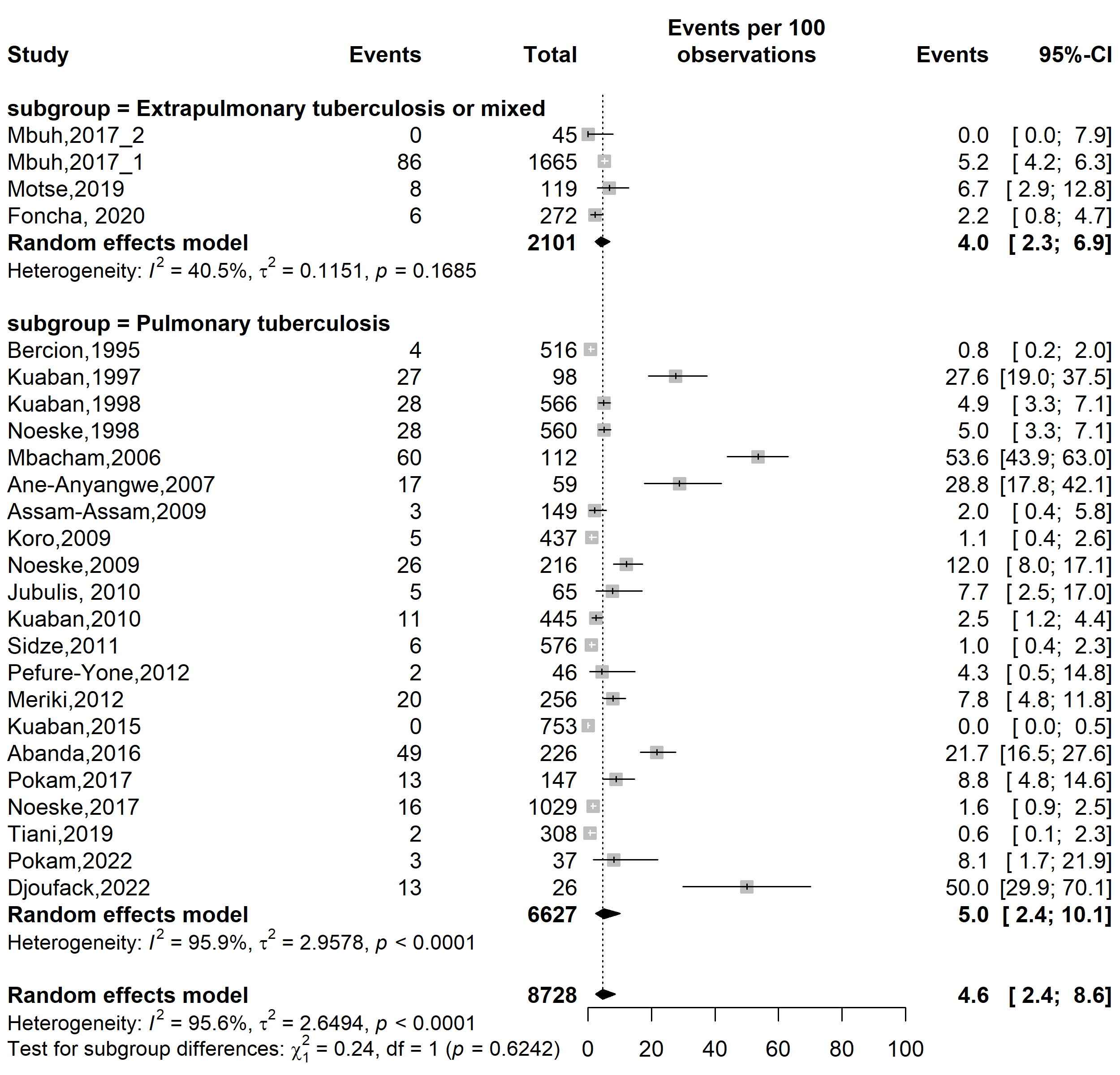

**Fig. S26** Pooled prevalence of any resistance to rifampicin among tuberculosis patients in Cameroon by type of tuberculosis infection localizations, 1995–2022

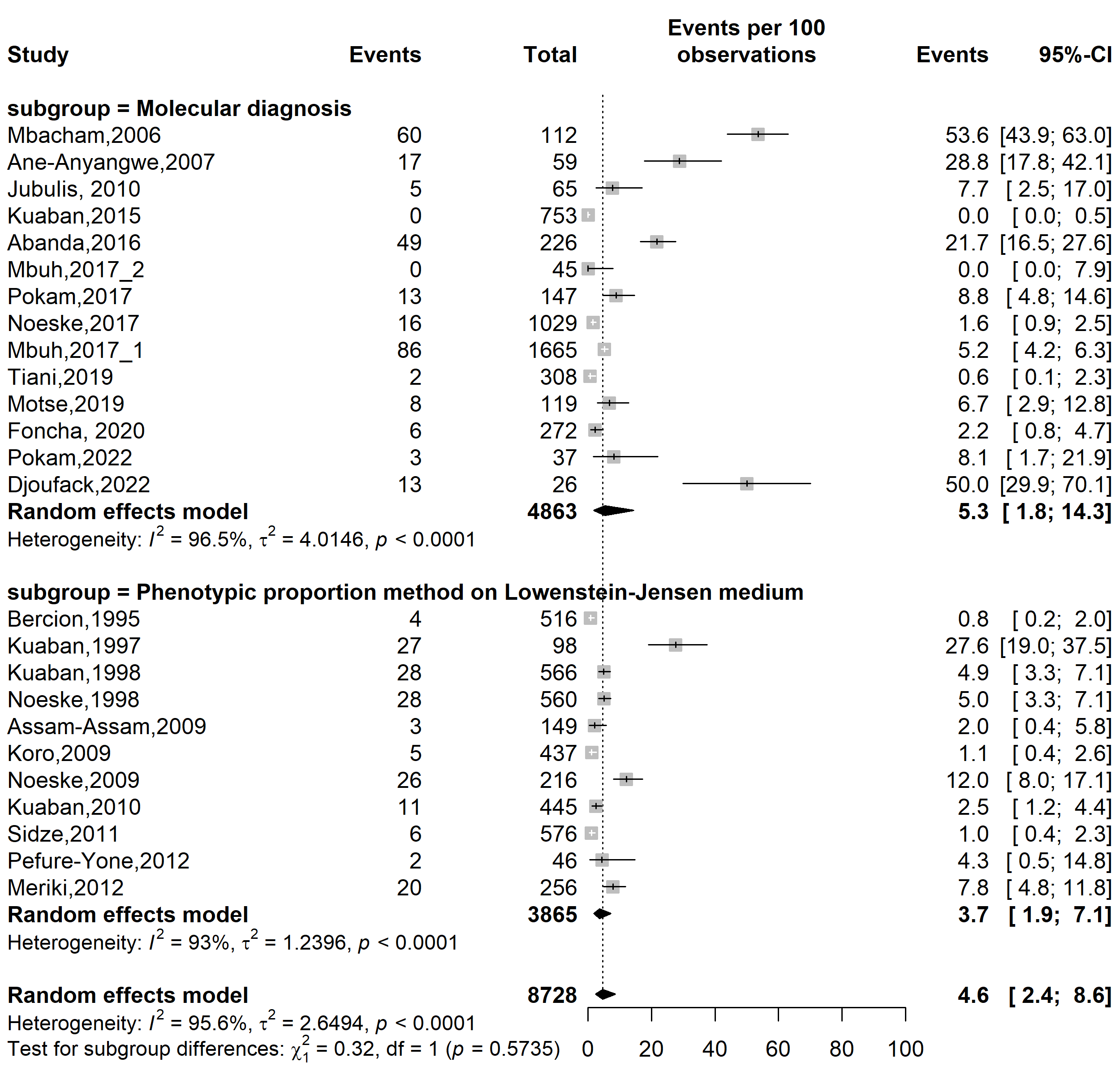

**Fig. S27** Pooled prevalence of any resistance to rifampicin among tuberculosis patients in Cameroon by type of drug susceptibility testing methods used, 1995–2022

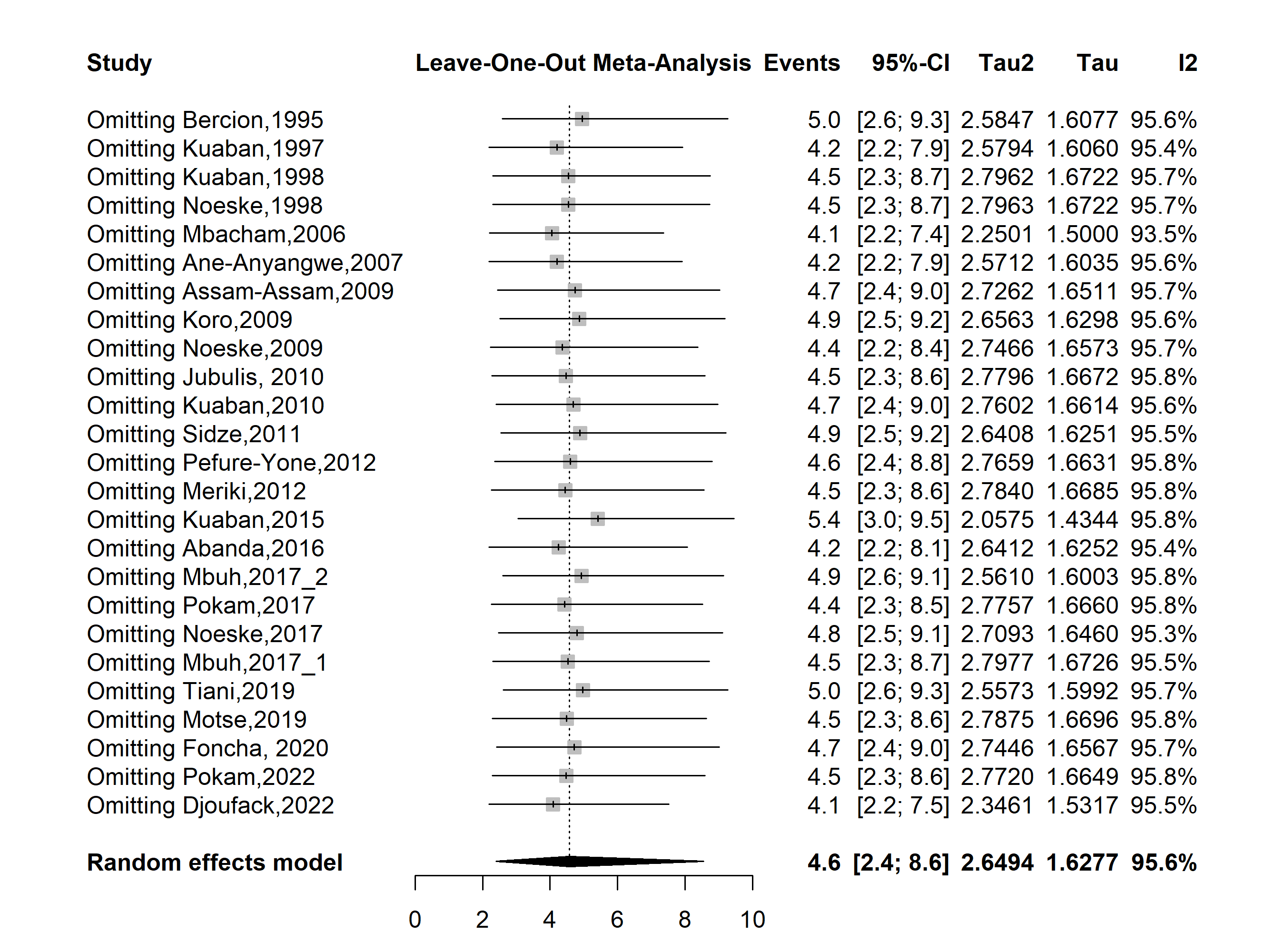

**Fig. S28** Sensitivity analysis of the pooled prevalence of any resistance to rifampicin among tuberculosis patients in Cameroon, 1995–2022

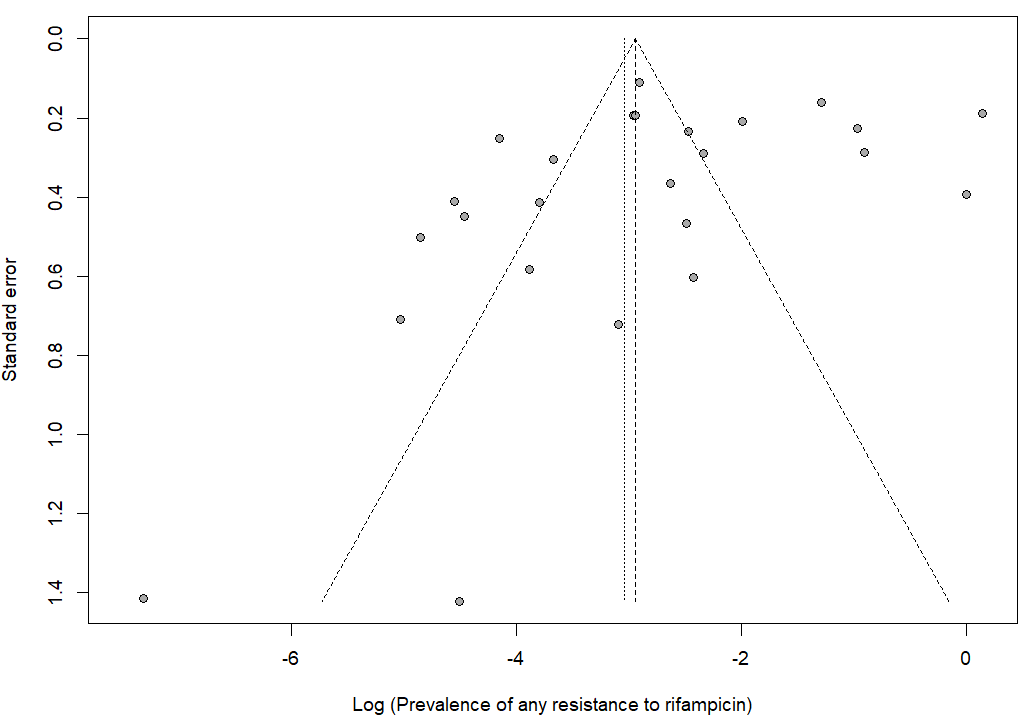

Egger’s test *p*-value = 0.189

Begg’s test *p*-value = 0.225

**Fig. S29** Publication bias of study reports assessing the pooled prevalence of any resistance to rifampicin among tuberculosis patients in Cameroon, 1995–2022

**Monoresistance to rifampicin**

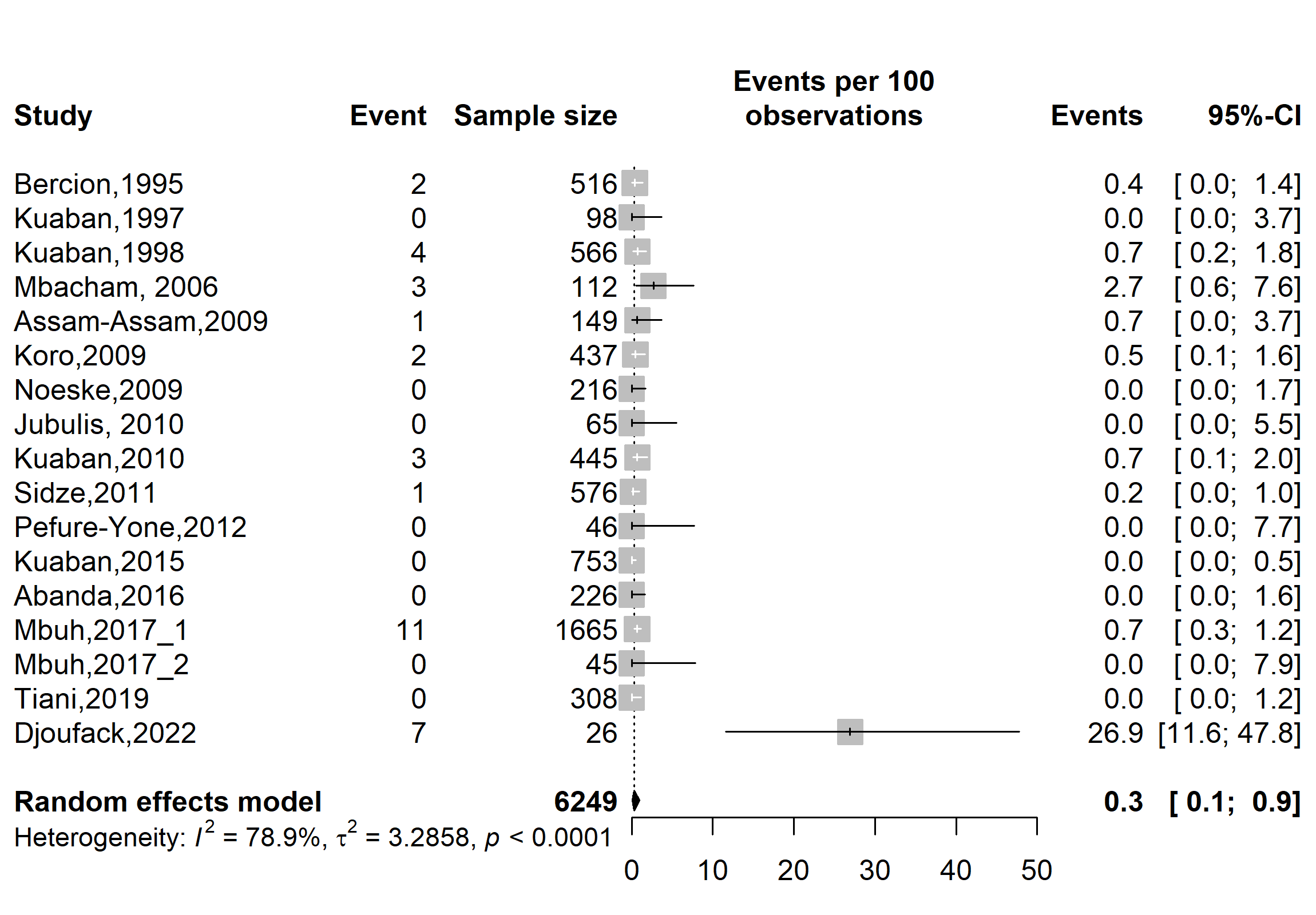

**Fig. S30** Pooled prevalence of rifampicin monoresistance among tuberculosis patients in Cameroon, 1995–2022

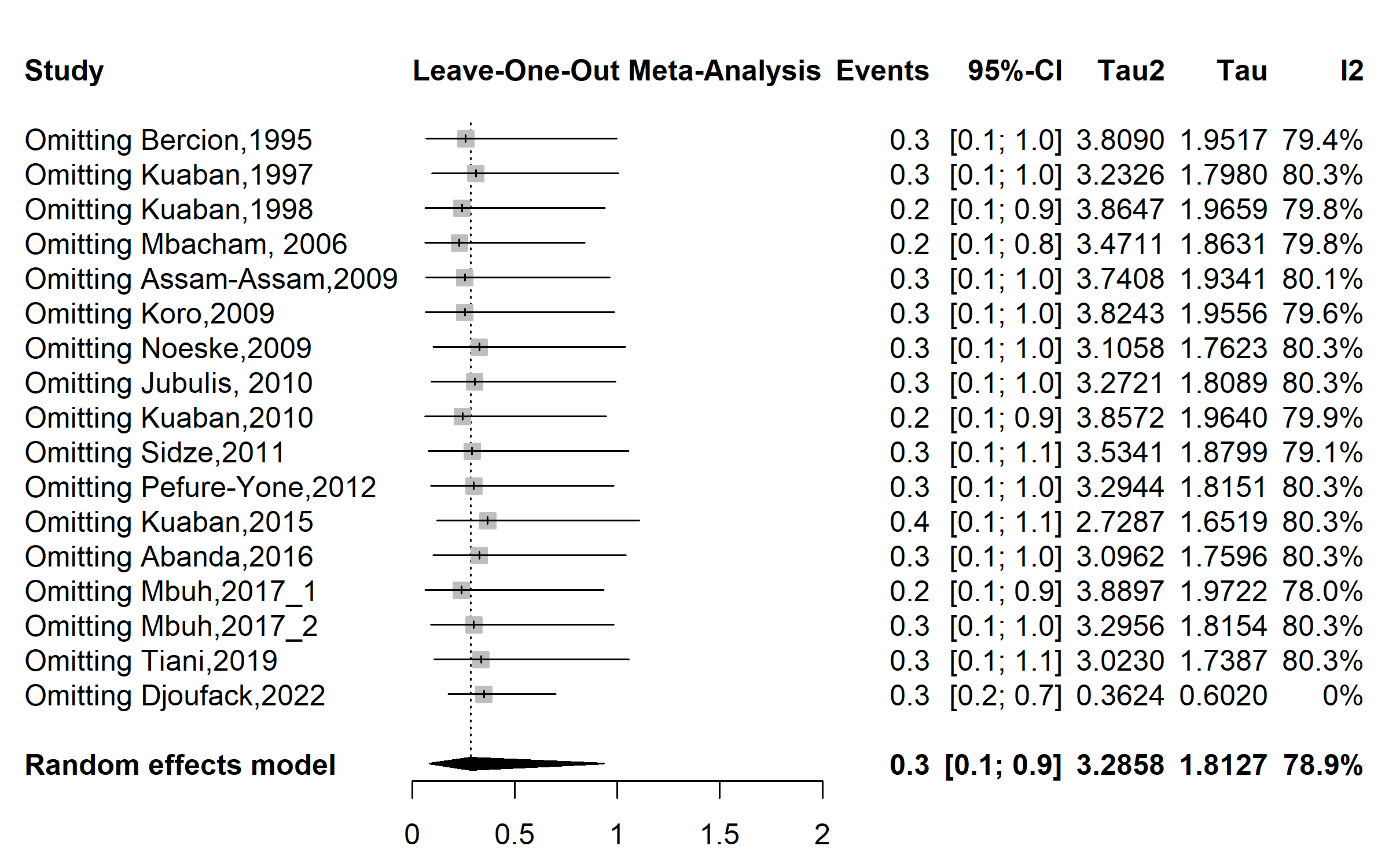

**Fig. S31** Sensitivity analysis of the pooled prevalence of rifampicin monoresistance among tuberculosis patients in Cameroon, 1995–2022

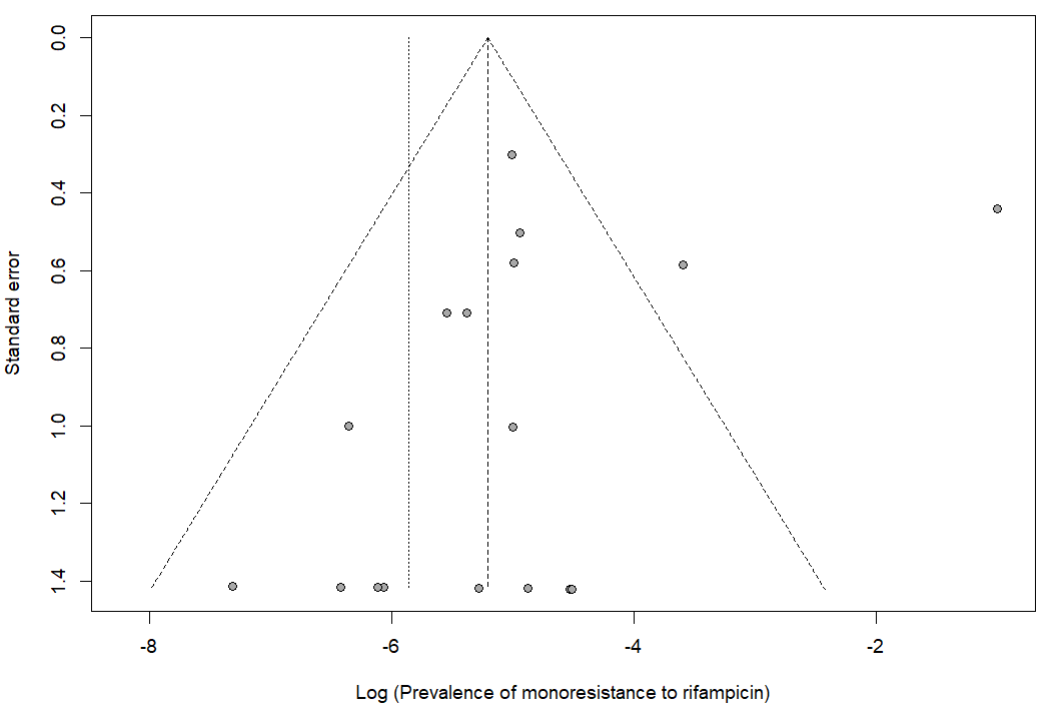

Egger’s test *p*-value = 0.209

Begg’s test *p*-value = 0.187

**Fig. S32** Publication bias of study reports assessing the pooled prevalence of rifampicin monoresistance among tuberculosis patients in Cameroon, 1995–2022

**Monoresistance to isoniazid**

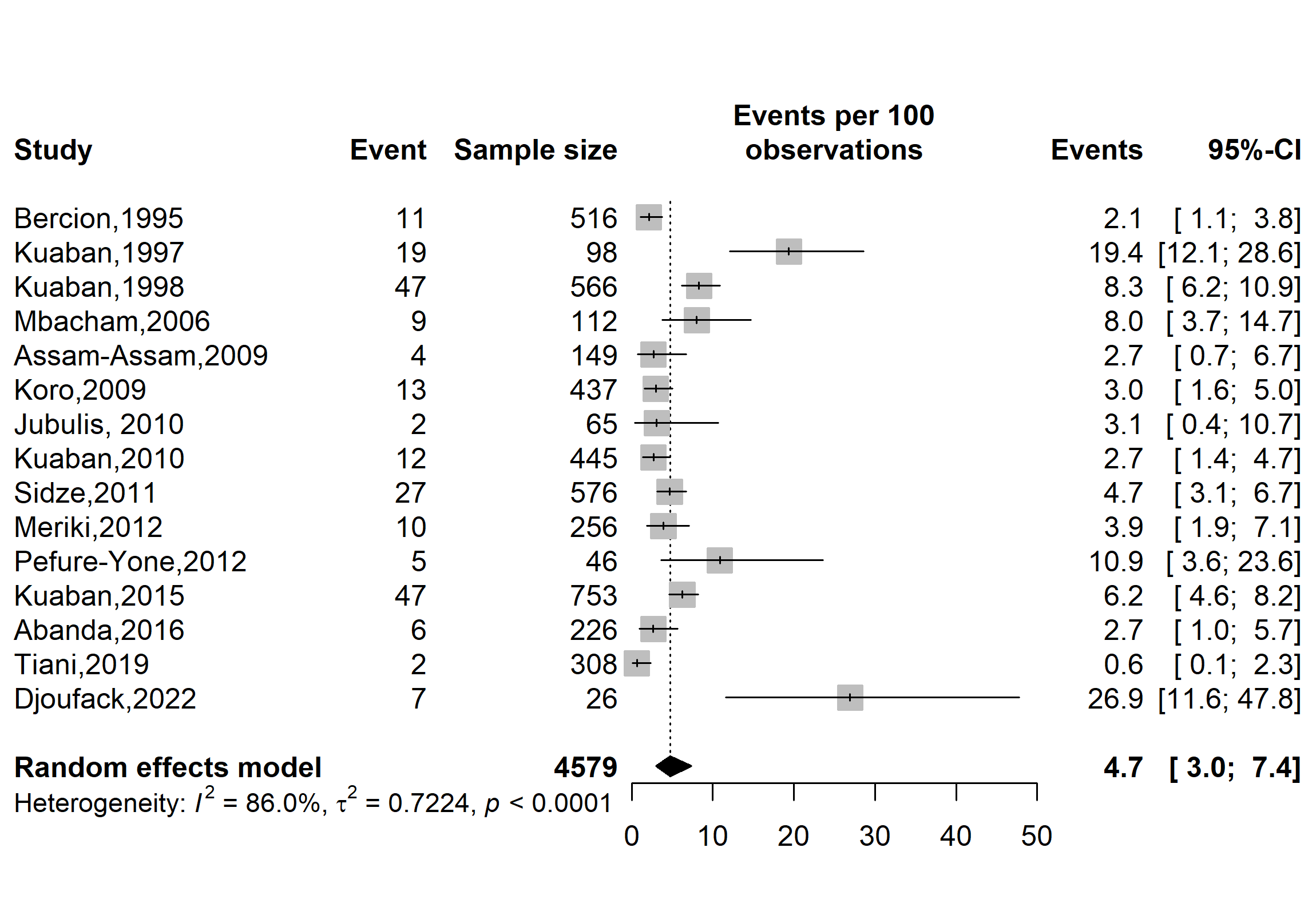

**Fig. S33** Pooled prevalence of isoniazid monoresistance among tuberculosis patients in Cameroon, 1995–2022

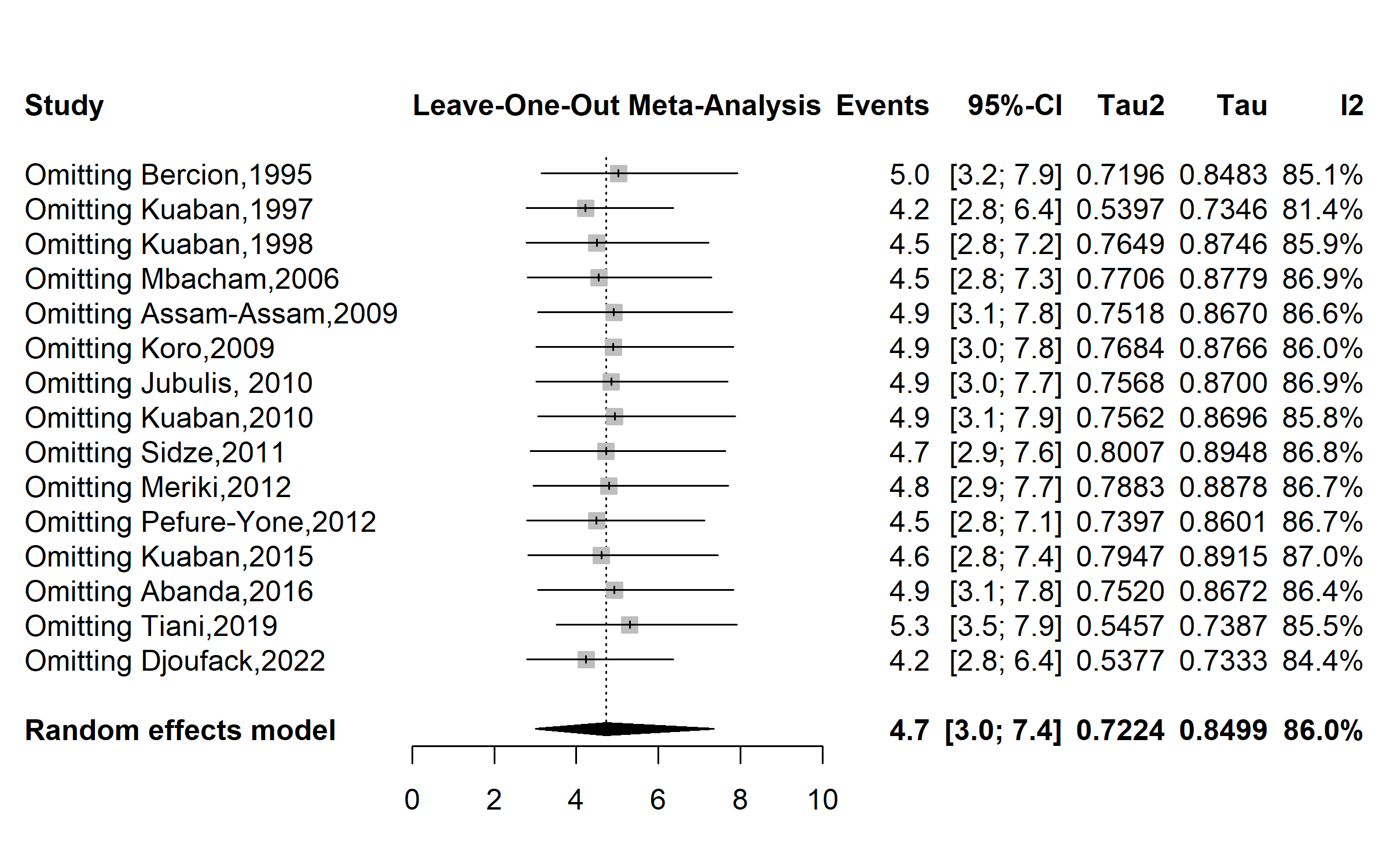

**Fig. S34** Sensitivity analysis of the pooled prevalence of isoniazid monoresistance among tuberculosis patients in Cameroon, 1995–2022

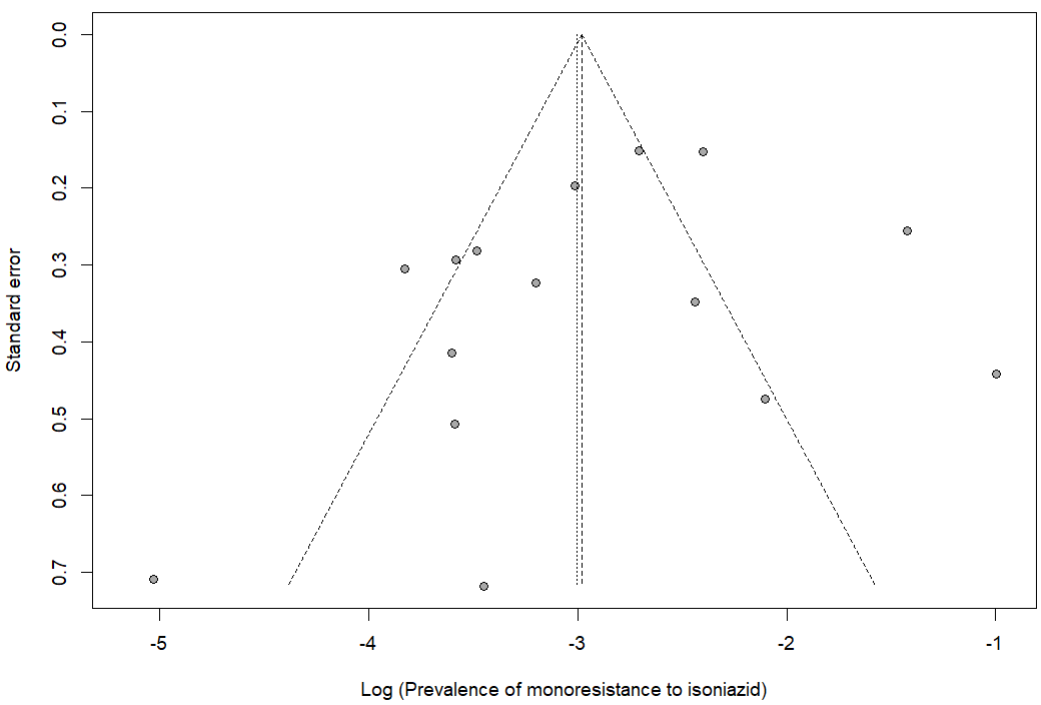

Egger’s test *p*-value = 0.318

Begg’s test *p*-value = 0.520

**Fig. S35** Publication bias of study reports assessing the pooled prevalence of isoniazid monoresistance among tuberculosis patients in Cameroon, 1995–2022

**Monoresistance to ethambutol**

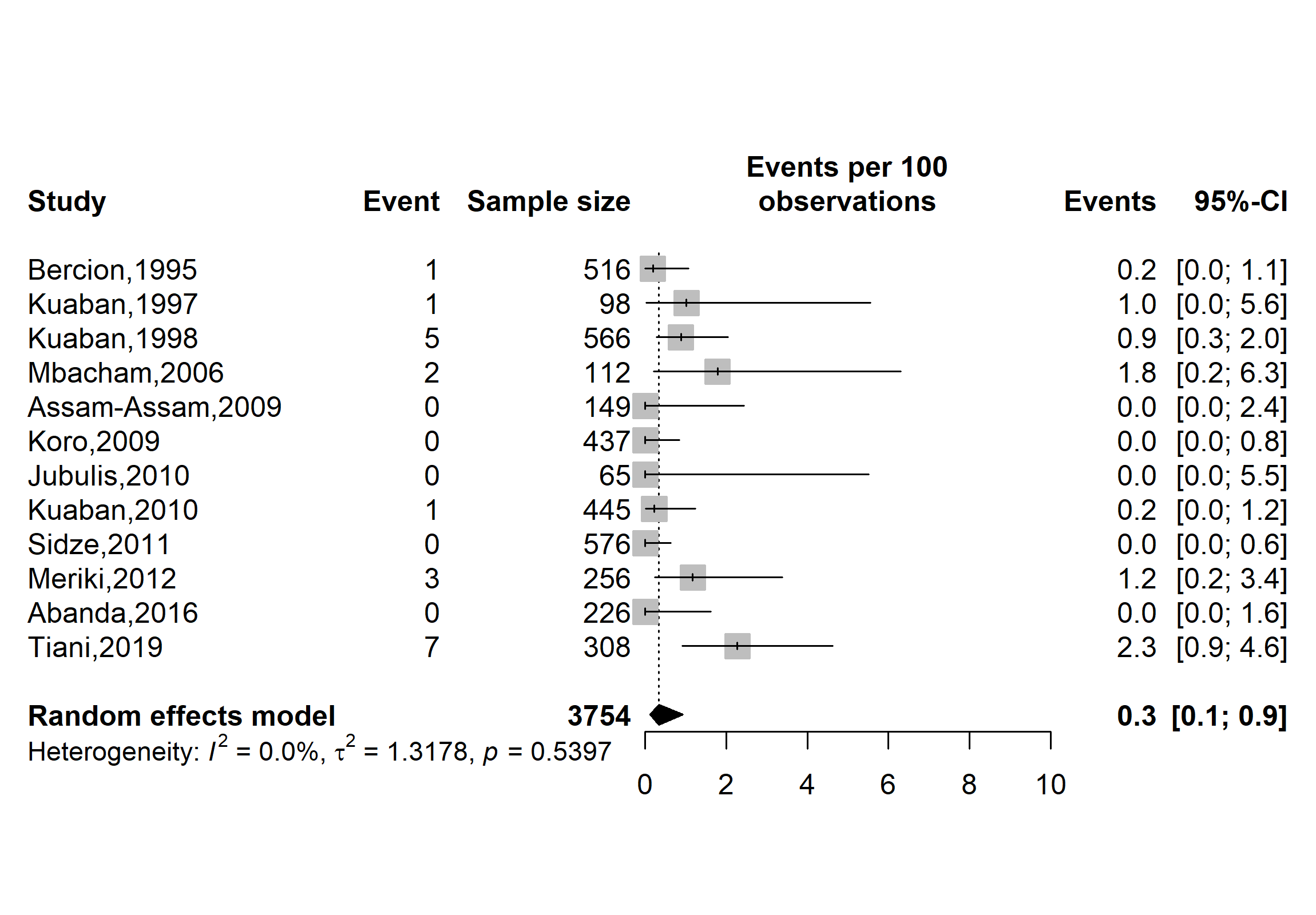

**Fig. S36** Pooled prevalence of ethambutol monoresistance among tuberculosis patients in Cameroon, 1995–2019

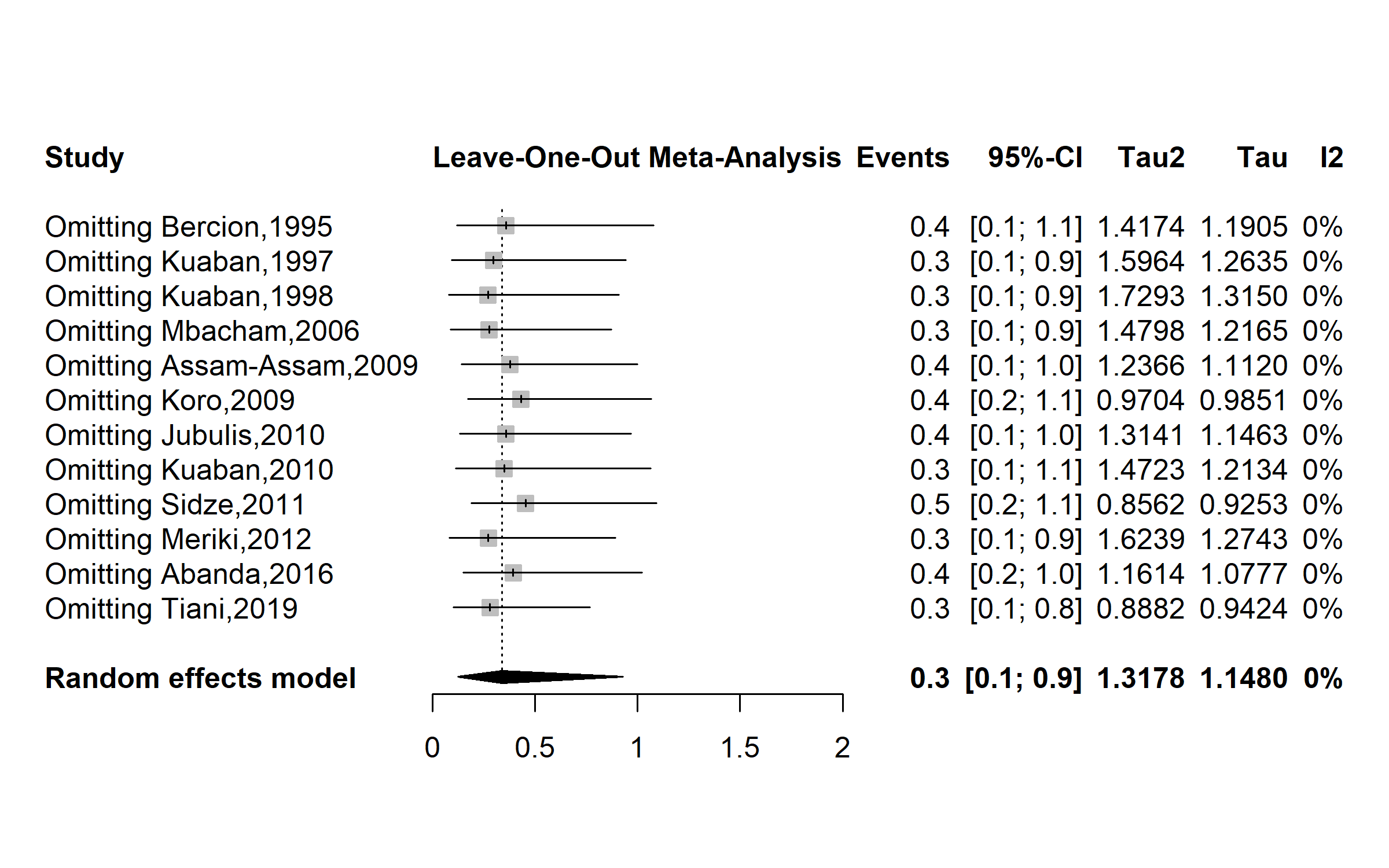

**Fig. S37** Sensitivity analysis of the pooled prevalence of ethambutol monoresistance among tuberculosis patients in Cameroon, 1995–2019

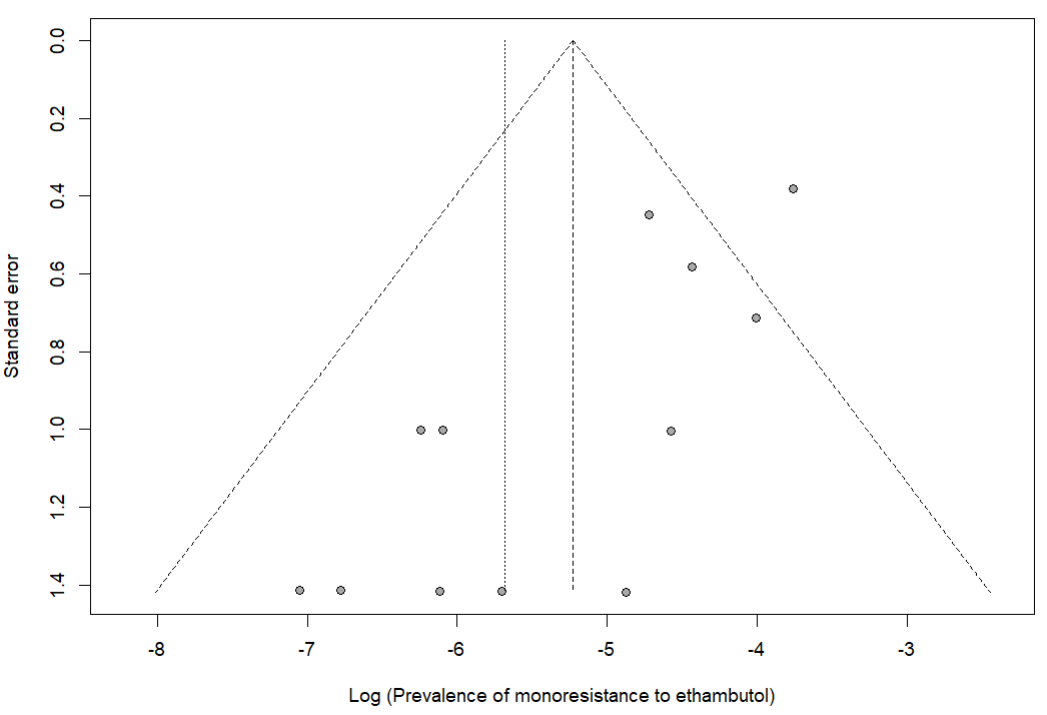

Egger’s test *p*-value = 0.002

Begg’s test *p*-value = 0.273

**Fig. S38** Publication bias of study reports assessing the pooled prevalence of ethambutol monoresistance among tuberculosis patients in Cameroon, 1995–2019

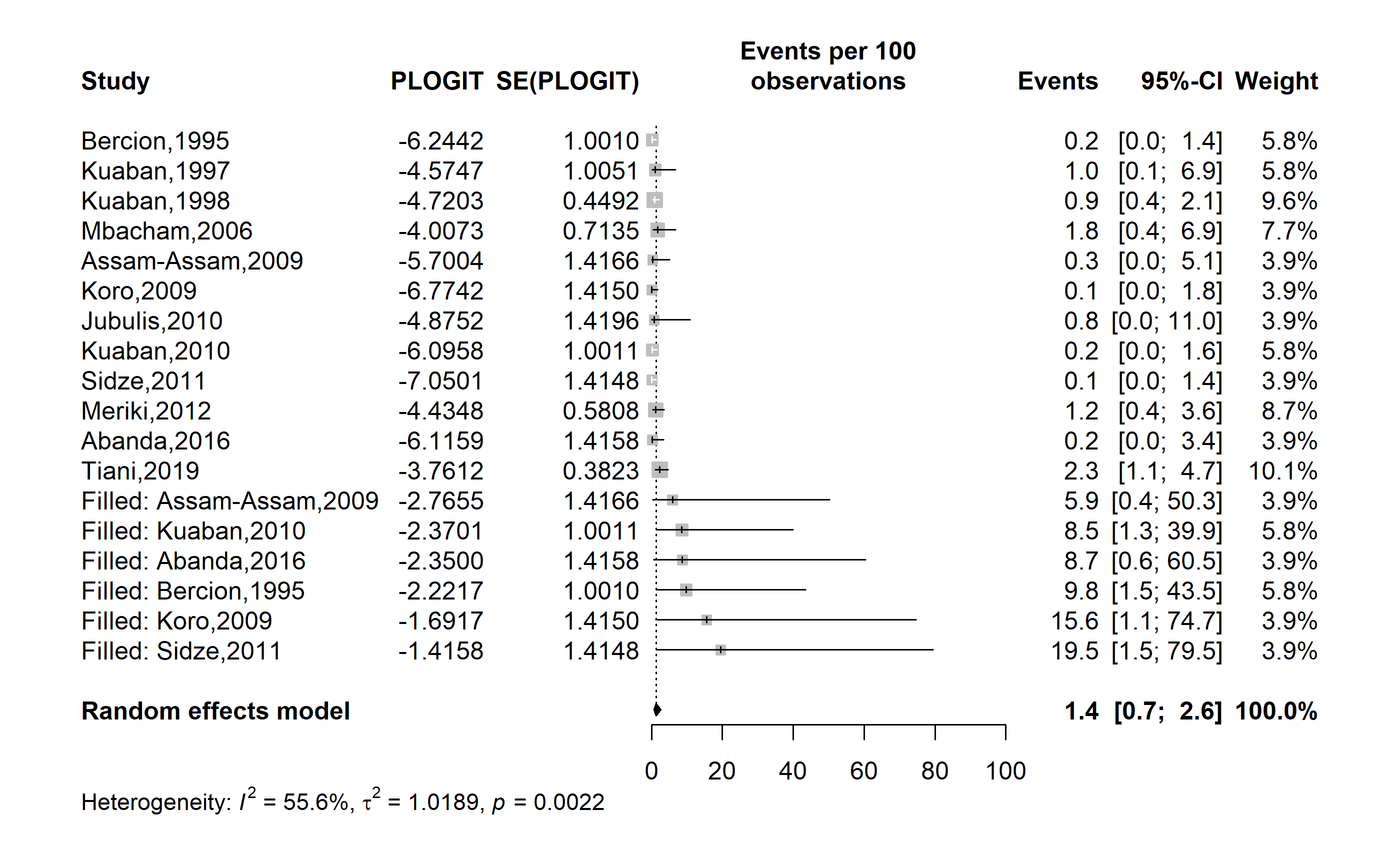

**Fig. S39** Trim-and-fill analysis adjusting for publication bias among studies assessing the pooled prevalence of ethambutol monoresistance among tuberculosis patients in Cameroon, 1995–2019

**Monoresistance to streptomycin**

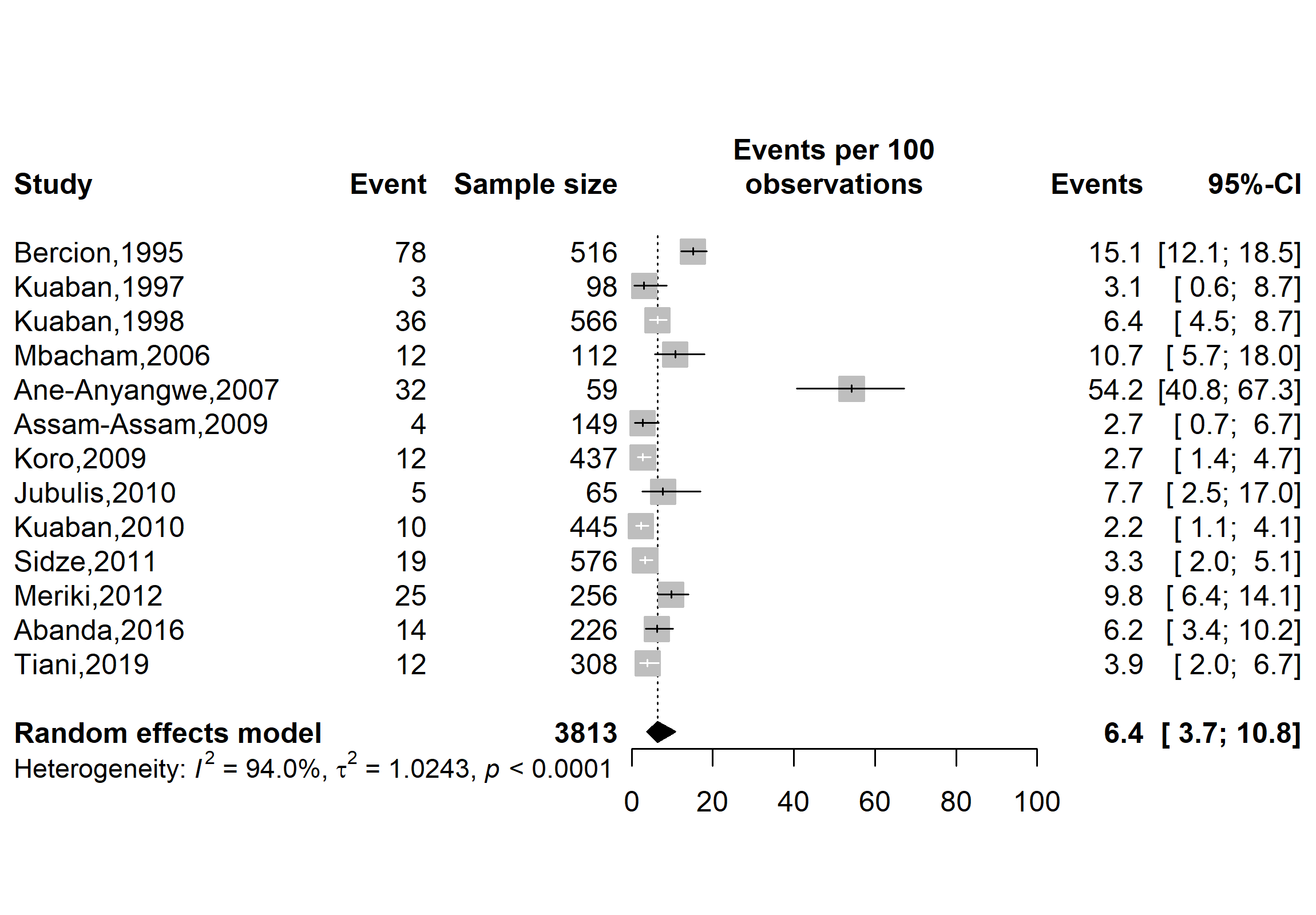

**Fig. S40** Pooled prevalence of streptomycin monoresistance among tuberculosis patients in Cameroon, 1995–2019

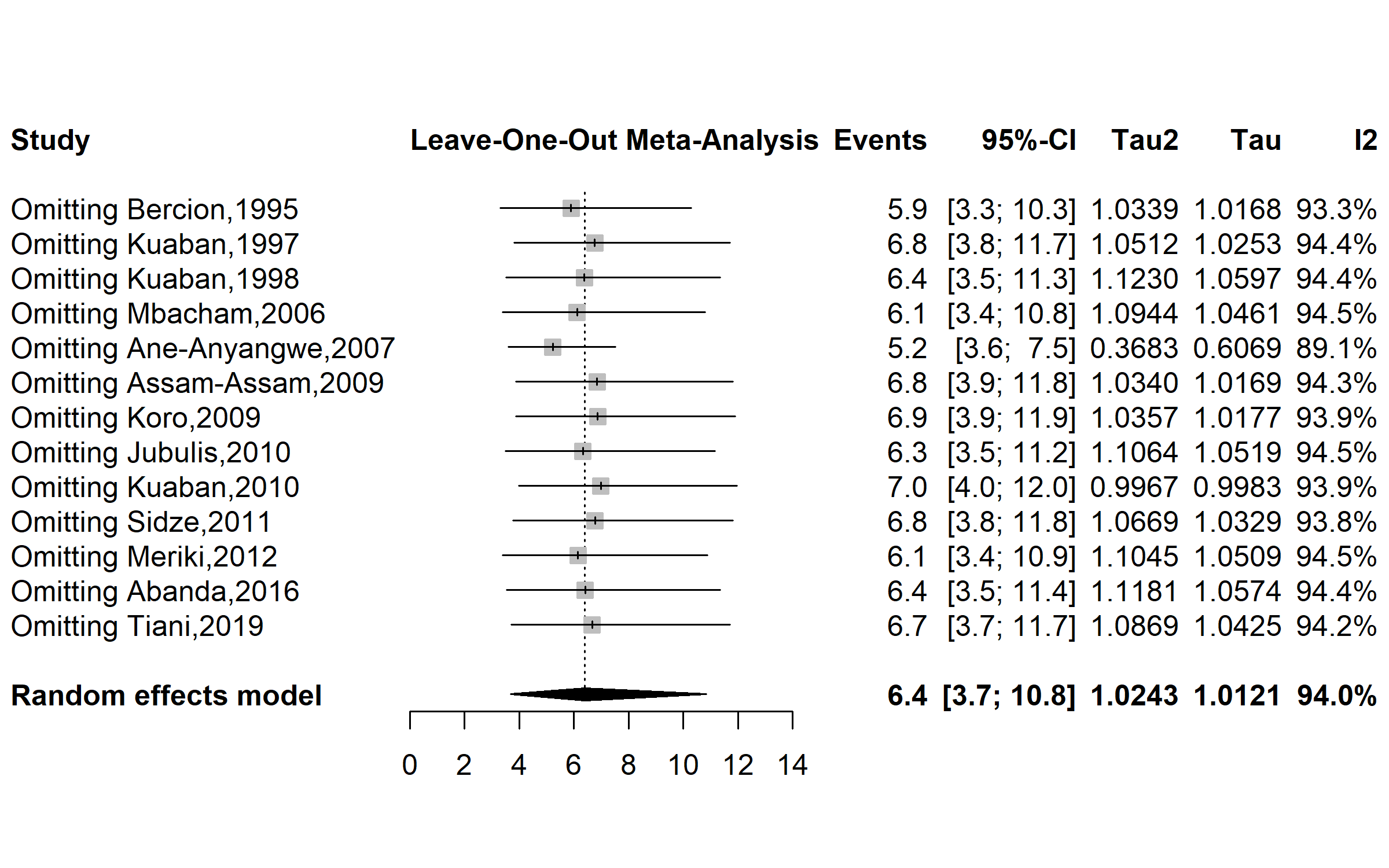

**Fig. S41** Sensitivity analysis of the pooled prevalence of streptomycin monoresistance among tuberculosis patients in Cameroon, 1995–2019

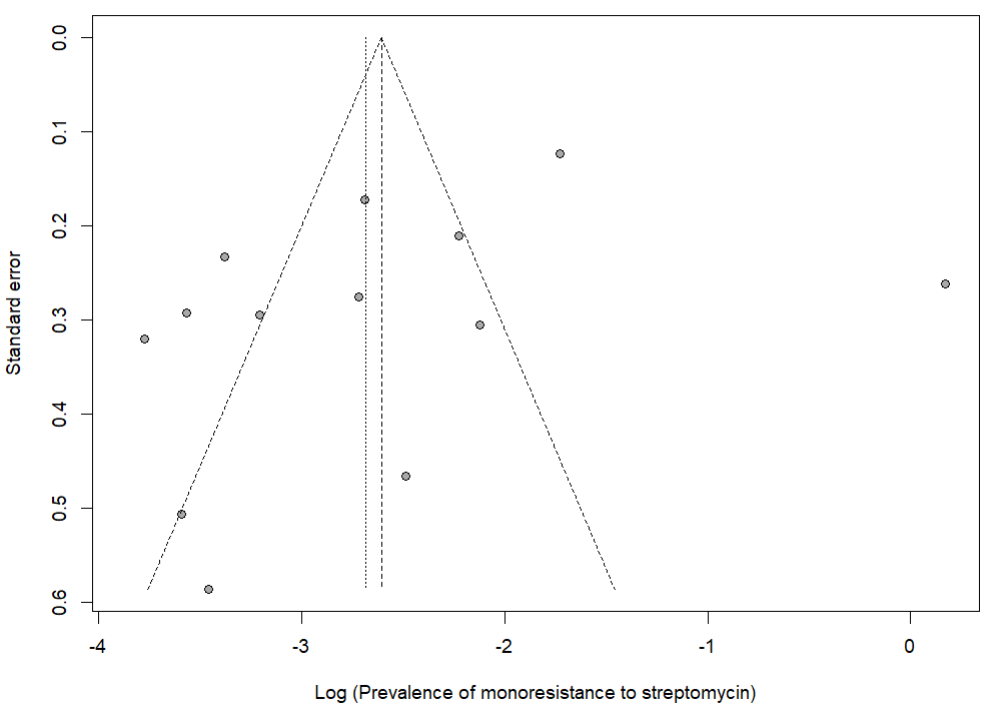

Egger’s test *p*-value = 0.174

Begg’s test *p*-value = 0.464

**Fig. S42** Publication bias of study reports assessing the pooled prevalence of streptomycin monoresistance among tuberculosis patients in Cameroon, 1995–2019

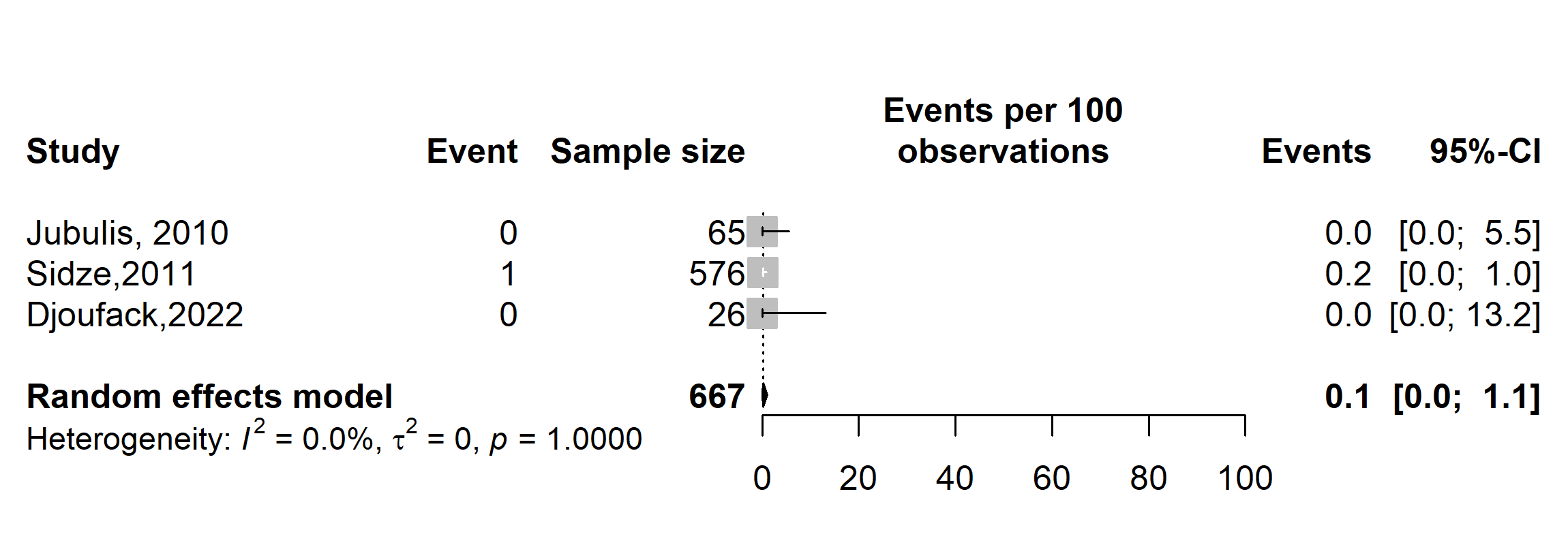

**Fig. S43** Pooled prevalence of fluroquinolones monoresistance among tuberculosis patients in Cameroon, 2010-2022

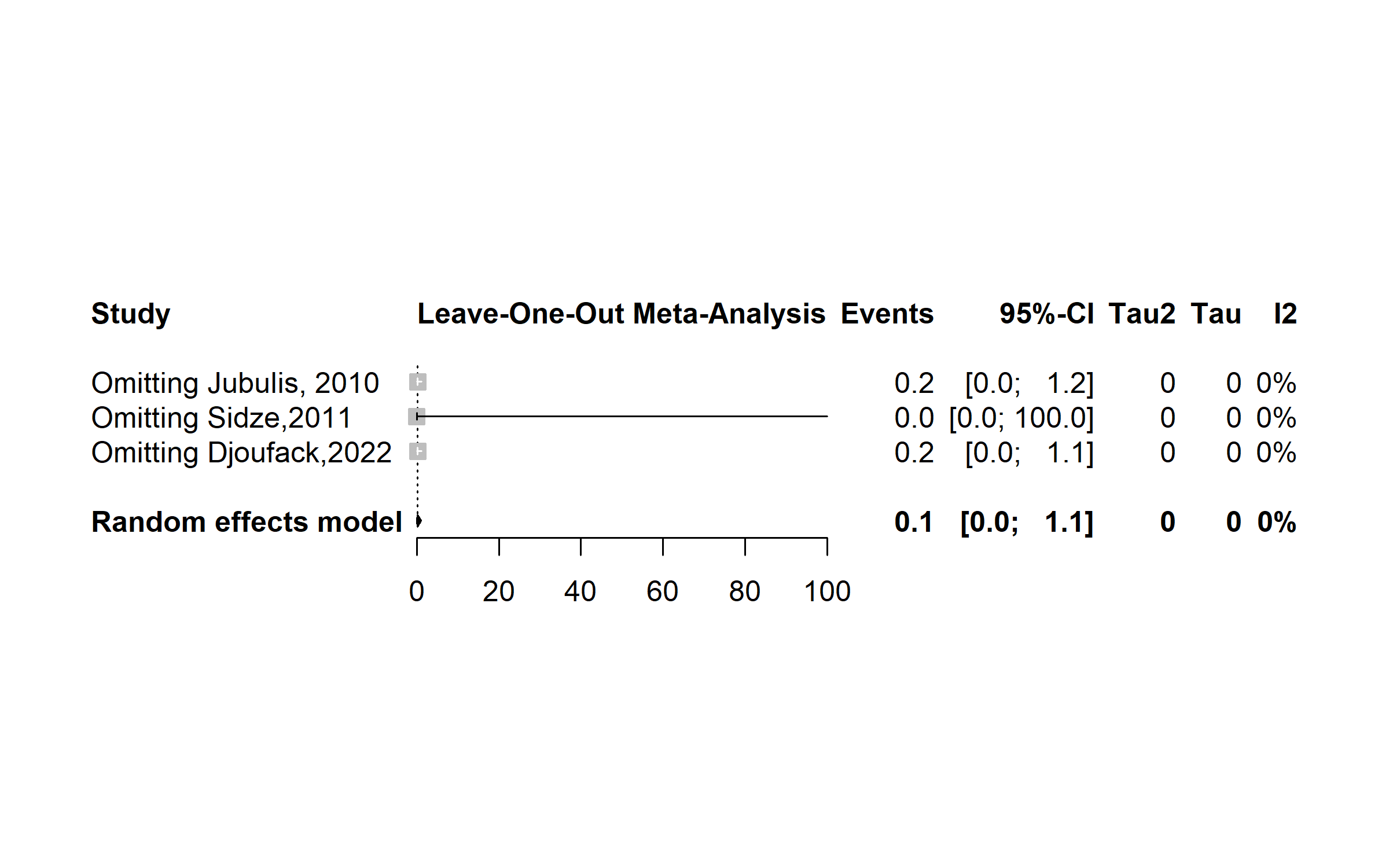

**Fig. S44** Sensitivity analysis of the pooled prevalence of fluroquinolones monoresistance among tuberculosis patients in Cameroon, 2010-2022

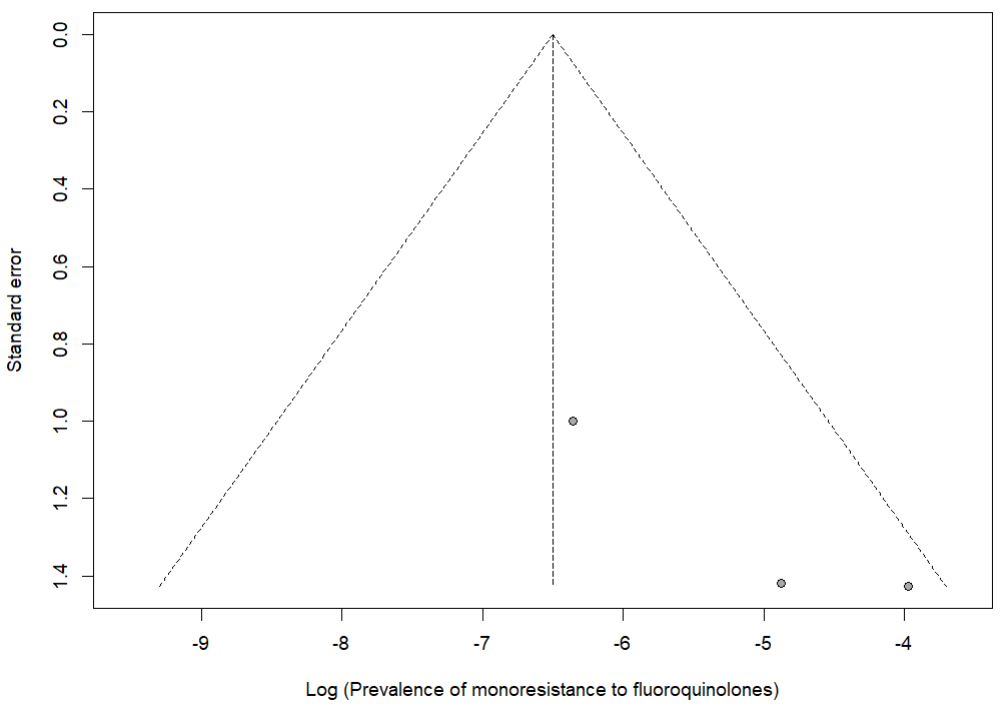

**Fig. S45** Publication bias of study reports assessing the pooled prevalence of fluoroquinolones monoresistance among tuberculosis patients in Cameroon, 1995–2019

**Predictor of resistance to anti-tuberculosis drugs**

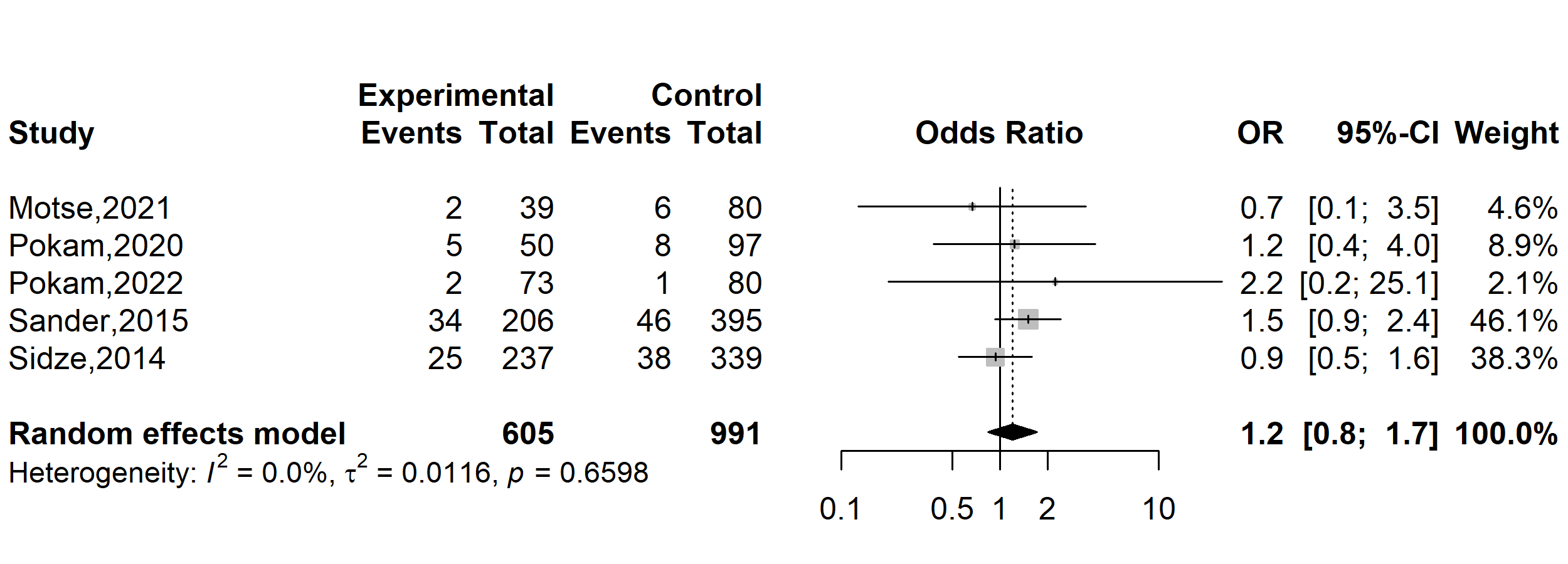

**Fig. S46** Pooled odds ratio of anti-tuberculosis drugs resistance in Cameroon, 1995-2022 (Gender: Female vs. Male)

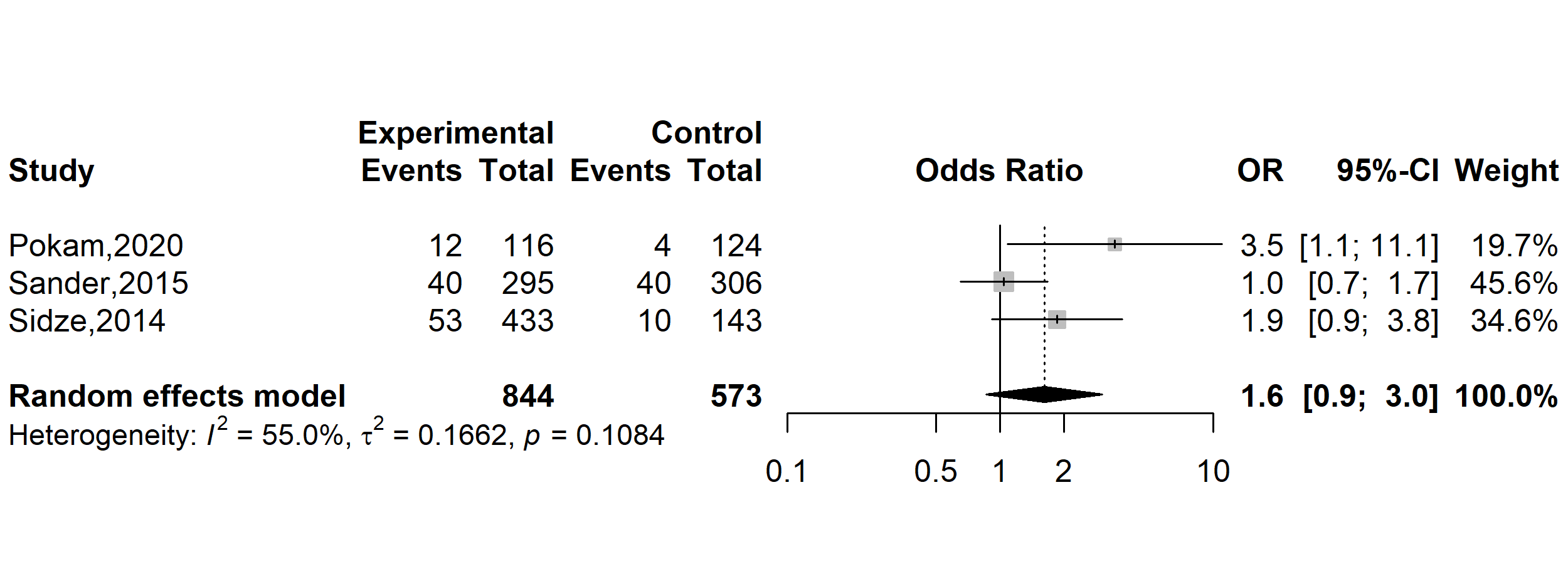

**Fig. S47** Pooled odds ratio of anti-tuberculosis drugs resistance in Cameroon, 1995-2022 (Age: ˂ 25 vs. 25+ years)

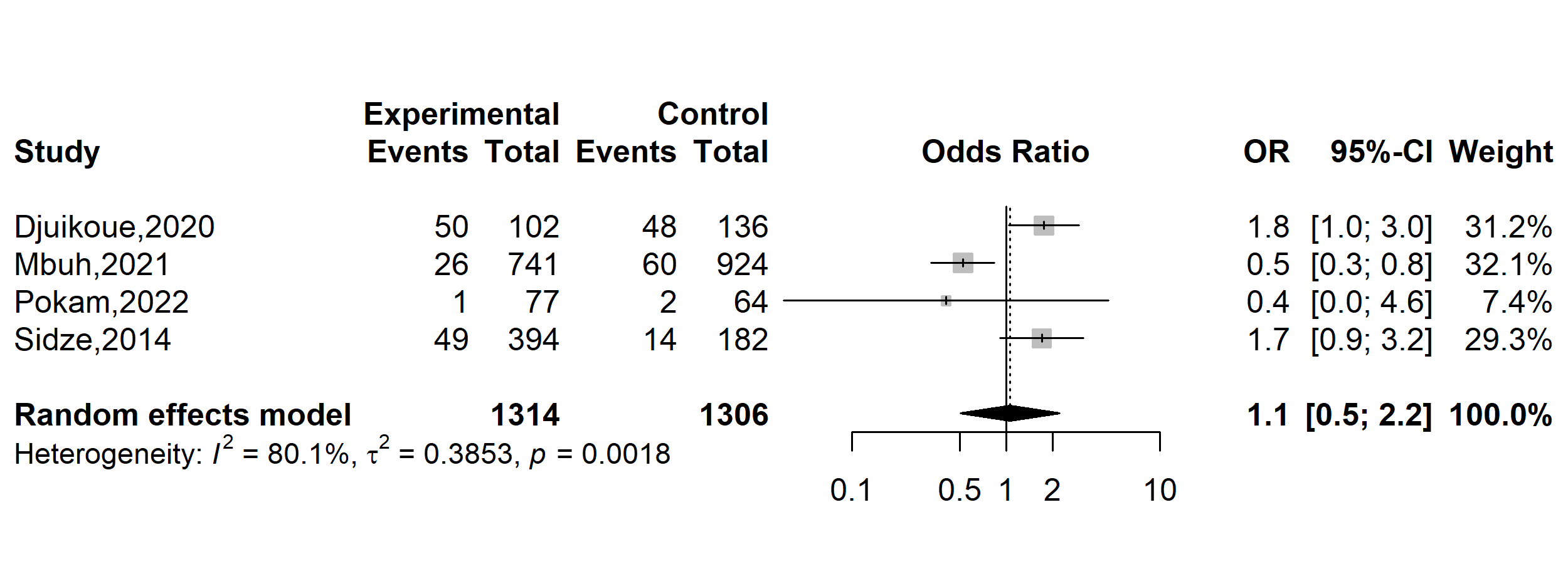

**Fig. S48** Pooled odds ratio of anti-tuberculosis drugs resistance in Cameroon, 1995-2022 (Marital status: Single vs. Married)

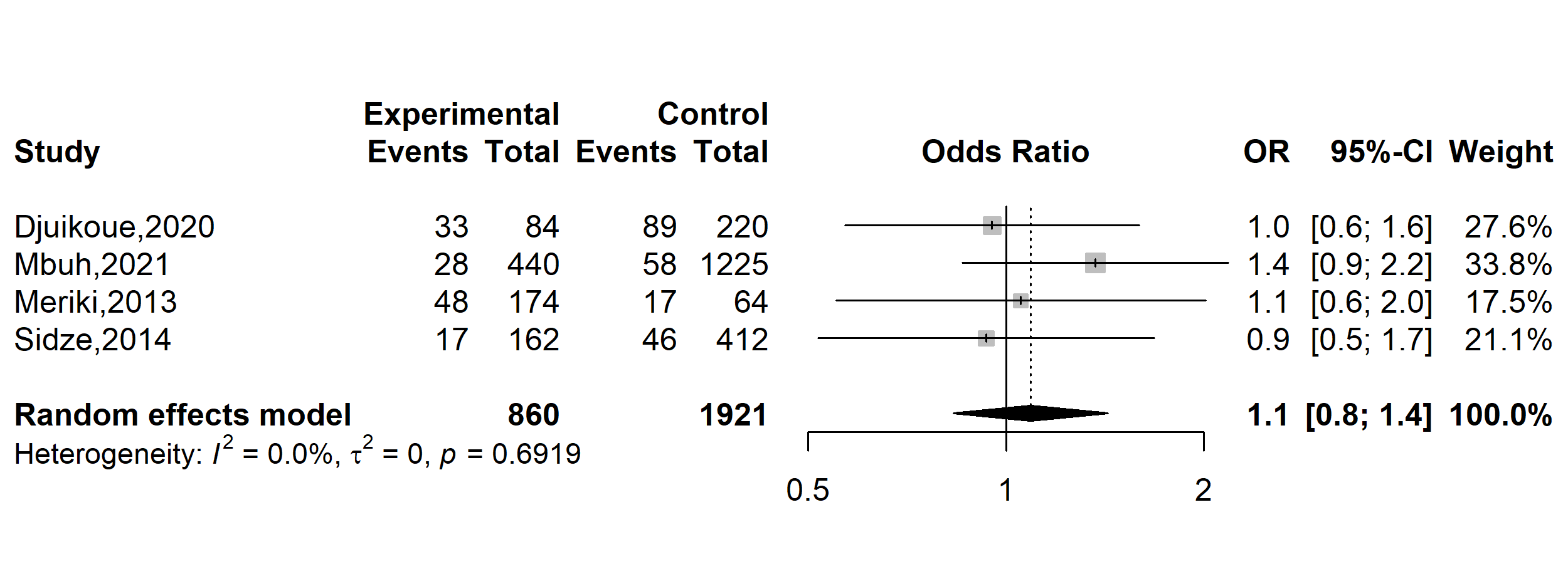

**Fig. S49** Pooled odds ratio of anti-tuberculosis drugs resistance in Cameroon, 1995-2022 (Human immunodeficiency virus status: Positive vs. Negative)

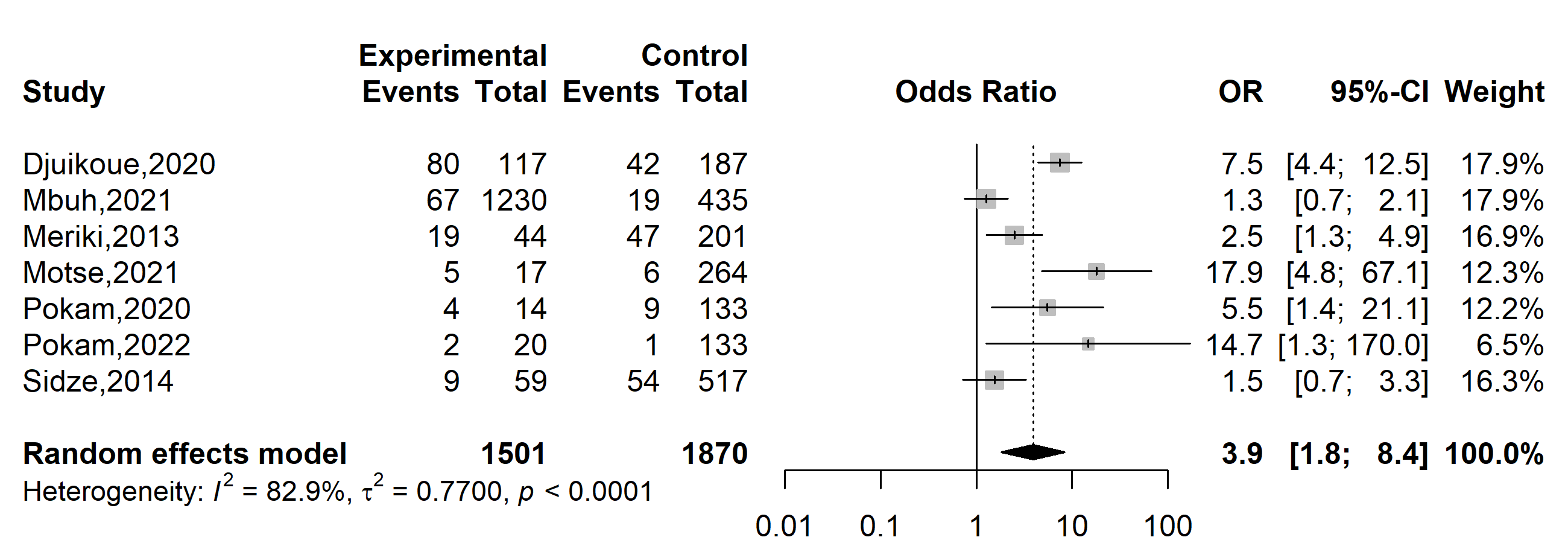

**Fig. S50** Pooled odds ratio of anti-tuberculosis drugs resistance in Cameroon, 1995-2022 (Previous tuberculosis infection: Yes vs. No)

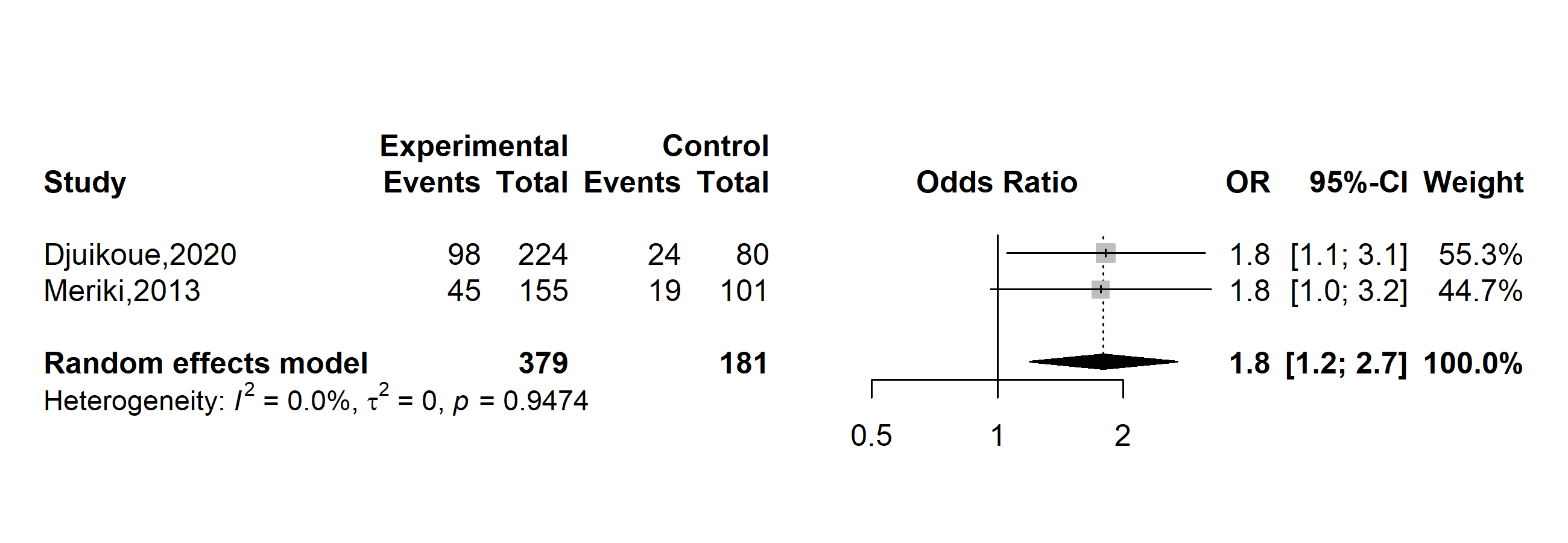

**Fig. S51** Pooled odds ratio of anti-tuberculosis drugs resistance in Cameroon, 1995-2022 (Alcohol consumption: Yes vs. No)

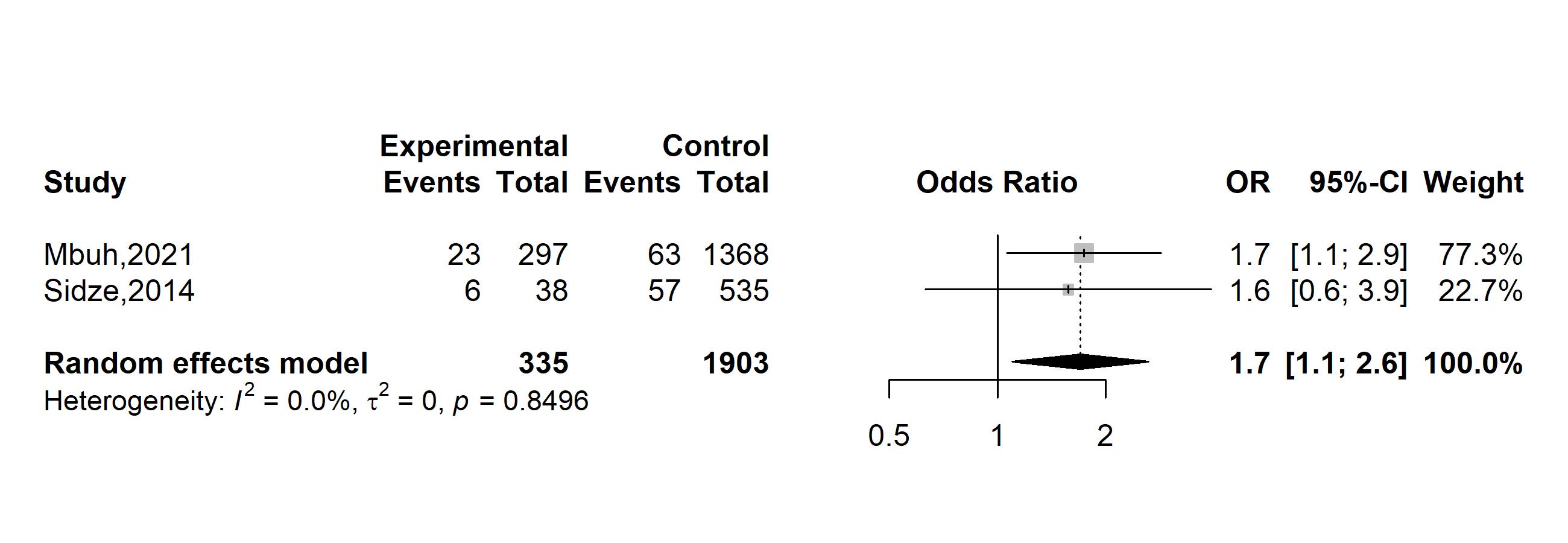

**Fig. S52** Pooled odds ratio of anti-tuberculosis drugs resistance in Cameroon, 1995-2022 (Notion of incarceration: Yes vs. No)
